# ChatGPT in Dermatology: A Comprehensive Systematic Review

**DOI:** 10.1101/2023.06.11.23291252

**Authors:** Irene S. Gabashvili

## Abstract

**Background:** In recent years, the field of dermatology has adopted the latest technologies to enhance patient care and medical education. Mobile technology and social media platforms have revolutionized the delivery of services, and AI-based procedures are poised to become part of dermatologists’ daily routines. There are already numerous papers on the use of the latest conversational AI tool, ChatGPT, in dermatology, and a systematic analysis of these studies can yield valuable insights.

**Objective:** To comprehensively evaluate the literature on the various applications of ChatGPT in dermatology and related areas.

**Methods:** We searched PubMed, Cochrane Library, EuropePMC, medRxiv, arXiv, bioRxiv, Dimensions AI, Semantic Scholar, and Google Scholar, to obtain articles published up until May 15, 2023. The eligibility criteria focused on studies examining the use of ChatGPT in dermatology-related areas. To address the risks of bias, we employed a meticulous selection process, incorporating diverse information sources, including preprints, in multiple languages. In addition to full text articles, acknowledgments and supplemental material were also examined to ensure a thorough analysis. The synthesis of findings utilized network analysis and thematic synthesis methodologies.

**Results:** There was a total of 87 manuscripts that fulfilled eligibility requirements. Over a third of them (36%) acknowledged the assistance of ChatGPT in writing, data analysis or software development. About a quarter (24%) were case reports describing dermatological manifestations and complications. ChatGPT demonstrated successful performance answering questions related to dermatology, ranging from excellent in cancer to barely passable in specialized and lesser-known dermatology areas, although its performance improved with GPT 4. There are advancements in interactive learning, integrations with image-based AI, and enhancing language models for dermatology applications.

**Conclusions:** There has been a remarkable surge in the adoption of ChatGPT in areas related to dermatology, especially in writing case reports. As researchers are aware of safety and uncertainty, a continued feedback loop for reporting errors is crucial for the ongoing improvement and training of AI models, ensuring their reliability and effectiveness in the field.

## Introduction

### Background

The dermatological field has been undergoing a significant transformation in recent years, with the adoption of digital technologies and artificial intelligence (AI) revolutionizing image-based diagnostics and assessment [1]. Mobile technology, including smartphones and instant messaging, are transforming the delivery of dermatology services by providing easy-to-use and cost-effective tools for remote consultation and diagnosis [2].

Social media platforms like TikTok™ have become an important source of information for patients seeking dermatology-related information, as well as for the new generation of medical students, who are “digital natives” immersed in online learning [3, 4]. The prevalence of social media integration in private practices, residency programs and research journals has increased in recent years [5].

Gamification and serious games are being introduced in dermatology education to boost learning motivation and outcomes [6]. AI has the potential to improve the allocation of high-quality medical resources and promote the formation of medical consortia, resulting in a more efficient healthcare system. ChatGPT, the latest advancement in AI, has generated significant interest in the healthcare and biomedical fields [7, 8] due to its potential to assist in medical education, research, and clinical management. The use of ChatGPT in dermatology is still in its early stages, but there are already a considerable number of relevant papers. A systematic analysis of these studies can provide valuable insights into the potential benefits and limitations of Large Language Models (LLMs) in this field.

### Objective

The goals of this study are to evaluate the literature concerning the applications of ChatGPT and its potential impact on various fields within dermatology. This will be achieved through a systematic review of and bibliometric analysis of relevant studies. The findings from these analyses will provide insights into the potential impact of ChatGPT in different subcategories, identify promising future applications, and highlight areas that require further research.

## Methods

### Data Sources and Search Strategy

In this study, an extensive search was conducted across multiple databases using keywords relevant to dermatology, including terms related to skin, hair, and nails. Specific dermatological conditions such as psoriasis, dermatitis, acne, vitiligo, alopecia, lichen planus, as well as topics like cosmetics, eczema, malodor, hyperhidrosis, bromhidrosis, olfactory reference syndrome, rosacea, and sunburn were also included in the search. These terms were examined for their co occurrence with the keyword “ChatGPT”.

Repositories searched for relevant articles included EuropePMC, Semantic Scholar, Pubmed, Dimensions AI, Medrxiv, Biorxiv, and Google Scholar.

### Protocol Registration

The protocol was registered for systematic reviews of health-related outcomes on PROSPERO on April 15 2023 (registration DOI: CRD42023417336, available from: https://www.crd.york.ac.uk/prospero/display_record.php?ID=CRD42023417336. Search strategy was modified to narrow the scope of the review, based on the criteria outlined above.

### Study Selection

The inclusion and exclusion criteria encompassed studies in all languages, provided that sufficient information (from abstract and full text) was available to determine their eligibility. The exclusion criteria were (1) articles with non-dermatological applications of dermatology-related keywords, (2) articles mentioning ChatGPT, but not its applications in dermatology-related contexts (2) short abstracts from conferences unrelated to dermatology, (3) news articles, press releases, blogs, corrigendum documents and (4) retracted publications.

Studies published in all languages were considered. Translation tools provided by Google and OpenAI were employed for non-English studies. multilingual text analysis

To minimize bias in our review selection, we employed a team approach consisting of the human author, ChatGPT and Bard. In addition, we implemented a rigorous search strategy encompassing multiple databases, which allowed for redundancy. We also applied various filters and employed checkers to ensure the inclusion of a wide range of potentially relevant studies, thereby reducing the potential for selection bias.

At first step, titles were checked for relevance and duplication. Similar titles belonging to different versions of the same study were identified and the most recent and complete data was included.

The human author screened the list of unique titles, article types and journal names and abstracts and read full text retrieved via APIs, electronic and manual searches. Titles, abstracts and portions of full text were then screened by ChatGPT according to the same criteria and categorized under one of dermatology subcategories, if applicable. Any discrepancy was solved by repeated review/better prompt engineering and text pre-processing in order to reach a consensus.

### Data Extraction & Synthesis

The following data items were extracted from the articles in our systematic review: External IDs including DOI: Digital Object Identifier, PMID: PubMed ID, PMCID: PubMed Central ID, Title, Abstract, Acknowledgements, Funding source, MeSH terms, Fields of Research, Publication Date, Publication Type, the names and affiliations of the authors who contributed to the article, the number of times the article has been cited by other publications, information on recent citations received by the article. RCR: Relative Citation Ratio, a measure of the citation impact of an article relative to others in the same field, Impact Factor of the journal in which the article was published, a link to the original source of the article, along with the details of literature search and data sources.

In addition to processing the full text and extracting relevant snippets related to taxonomical classification, the quality assessment questions for the included reviews covered various aspects, including review selection and inclusion criteria, assessment of publication bias, discussion of heterogeneity tests, and comparability of included reviews in terms of eligibility criteria, study characteristics, and primary outcome of interest.

For data upload and analysis, Python scripts were utilized, incorporating modifications from [7] for the Semantic Scholar API.

To facilitate data analysis and visualization, we employed VOS Viewer software [9], specifically Version 1.6.18. VOS Viewer enabled us to perform descriptive statistics and scientific mapping, allowing exploration and visualization of author collaboration networks, keywords, and connections between analytical themes. Widely used in academic literature, this tool employs text mining algorithms to identify noun phrases from publication titles and abstracts, enabling the construction of networks, clusters, and heatmaps.

In addition to network analysis, we employed a thematic synthesis approach to identify and analyze themes or patterns across the included studies. This involved coding and categorizing the data to extract common themes or concepts, providing a comprehensive understanding of the research topic.

Data extraction was conducted by the human reviewer using standardized summary tables, and the extracted data were also categorized by ChatGPT and Bard. In case of any disagreements, a re-review was performed.

The human reviewer coded all the data, and ChatGPT and Bard were asked to perform the same task. Disagreements were resolved through the re-review of data by the human reviewer, who reformulated prompts as needed until an agreement was reached. Once common themes and topics were identified in the studies, the codes were grouped into descriptive themes. These themes were organized into a table, which facilitated understanding of the similarities and differences between the individual studies. In the final phase, interpretive or analytical themes were identified based on the insights gained from the previous phases.

## Results

### Overview of included studies

Figure 1 presents an overview of the study selection process. Initially, a total of 835 studies were identified from multiple databases, encompassing various languages such as English, German, French, Japanese, Polish, Portuguese, and Slovenian. Among these, 263 studies were excluded as duplicates. During the pre-screening phase, 91 studies were deemed ineligible due to reasons such as being blog posts or news articles. Furthermore, during the screening of publication titles, types, and publisher/journal names, 106 studies were excluded as they were conference proceedings or books unrelated to the topic of interest. Additionally, the DOIs of 2 studies were found to be invalid.

**Figure 1.**
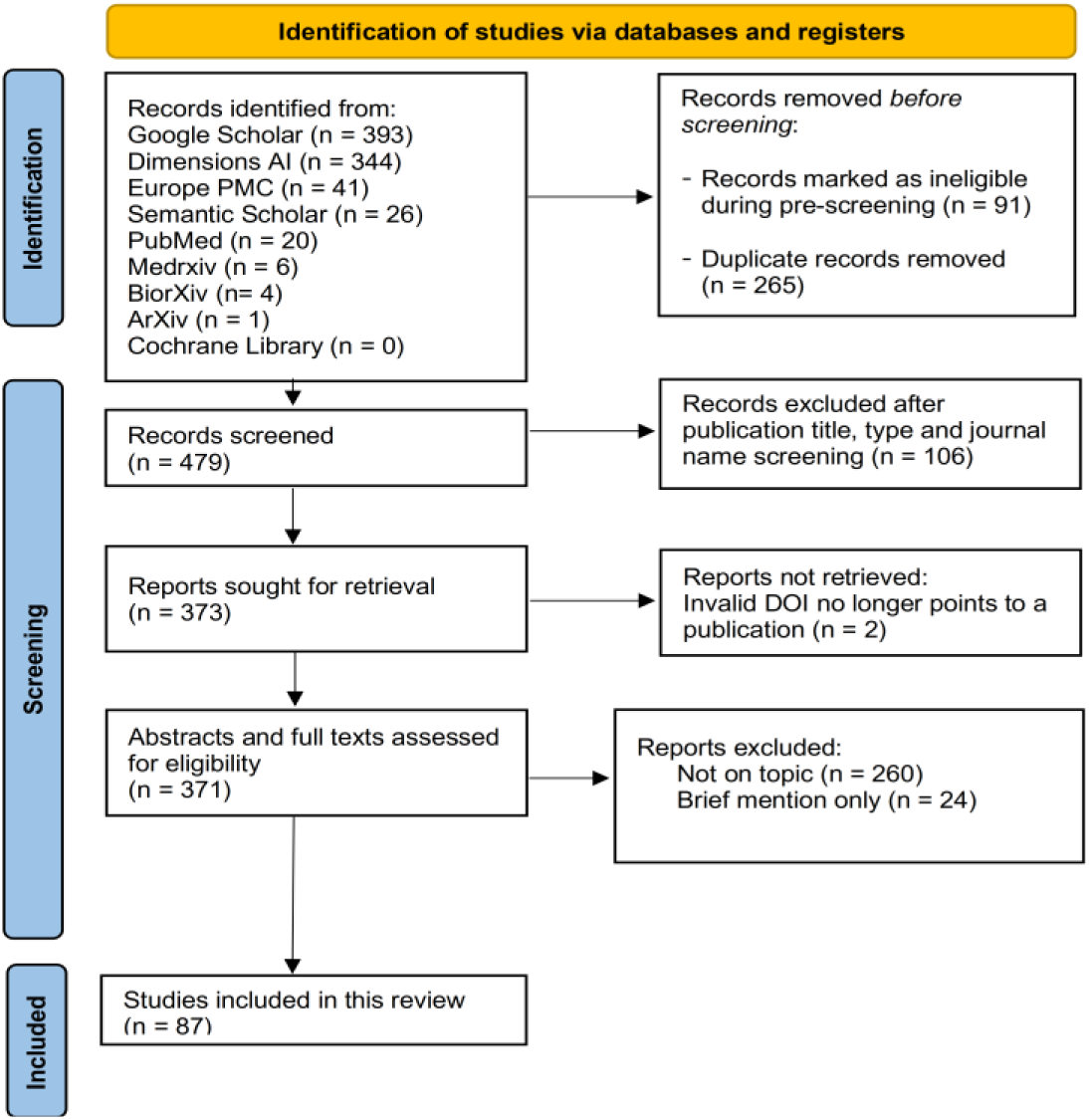
Prisma flow diagram for study inclusion

The majority of the remaining 373 studies were arXiv preprints, Cureus case reports and articles published in dermatological journals, mostly retrieved through Dimensions and Google Scholar searches. Besides arXiv, preprints from medrXiv, bioRxiv, psyarxiv, edarxiv, ResearchGate, osf, SSRN electronic journal, and JMIR preprint sites were included in this list. Out of 373, 21 papers were published in dermatology journals.

Further filtering was conducted, resulting in the exclusion of 284 additional articles. Some of these studies only briefly mentioned ChatGPT or its applications in dermatology. Other reports were found to use dermatology-related keywords in a non applicable context.

Examples included discussions on the natural tint of the skin in the context of the science of deduction, psychological contexts related to skin color, philosophical discussions on human beings losing skin sensation, ChatGPT generating prompts like “with porcelain skin and flowing hair”, comments like “ChatGPT’s brilliance is only skin deep”, and case reports mentioning the implantation of pacemakers under the skin of patients or skin swabs taken. It is noteworthy that several papers, while not meeting the inclusion criteria, discuss the application of ChatGPT and large language models (LLMs) in topics tangentially related to dermatology. For example, thermoregulation or using knowledge of dermatological elements to create metaverse characters or write science fiction stories with ChatGPT. We chose not to exclude four articles labeled as “Brief mention only” as they were consistent with the other 83 papers that met the inclusion criteria. Hence, a total of 87 articles were included in this review (Appendix 1: 84 in English, 1 in Japanese, 1 in German and 1 in Spanish; 34(39%) preprints), and their analysis is detailed in the subsequent sections.

### Classification frameworks

The selected articles in this systematic review demonstrate significant heterogeneity, as acknowledged in previous broader reviews on ChatGPT applications in healthcare [10–13]. The earliest study [10], with a literature search cutoff of 16 February 2023, identified “academic/scientific writing” as the predominant theme in over half of the records. Scientific research benefits were mentioned in a third of the records, including the analysis of large datasets such as electronic health records and genomic data. Benefits in healthcare practice were mentioned in a quarter of the records, such as personalized medicine, disease risk prediction, streamlined clinical workflows, improved diagnostics, cost savings, and enhanced health literacy. Educational benefits in healthcare disciplines were mentioned in just over a tenth of the records, including the generation of accurate clinical vignettes.

Li et al. [11], in their study conducted on paper published before 3/20/2023, proposed an application-oriented and user-oriented taxonomy to categorize the selected publications. For instance, papers on scientific writing, literature reviews, and research idea generation were grouped under the category of “medical research”. The “consultation” category included papers where ChatGPT was utilized for medical consulting in both corporate and individual settings. The “clinical workflow” category encompassed applications such as diagnostic decision-making, treatment recommendations, and the generation of clinical documentation. They also proposed to classify papers by their depth from generic comments about potential applications (Level 1), to comments with one or more use cases of ChatGPT in a specific scenario (Level 2) to Qualitative and quantitative evaluation of ChatGPT’s answers (Level 3). Muftić et al. [12] combined level 1 and some level 2 papers into one category and focused on 21 studies of the highest level including cross-sectional observational studies and performance evaluations.

Goedde et al. ([13], on papers published by the end of March 2023) found that about half of all articles were brief statements like editorials, or letters to the editor, while about a third were assays or commentaries. Studies, reviews/meta analyses, and case reports were less frequent.

The classification framework employed in this review utilized suggestions provided in these papers as well as potential utilization of ChatGPT by doctors, staff and patients discussed by a dermatologist [14]. We classified these applications as administrative dermatology (patient letters, standardized reports), education (patient’s leaflets), consultation (diagnosis, management) and academic writing.

### Interpretive themes

The 87 articles selected for the systematic review on the utilization of ChatGPT in dermatology shed light on various aspects of its involvement in the field. Four prominent themes emerged from the analysis, namely perspectives, utilization, evaluation, and development. Within the “Perspectives and Recommendations” category, a range of opinion pieces, editorials, and research papers offered recommendations, guidelines, and potential applications for ChatGPT’s utilization.

The “Utilization” category comprised papers that explicitly acknowledged the assistance of ChatGPT in enhancing readability, expediting the review process, developing computer programs, or enabling data analysis without coding. The “Evaluation” theme encompassed studies employing rigorous statistical analyses, as well as qualitative evaluations through exemplary cases. Lastly, the “Development” theme encompassed papers that explored alternative technologies to ChatGPT, focused on specific applications, and discussed fine-tuning and additional training methods.

VOSViewer’s clustering analysis of titles, abstracts, and full-text snippets revealed several highly connected general themes in the field of dermatology. These themes include Dermatology Practice and Patient Care, which encompasses patient cases, medical education, and decision-making. Another theme is Clinical Practice and Treatment Options, focusing on the evaluation, adoption, limitations, and improvements in dermatological practices. Additionally, smaller clusters were identified, such as Dermatology-related Basic Research, Cosmetic Dermatology and Plastic Surgery, Infection and Pediatric Dermatology, Dermatology-related Misinformation, and Specific Dermatological Conditions and Treatments, including sensitive topics like Alopecia and Hair Related Topics. The presence of diverse and overlapping themes makes it challenging to visualize and categorize them into well defined clusters. However, the clustering of labels provides a clearer differentiation of major topics, as depicted in Figure 2. In the following sections, we will discuss these prominent themes in more detail.

**Figure 2.**
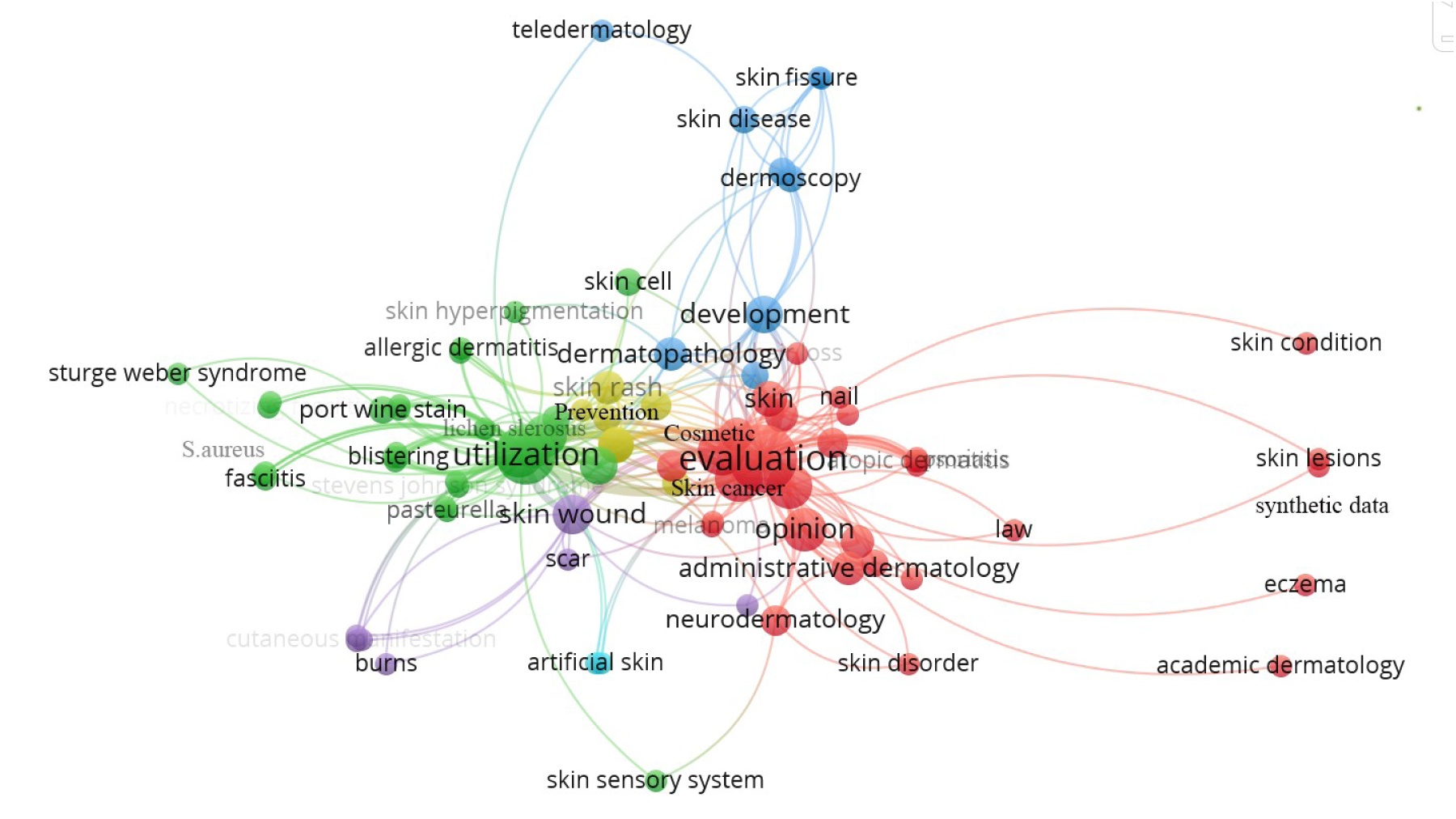
Categories and Keywords clustered/visu
alized with VOSViewer. Red: Evaluation & Opinion, Green: Utilization, Blue: Development; Aquamarine: Basic Research, Yellow: Prevention/Skincare, Purple: Skin injuries.

### Perspectives

The 10 articles from this category (see Appendix 1) explore the potential benefits and ethical considerations associated with using ChatGPT-like tools for tasks such as diagnosis prediction, patient care, practice efficiency, and medical education. They highlight the need for regulation, the limitations of relying solely on AI-generated recommendations. the importance of considering patient safety, monitoring AI development, and the implications for medical education. An example is an overview of positive and gray zone applications of ChatGPT in dermatology practice [14].

### Evaluation

In our systematic review, a significant portion of the selected articles (almost half, 47%) focused on evaluating ChatGPT’s writing capabilities and its application in various medical contexts, including dermatology. The evaluation encompassed tasks such as generating clinical case reports, creating persuasive messages, preparing medical documents, and providing answers to medical and dermatology-related questions. 22 out of 41 papers (54%) represented quantitative performance evaluations and cross-selection observational studies vs papers based on lower level evidence (as defined in earlier reviews of healthcare applications [11,12])

The findings revealed several noteworthy outcomes. Firstly, dermatology case reports generated by ChatGPT received higher scores in overall quality and readability compared to reports written by humans [15]. Collaborations between AI and human experts improved the readability of medical documents, such as surgical consent forms and insurance letters, without compromising clinical detail. Additionally, ChatGPT demonstrated potential in medical evidence summarization and the generation of persuasive narratives. Overall, a total of 37 studies (43%) either utilized ChatGPT for writing purposes or evaluated its writing abilities.

ChatGPT showcased its proficiency in passing medical exams across various languages. Notable evaluations included the Spanish MIR 2022 exam, the US Plastic Surgery Inservice Training Examination (PSITE), the United States Medical Licensing Exam (USMLE), Japanese National Nurse Examinations, the National Medical Licensing Examination (NMLE) in Japan, the New England Journal of Medicine (NEJM) quiz, test based on the National Medical Commission recommended competency-based medical education (CBME) curriculum on microbiology, and the American College of Radiology (ACR) radiation oncology in training (TXIT) exam.

ChatGPT (GPT-4) demonstrated commendable performance on radiation oncology physics exam questions. However, its intrinsic properties limited further improvement when scoring was based on a majority vote across trials. A team of medical physicists outperformed ChatGPT (GPT-4) significantly using a majority vote. Humans’ ability to leverage the collective knowledge or expertise to arrive at the most reliable or accurate response indicates the potential for collaborative work between large language models (LLMs) and specialists such as radiation oncology experts.

A study comparing ChatGPT responses to verified physicians’ responses on Reddit’s r/AskDocs demonstrated that the chatbot responses were of better-quality and more empathetic compared to physicians [16].

Three papers reviewed in this study highlighted instances of hallucinated references within the outputs generated by ChatGPT. In an evaluation involving sixty-six questions from an informatics consult service, with a notable emphasis on internal medicine and dermatology, both GPT-3.5 and GPT-4 were prompted and yielded responses (supported by fictitious references) that aligned with informatics consultations in less than 20% of cases [17]. We also observed this behavior when posing questions regarding lesser-known topics such as microbial basis of PATM. In this instance, ChatGPT 3.5-turbo provided a hallucinated definition of PATM (Psychogenic Attributed Misattribution), while Google Bard, correctly defined it and cited a source [18] along with additional hallucinated references. The performance of large language models (LLMs) varied depending on the medical question being asked, with the best results observed in skin cancer-related topics [19], while specialized dermatology topics like eczema posed challenges, especially when supporting answers with citations [20]. To comprehensively evaluate the effectiveness of LLMs in healthcare, further exploration is needed in prompt engineering, calibration, and customizing these models for specific medical contexts.

In a separate study, the successful application of the ChatGPT-DALL-E pipeline was demonstrated in generating datasets for common thoracic diseases, which can include dermatological manifestations such as fibrosis or nodules [21]. The article emphasized the significance of diverse and well-annotated data in improving the quality of image-based diagnosis. Various techniques were discussed, including data augmentation, transfer learning, federated learning, and generative adversarial networks (GANs), along with the incorporation of domain knowledge using knowledge-guided GANs.

### Utilization

Out of the selected articles, 31 (36%) acknowledged the assistance of ChatGPT in the writing process, data analysis and new tool development. 21 of these (24%) were case reports/case series describing dermatological conditions or dermatological manifestations of other diseases. ChatGPT proved valuable in improving the readability of the reports and providing information in areas where the authors had limited expertise. Pediatricians, for instance, found ChatGPT particularly helpful in the absence of infectious disease specialists on their team [22.]. Additionally, one study utilized ChatGPT in the writing of a dissertation, showcasing its contribution to academic work. Seven articles included in the review were either reviews, editorials or research papers that were written with the assistance of ChatGPT. In these cases, ChatGPT played a role in facilitating the writing process, drafting initial manuscript outline, assisting with data analysis and enhancing the quality of the final publications.

### Development

Traditional large language models could struggle to provide accurate medical responses due to their lack of domain-specific knowledge. To address this limitation, a team of computer scientists has developed the ChatDoctor model, which is fine tuned in the medical domain using Meta’s open-source LLaMA framework. Trained on a dataset of 100k patient-physician conversations, ChatDoctor incorporates real time autonomous knowledge retrieval capabilities, resulting in superior performance compared to ChatGPT, particularly in evaluating various skin lesions [23]. The authors have generously shared the source code, datasets, and model weights to encourage further advancements in the field.

Chinese language-speaking HuaTuo [24] is another open-source model based on LLaMA, trained on skin and hair-related in addition to other health-related questions. The authors introduced a novel SUS metrics to assess LLMs, and HuaTuo outperformed similar models to ChatGPT in terms of “Safety” (potential to mislead), “Usability” (alignment with medical expertise), and “Smoothness” (proficiency as a language model).

For dermatology diagnostics, SkinGPT [25] presents an innovative system that combines MiniGPT-4, an advanced vision-based large language model, with an extensive collection of in-house skin disease images and doctor’s notes. Unlike generic ChatGPT, SkinGPT can be locally deployed and effectively handle sensitive images.

Furthermore, researchers have explored interactive learning of dermatology in a 3D metaverse, enabling users to engage with ChatGPT [6].

Chatbot CHIE [26] was designed to address sensitive inquiries related to cosmetic dermatology.

Two additional papers propose methods to enhance LLMS. One approach involves an over-generate then-filter strategy, where ChatGPT is utilized to identify sensitive domains, specifically tested on questions from preventive dermatology [27]. Another method focuses on fine-tuning a domain-specific language model like BioBERT, comparing it to the general-purpose ChatGPT [28].

These advancements highlight the ongoing efforts to augment the capabilities and applications of language models, which would make AI tools more effective in the field of dermatology.

## Discussion

### Principal Findings

The findings of this review highlight the potential of ChatGPT as a valuable tool in various dermatology-related tasks. ChatGPT can effectively support documentation preparation, scientific writing, decision support, and education in the field of dermatology. These advancements promise to revolutionize patient consultations, treatment planning, and follow-up care.

The analysis of major themes extracted from the selected papers in this review (Fig.2) provides valuable insights into the diverse applications of ChatGPT in dermatology. The largest cluster, categorized as “evaluation/opinion” (depicted in red), encompasses several subcategories within dermatology, including cosmetic dermatology, skin cancers, academic dermatology, and administrative dermatology. The “utilization” cluster (green) incorporates autoimmune dermatology and a wide range of dermatological complications described in case reports related to various diseases. Furthermore, the “Development” cluster (teal) focuses on dermatopathology and telemedicine. Additionally, three smaller clusters emerge, namely preventive dermatology (yellow) encompassing topics like sun exposure and hair loss, wound care (navy blue), and basic research with the potential to develop new treatments for skin conditions (aquamarine).

A notable area of development in ChatGPT’s application is the integration with AI aided dermatological diagnosis based on image recognition. The integration of these two technologies holds promise for advancing dermatological research. In fact, basic biomedical research applicable to dermatology is already utilizing ChatGPT’s capabilities in coding and data analysis.

One particularly valuable application of ChatGPT is in the writing of case reports. Case reports play a crucial role in identifying novel findings, facilitating learning, providing training opportunities, and offering concise clinical guidance. A significant number of cases remain unreported due to difficulties of practitioners to effectively communicate their findings in a clear and engaging manner. The utilization of ChatGPT addresses these challenges by enabling physicians to articulate their observations more effectively and share valuable clinical insights and generalize their findings. By leveraging the capabilities of ChatGPT, physicians can overcome barriers in case reporting, thereby enhancing knowledge dissemination and contributing to advancements in dermatology.

Although ChatGPT was not explicitly developed for research and medical applications, including dermatology, it has demonstrated surprisingly good performance in research and clinical settings, providing basic-level support. However, it is important to acknowledge that ChatGPT lacks the depth of scientific and medical knowledge required to fully comprehend disease mechanisms and treatments. Therefore, while ChatGPT and other generic tools offer fast and accurate information, they should be used with caution and the limitations of their knowledge base should be considered. It is also crucial to recognize that specialized AI tools will continue to be developed, but the general public will likely rely on generic tools, necessitating the awareness of potential limitations.

While the accuracy of ChatGPT in some domains might have been influenced by reliable sources like the public cancer databases, many other specialties in dermatology lack publicly available data sources. This highlights the need for additional resources and the importance of developing domain-specific models to cater to the specific needs of dermatology and address its unique challenges.

### Limitations

To minimize the risk of bias and ensure a comprehensive approach, this study employed rigorous literature searches from various sources, including preprint servers and multiple languages (selection bias). Limitations associated with the evidence included in this review include:

#### Study Risk of Bias

The studies selected for this review demonstrate heterogeneity in terms of their focus, scope, outcomes, methodology (eg, different scoring scales), and quality. Most of these papers do not meet all of the PRISMA criteria.

#### Peer Review

Most of the papers included in this review are preprints that have not undergone formal peer review which may impact the reliability and accuracy of the results.

#### Single-Author Review

This systematic review was conducted by a single author, which may introduce subjectivity and limit the interpretability of the results.

#### ChatGPT Version Variation

As ChatGPT technology is still evolving, some studies examined the GPT 3.6 version, while others utilized GPT-4 and fine-tuning techniques. This variation in versions and methodologies may result in a limited understanding of the technology’s potential applications.

### Comparison with Prior Work

This study presents, to the best of the authors’ knowledge, the first systematic review focused on the applications and implementation of ChatGPT specifically in the field of dermatology and related areas.

While several reviews have been published on the applications of ChatGPT in healthcare [8], this paper uses insights generated by these reviews, but goes beyond the scope of prior work. Our study not only analyzes articles that express opinions about ChatGPT, supported by qualitative or quantitative evidence, but also includes papers that have actively utilized ChatGPT in their research and have explicitly mentioned it in acknowledgments, methods, and supplementary materials. By taking this comprehensive approach, the review identifies key themes and trends observed in the latest publications, showcasing the diverse range of fields that have benefited from the integration of ChatGPT.

### Conclusions

This review demonstrates a shift towards evidence-based literature and an increase in projects utilizing ChatGPT in dermatology-related areas, indicating a growing interest in its practical applications. The use of ChatGPT for case reports, starting with practitioners inputting anonymized electronic health records to get a first draft, showcases a potential paradigm shift in the field. ChatGPT can serve as a virtual mentor, offering interactive interactions, and facilitating the acquisition of essential medical knowledge and skills. The potential impact of language models on administrative personnel’s work should be also recognized, as they offer various benefits such as clinical decision support, text summarization, efficient writing, and multilingual communication.

LLMs have the potential to revolutionize collaboration and communication among researchers from diverse fields. Even though specialized LLMs are being developed and will be available in various fields, including dermatology, the public will be still using freely-accessible tools like ChatGPT. To optimize effectiveness of publicly available general intelligence tools in dermatology, future efforts should prioritize improving the quality of training datasets and incorporate feedback of dermatologists in its training.

## Data Availability

Data produced in the present work are contained in the manuscript (Appendix 1). Additional data are available upon reasonable request to the author.

## Acknowledgements

The author thanks ChatGPT and Bard for assistance.

## Data Availability Statement

All papers selected for final analysis are available in appendix

## Abbreviations

AI: Artificial Intelligence
FoR: Field of Research
GAN: generative adversarial networks
GPT: generative pre-trained transformer
LLM: Large Language Model
PRISMA: Preferred Reporting Items for Systematic Reviews and Meta-Analyses

## Appendix 1

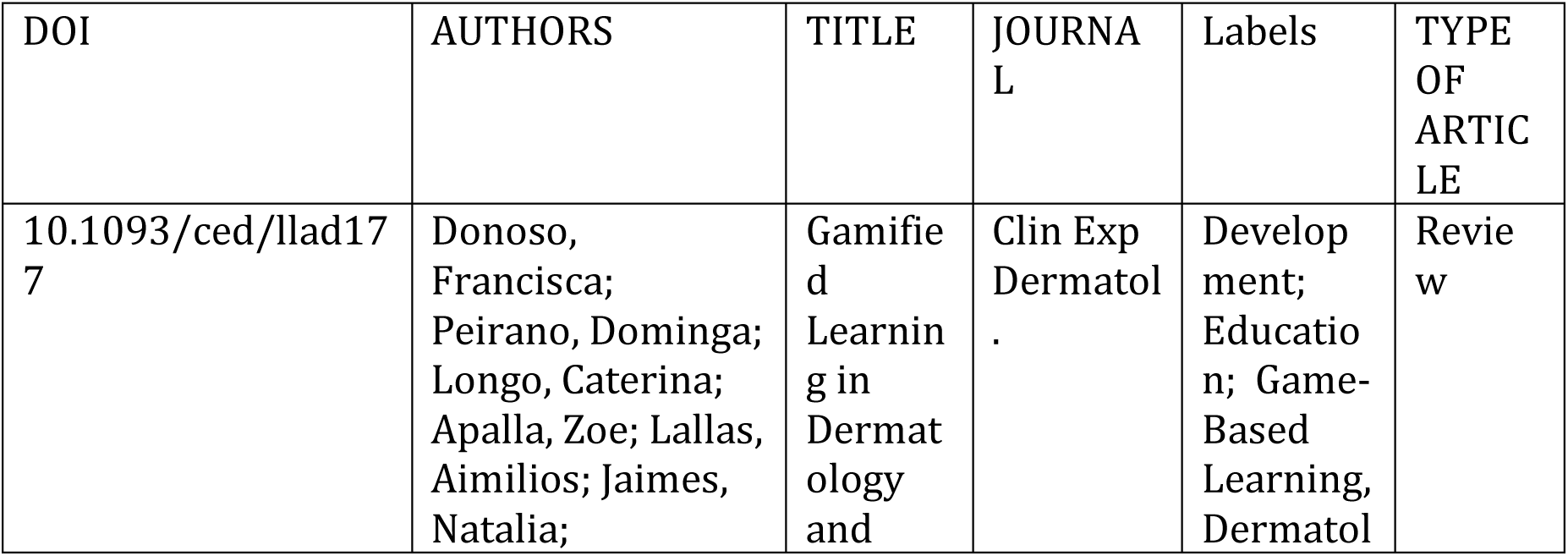
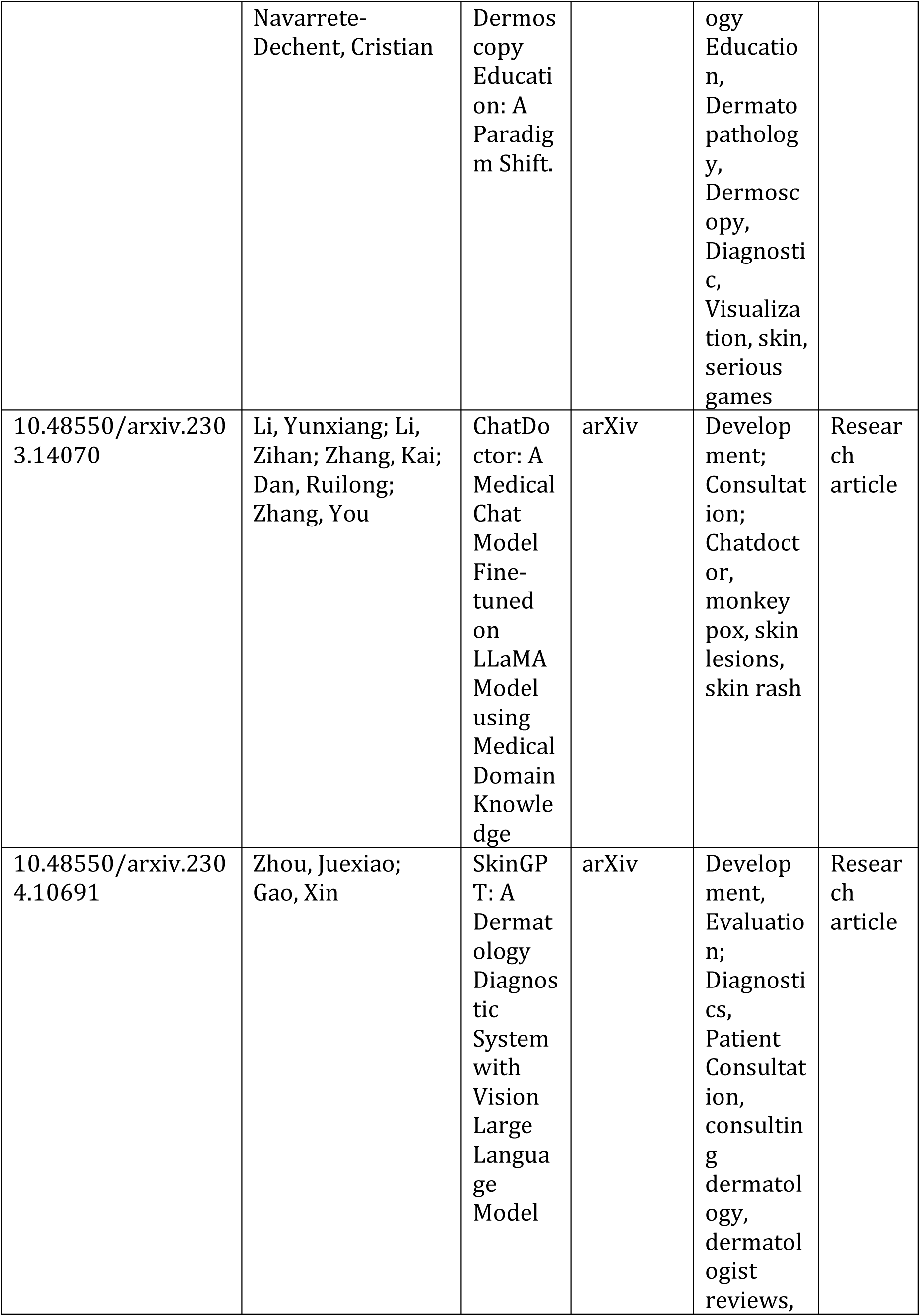
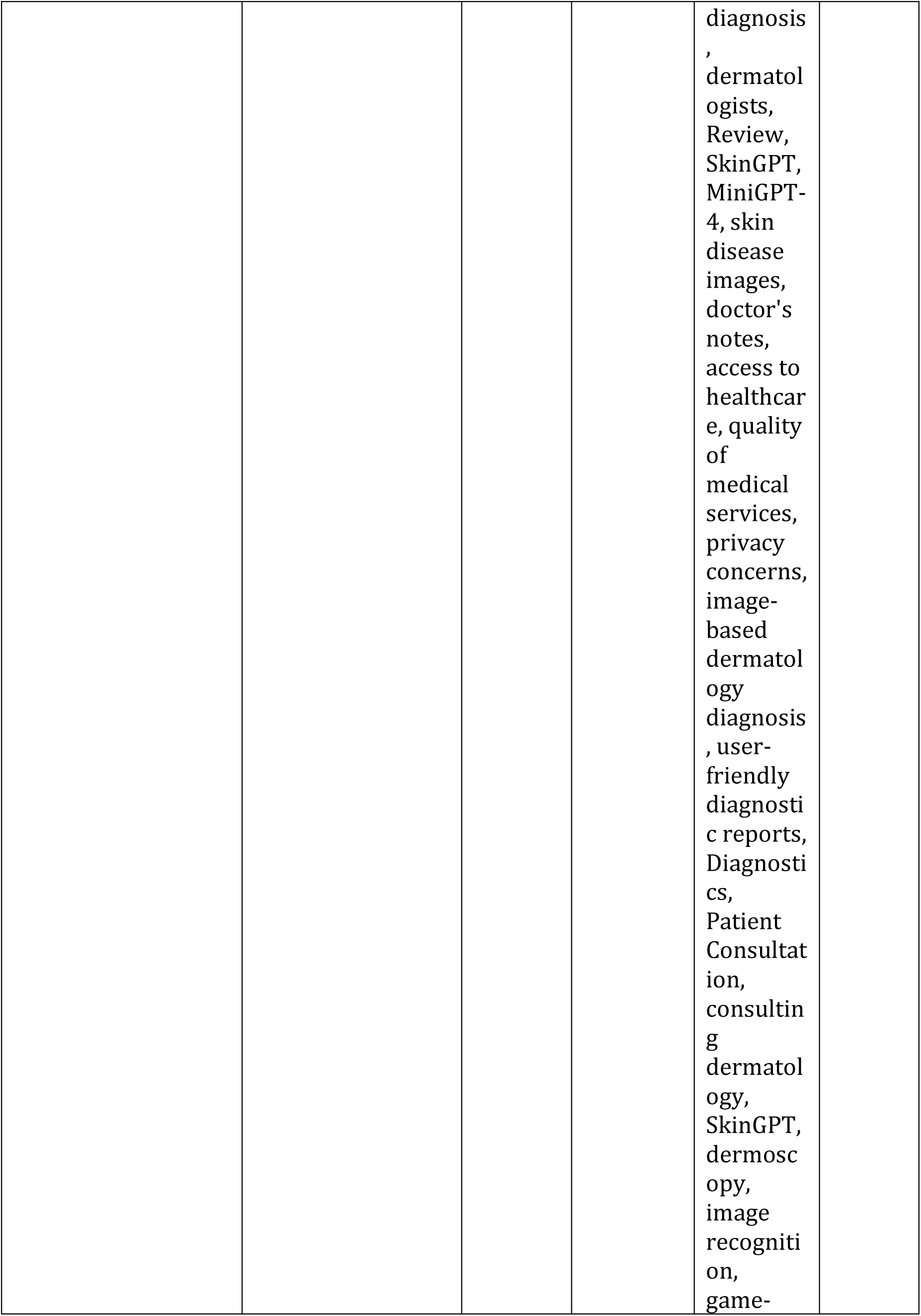
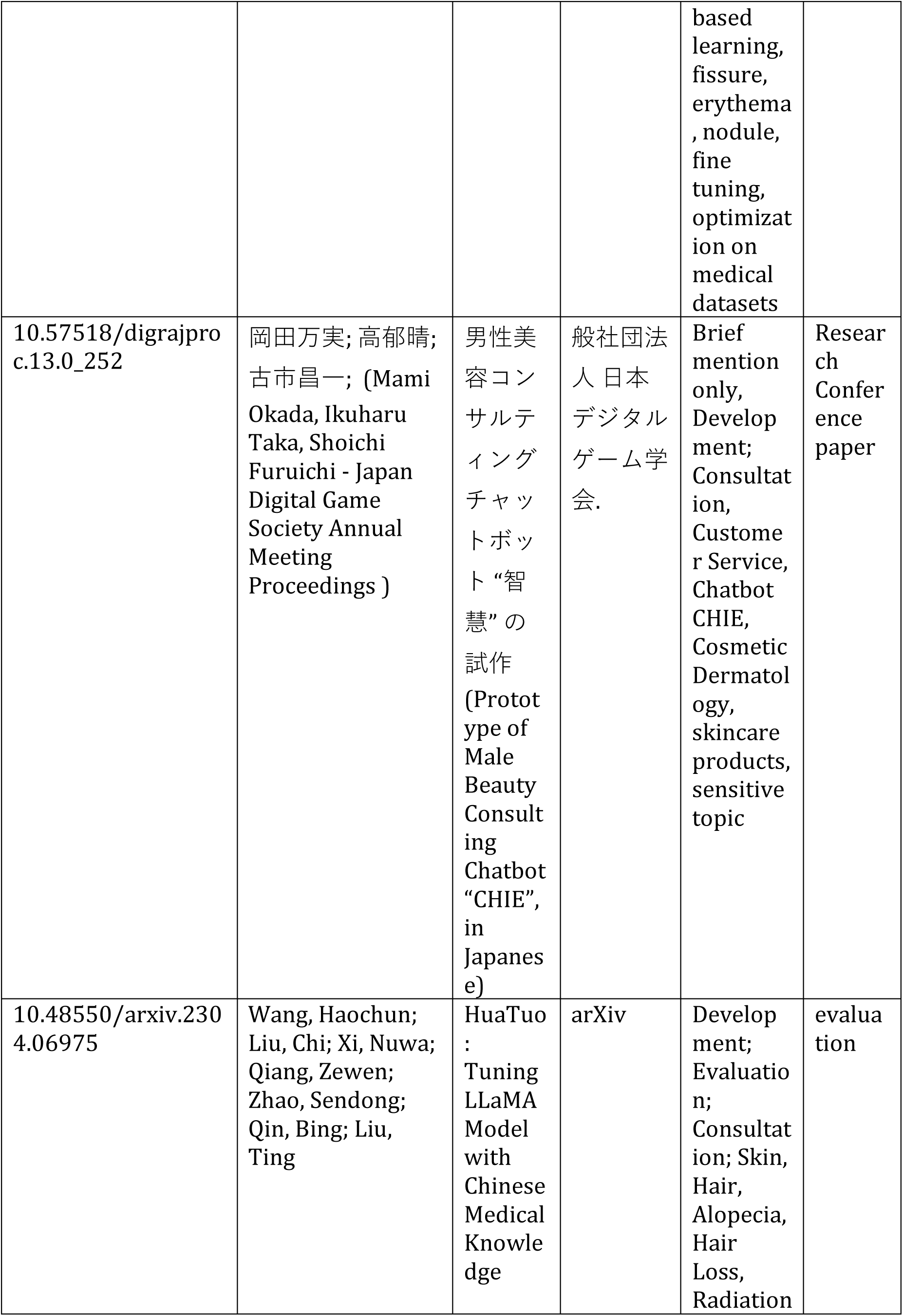
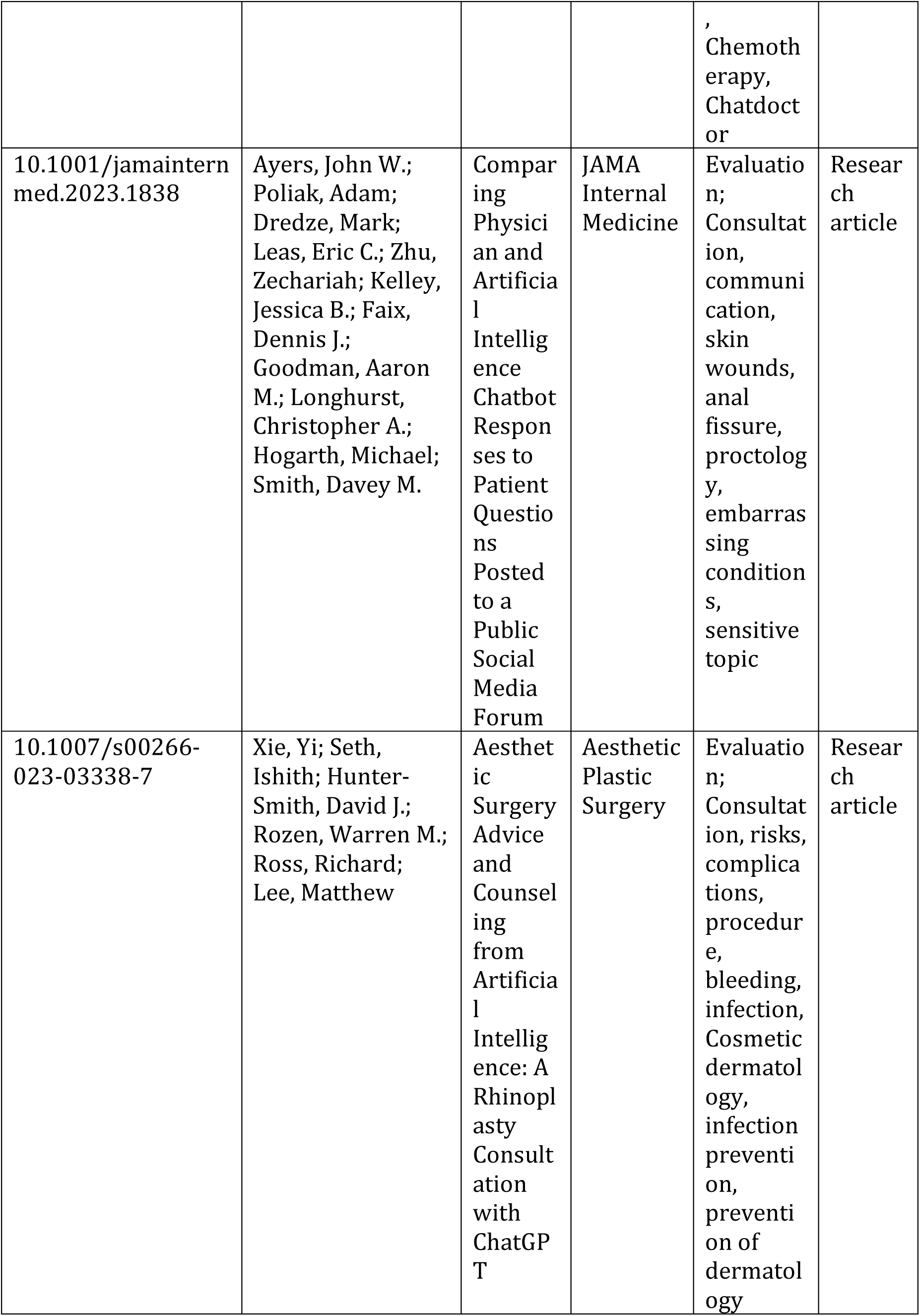
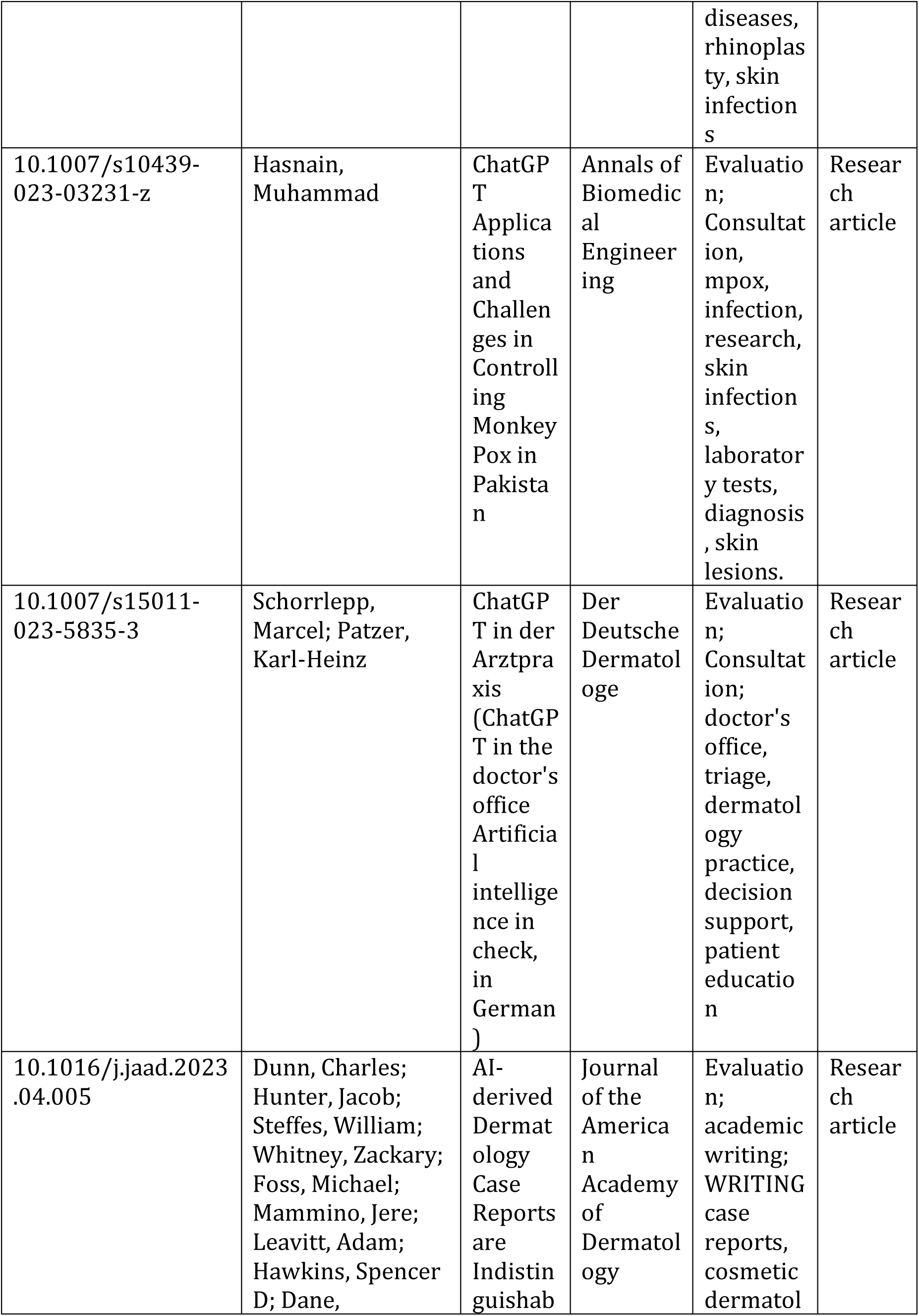
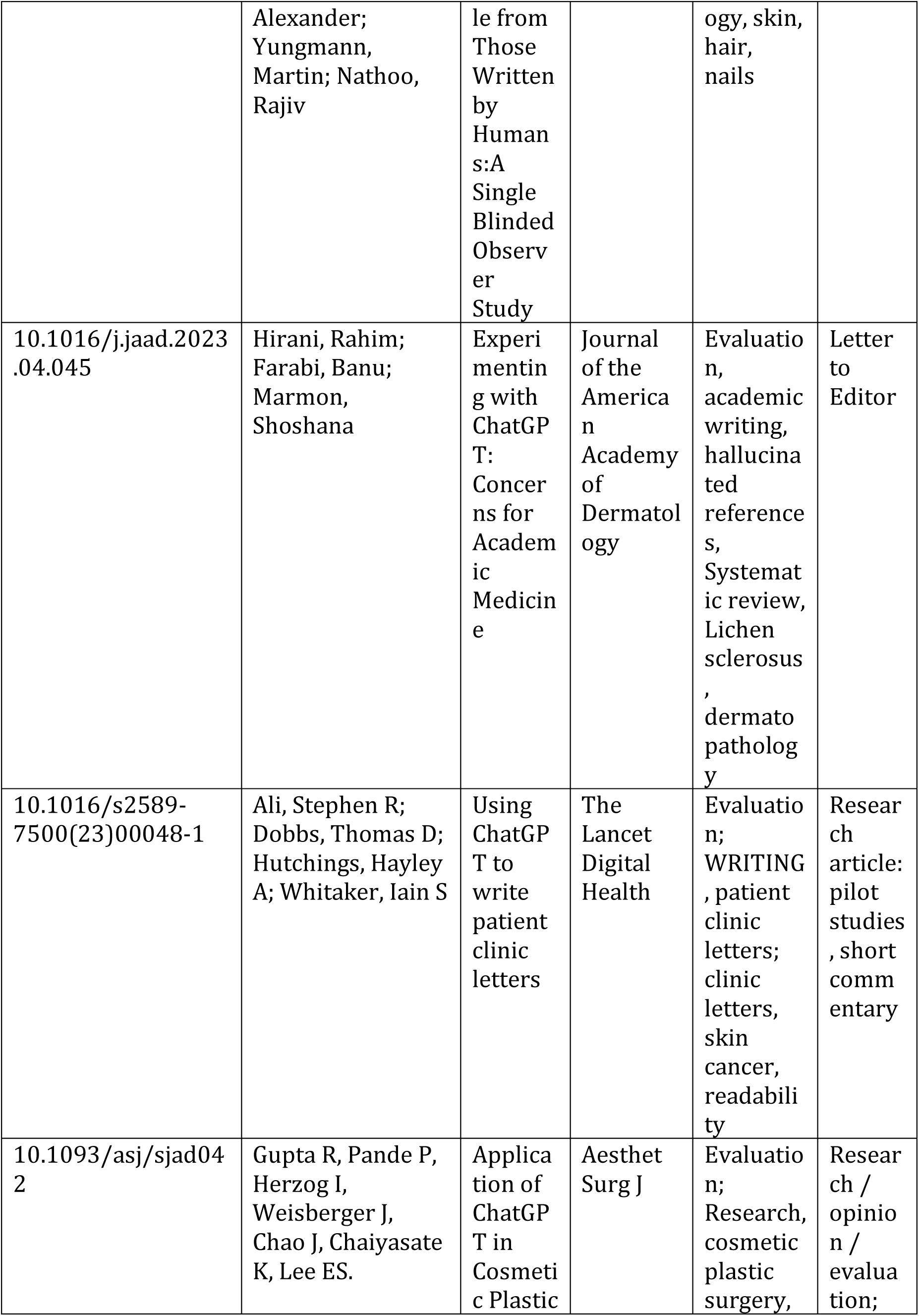
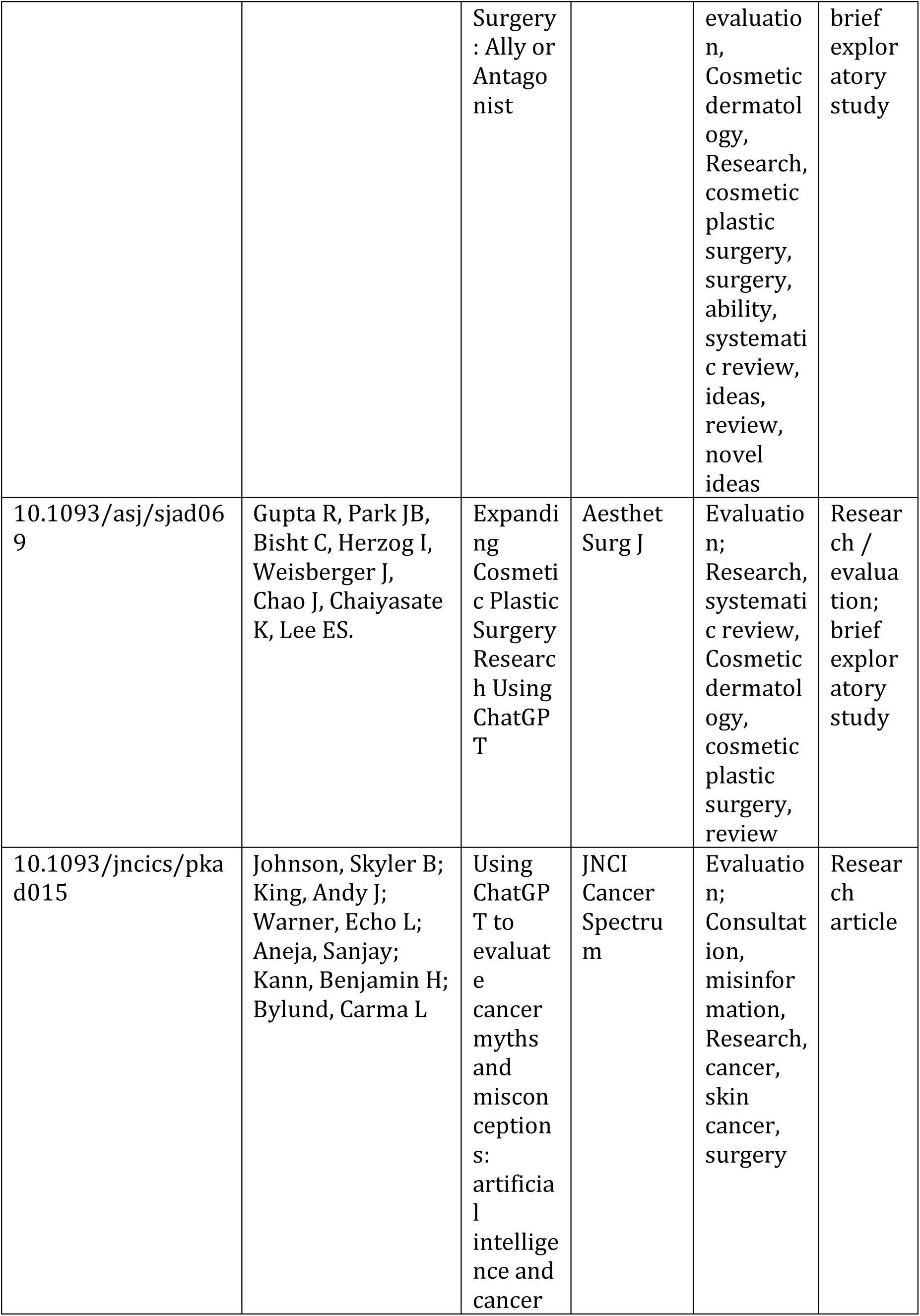
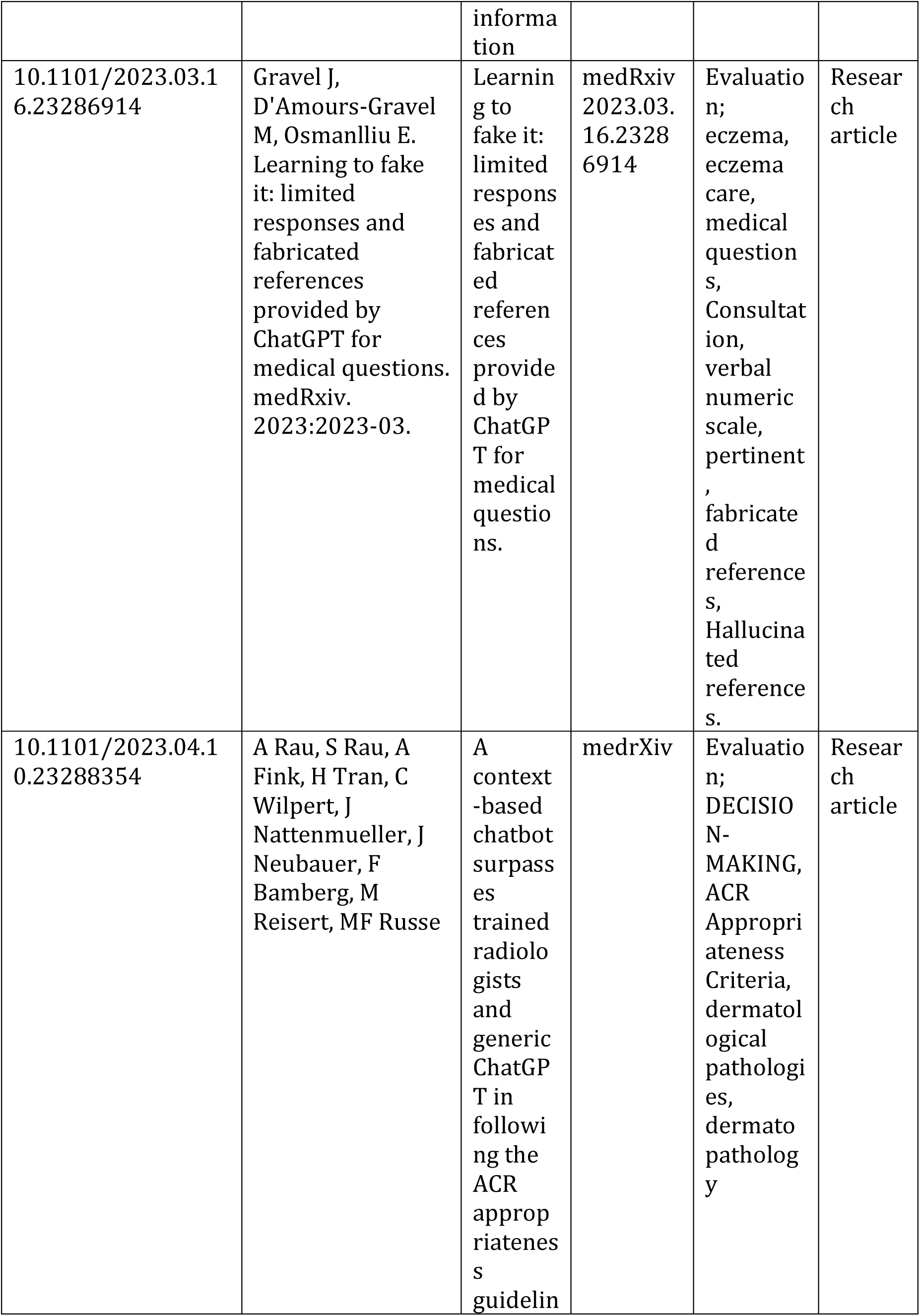
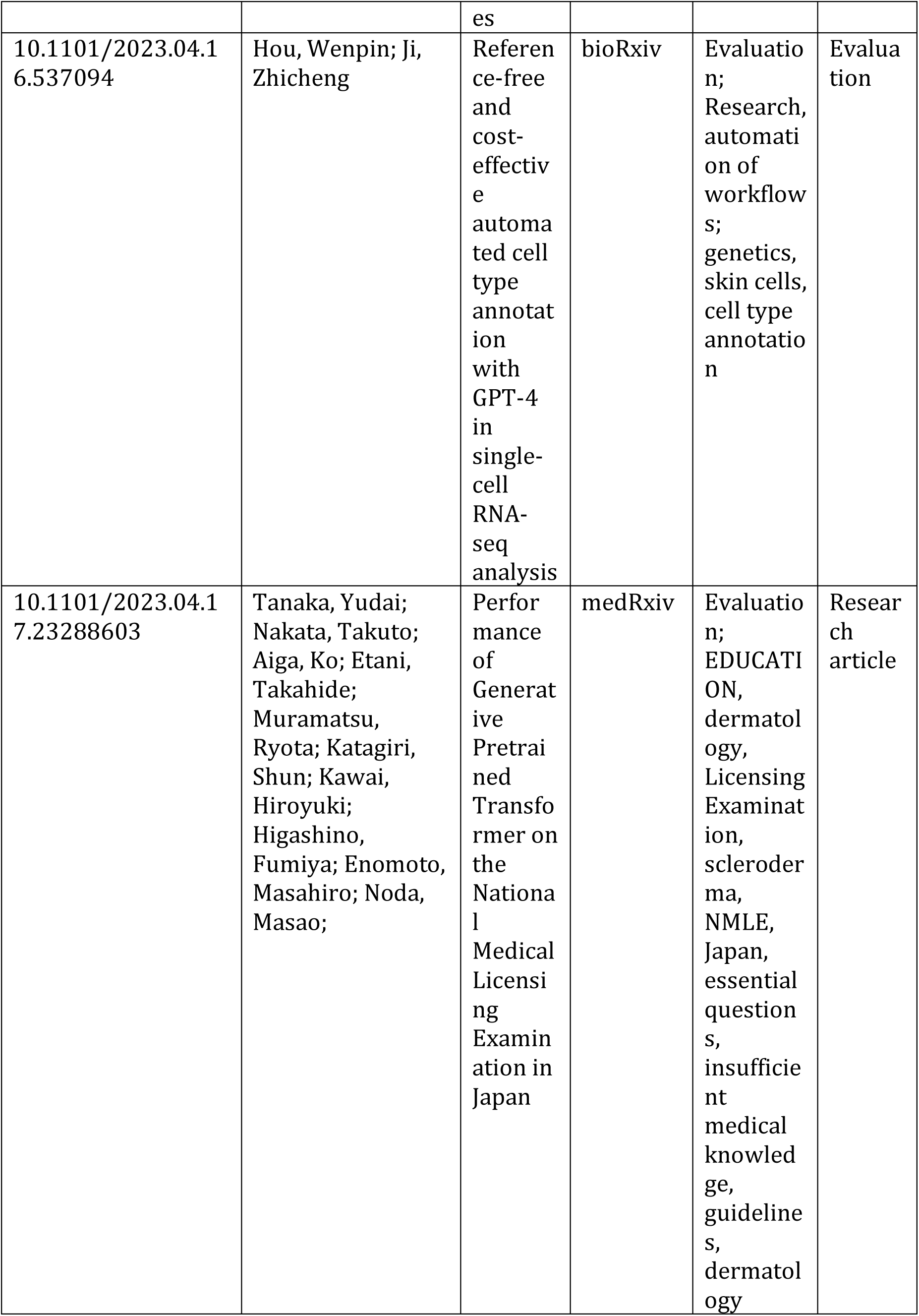
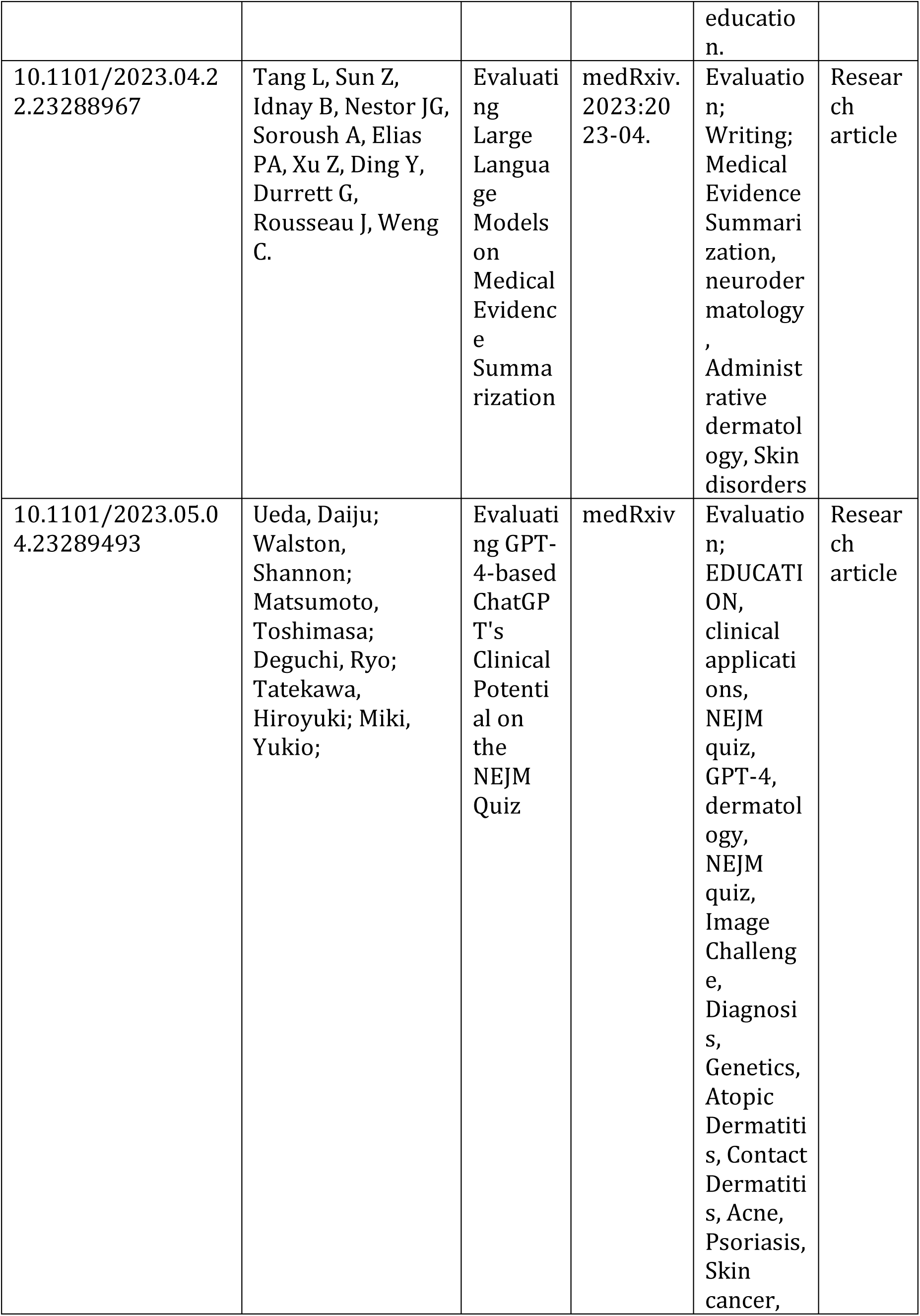
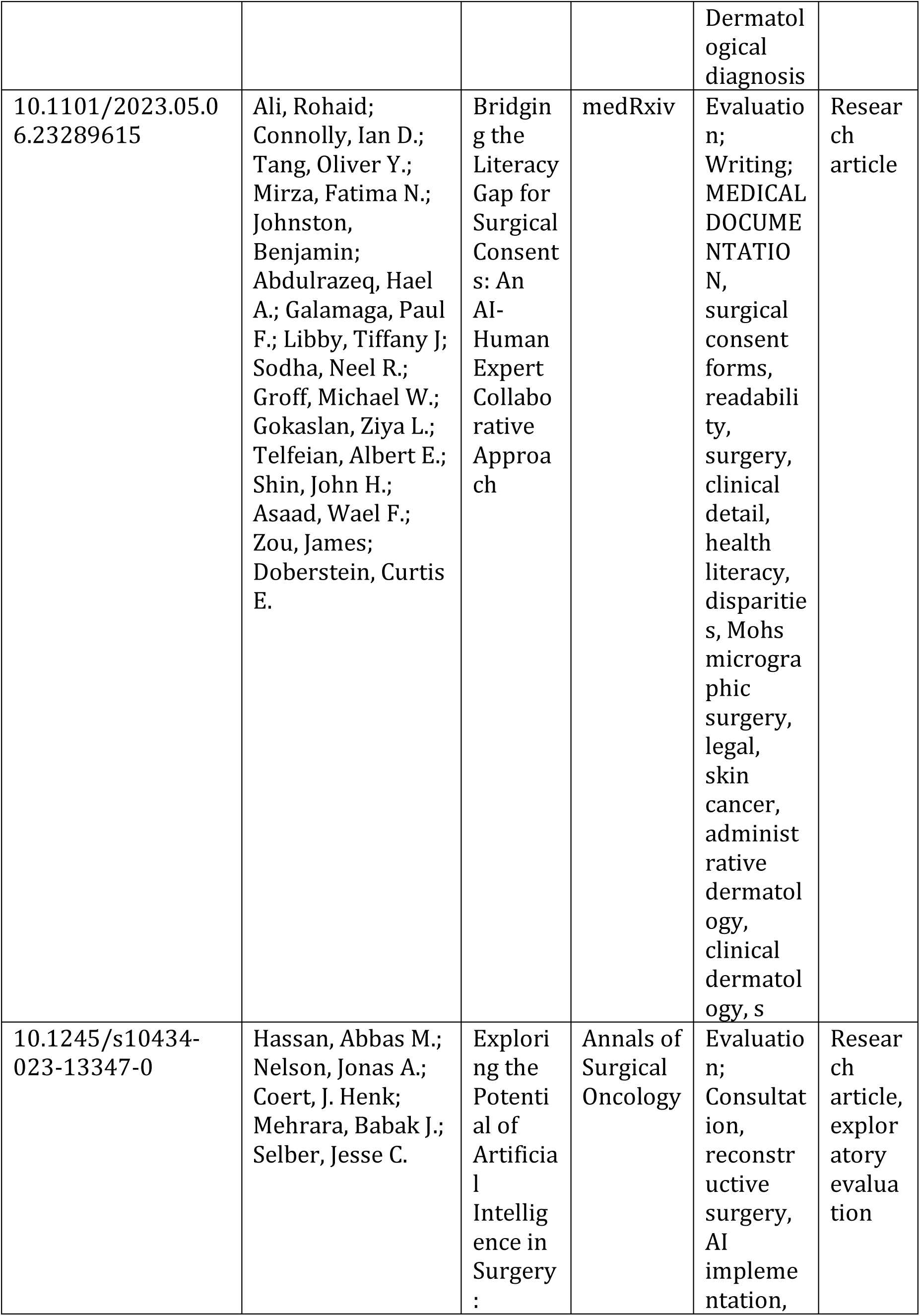
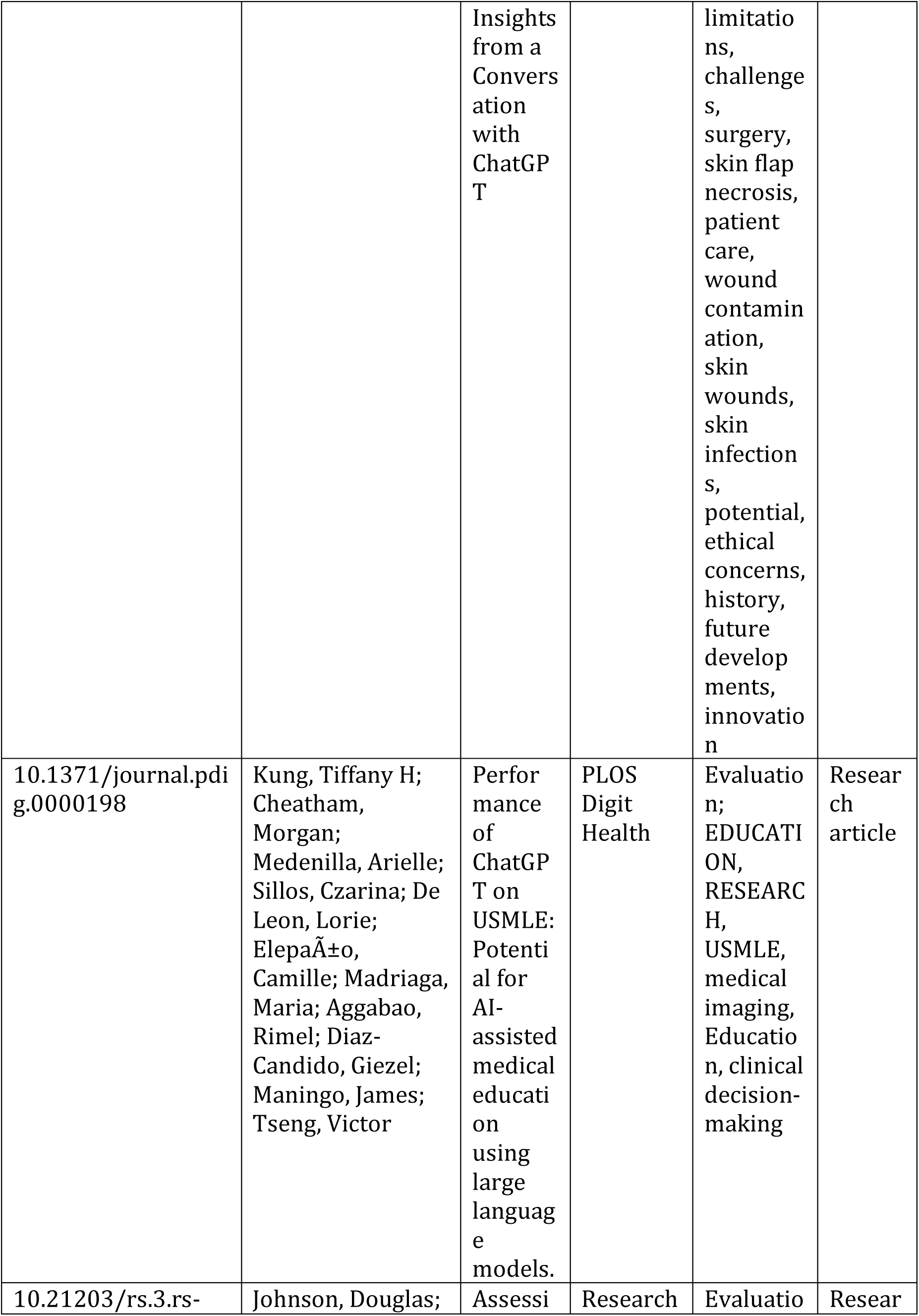
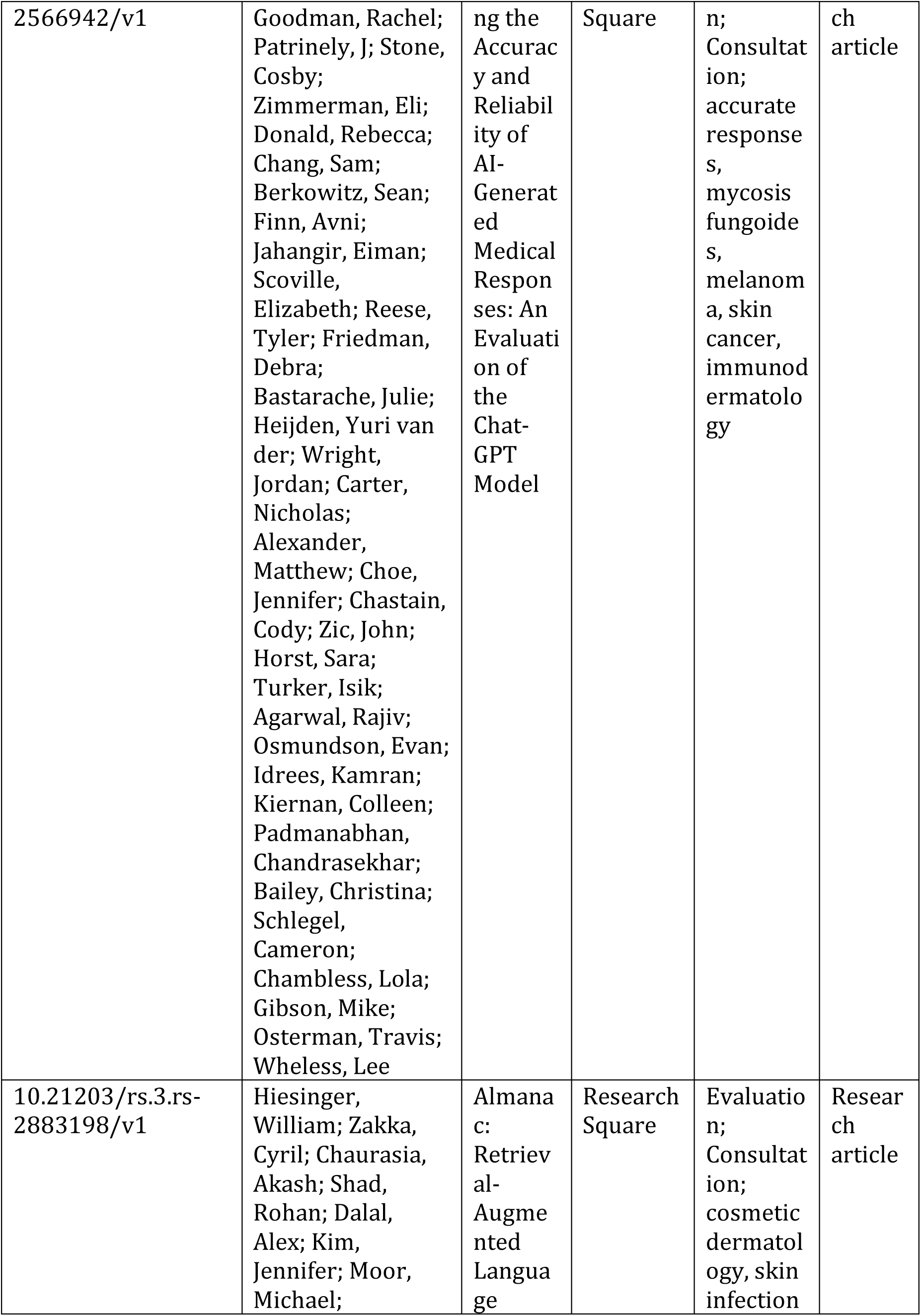
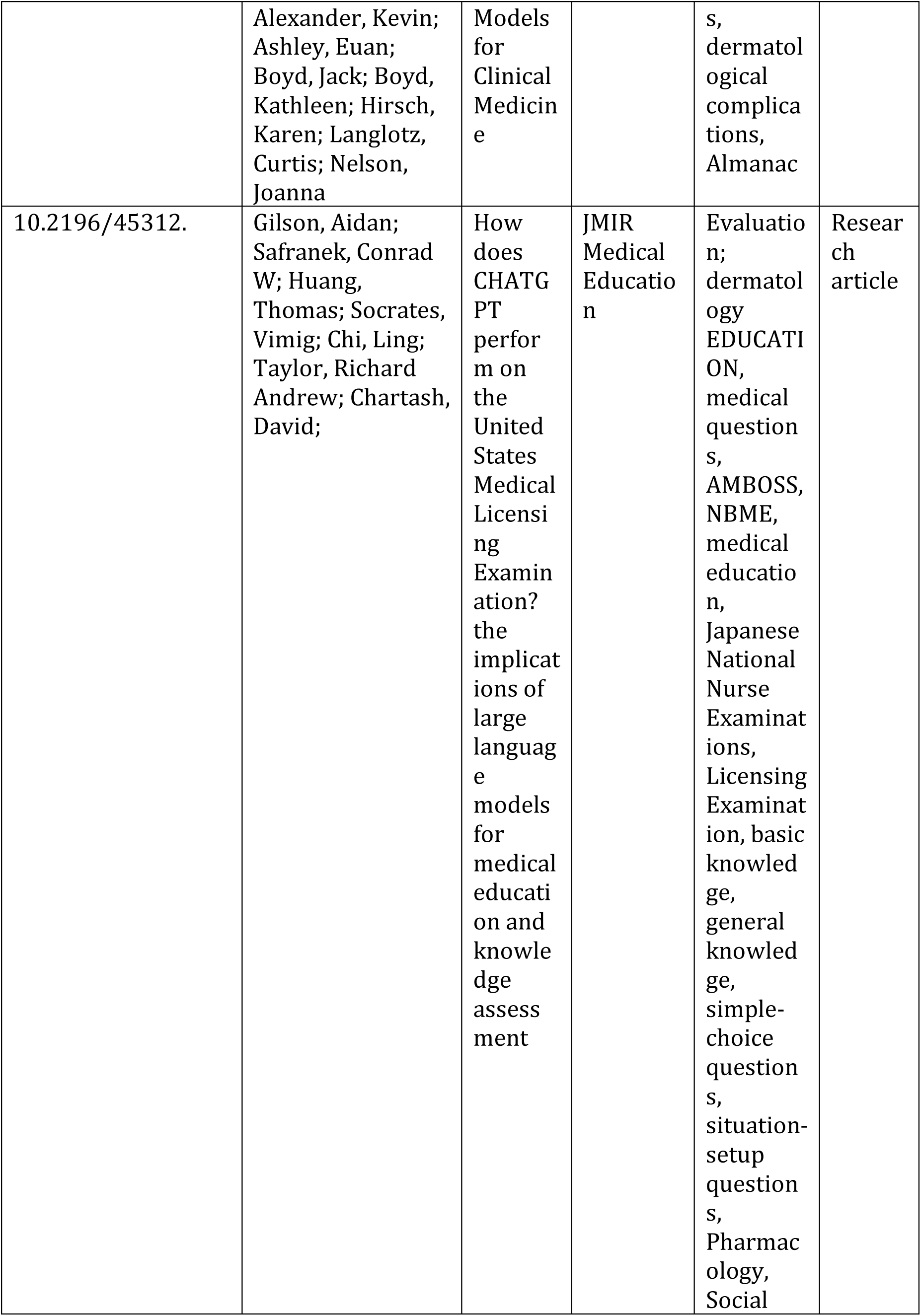
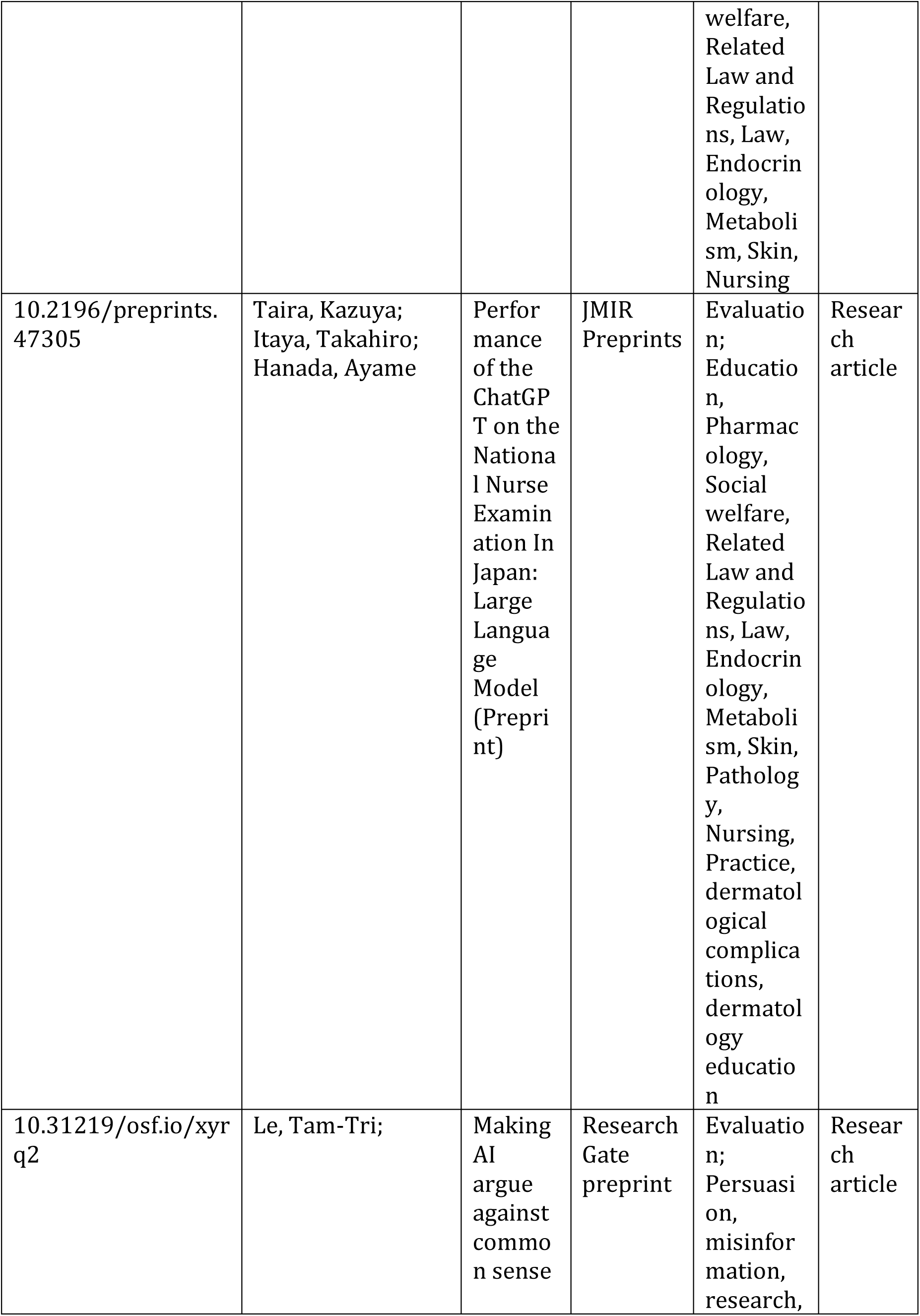
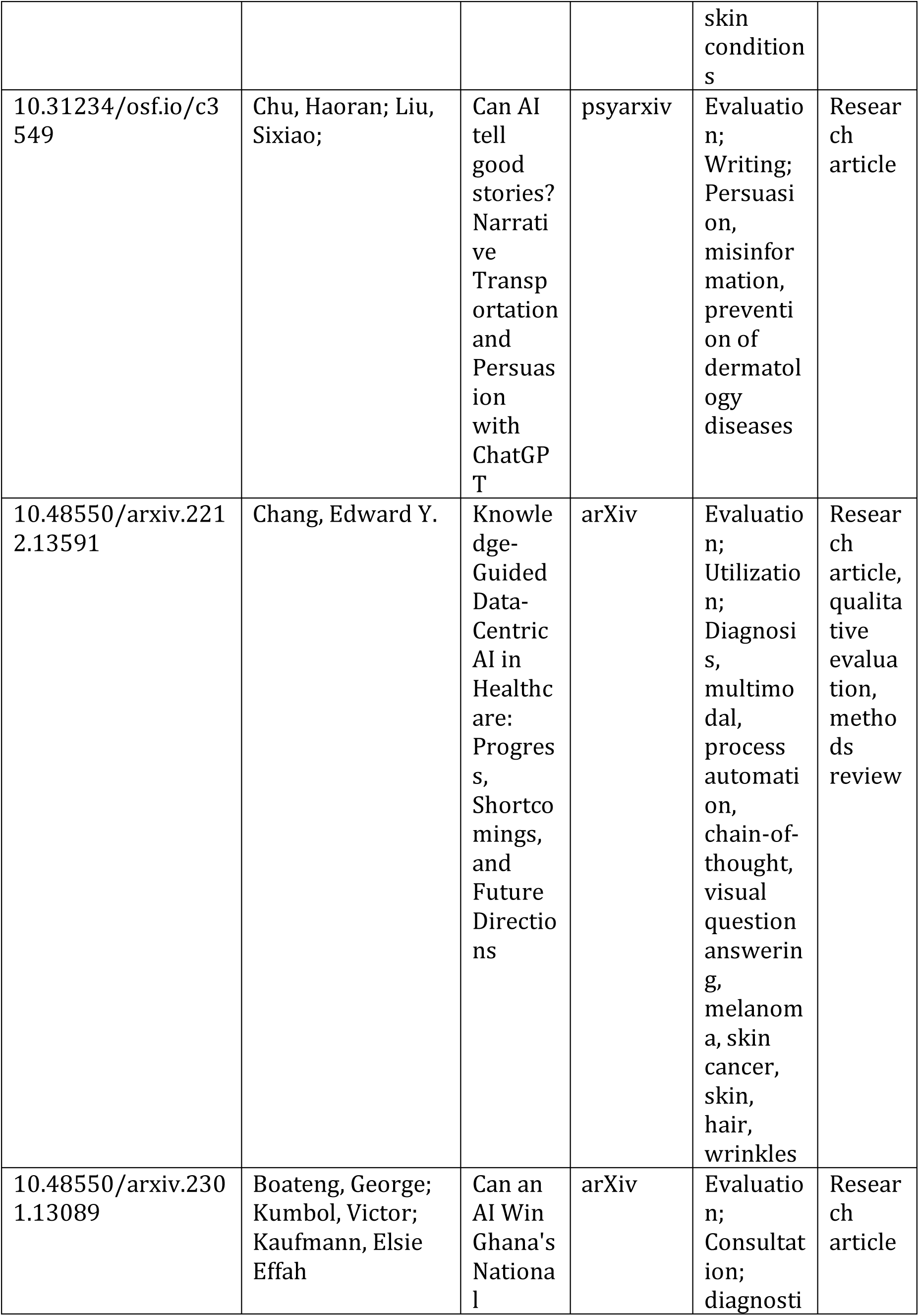
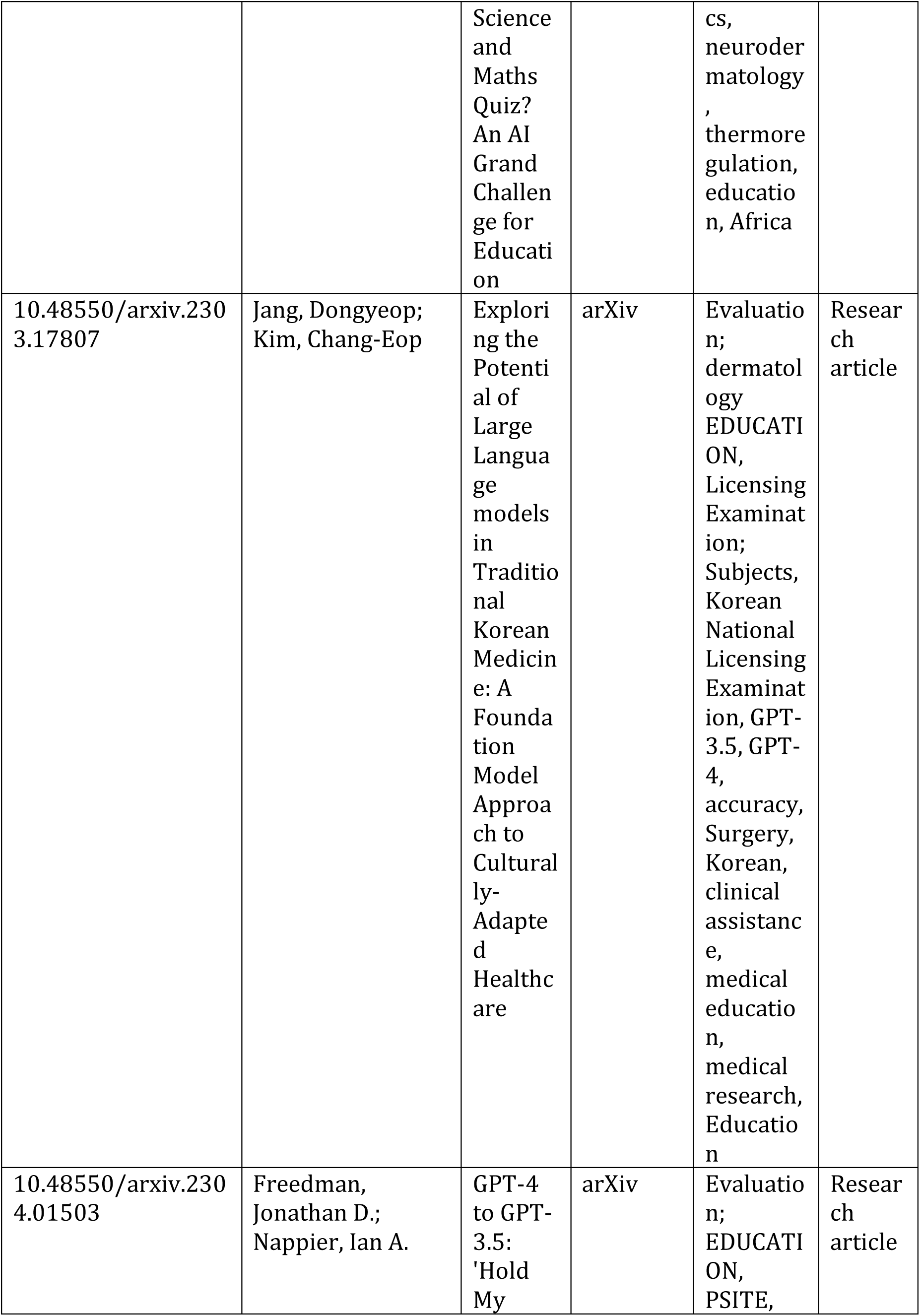
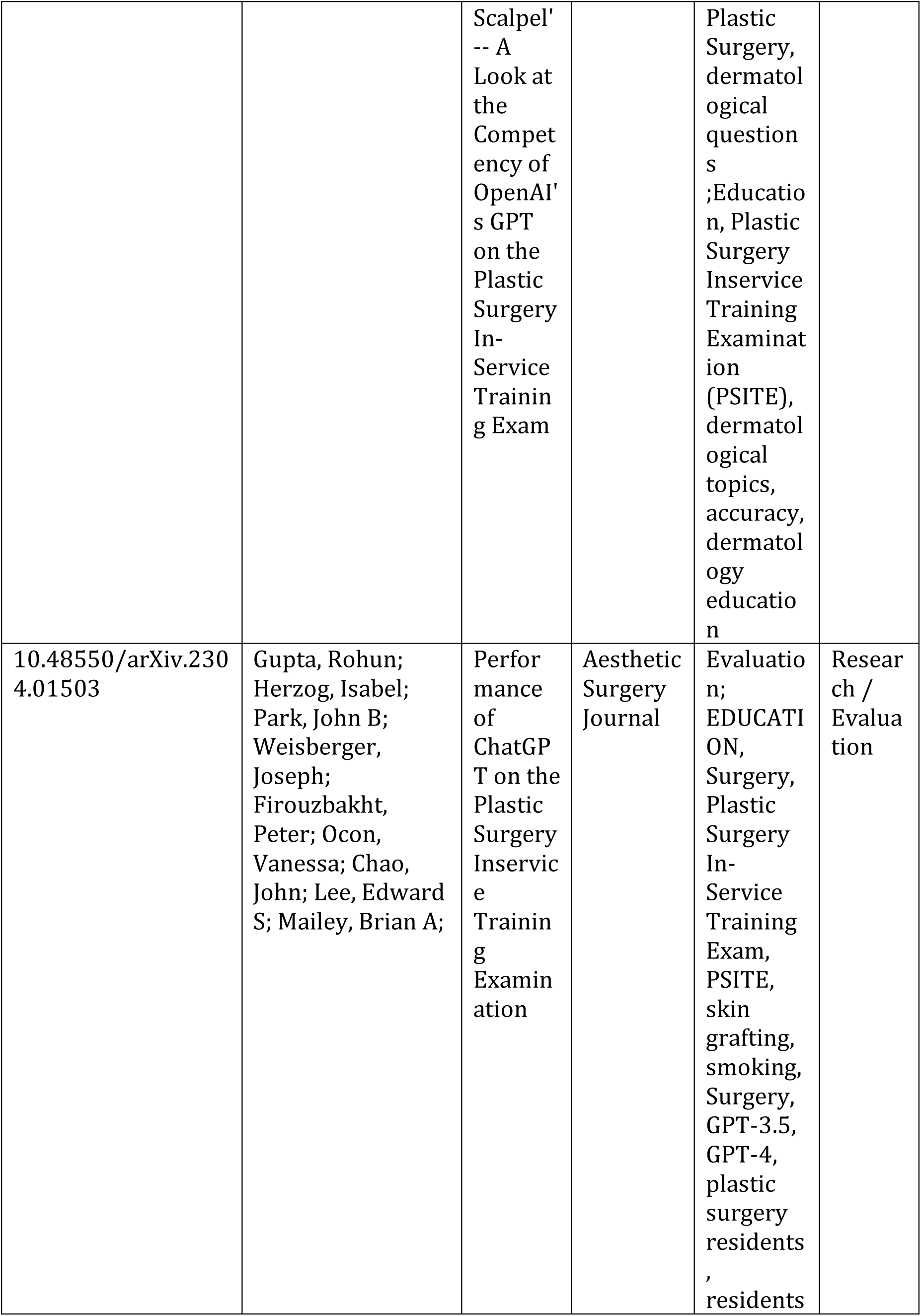
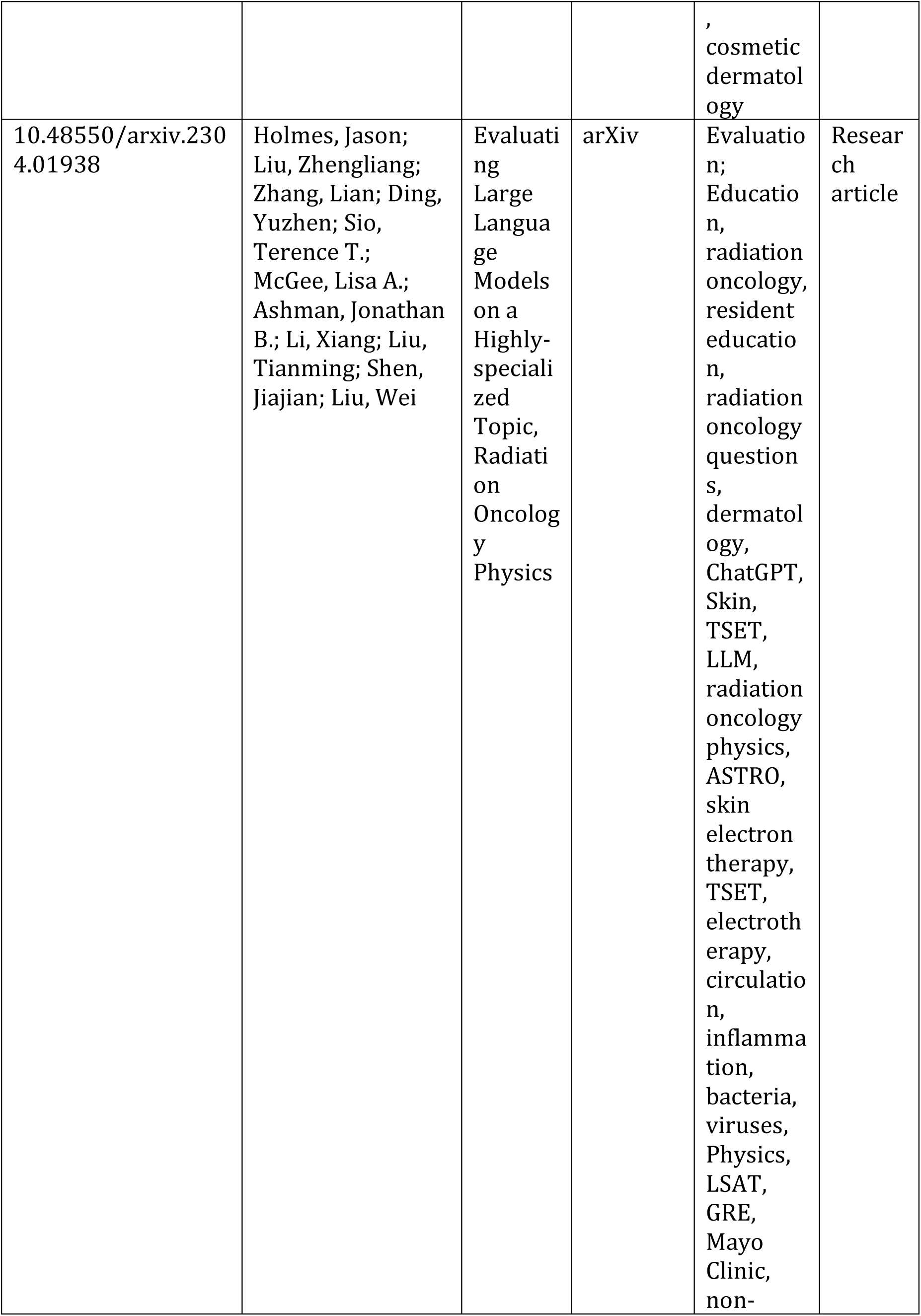
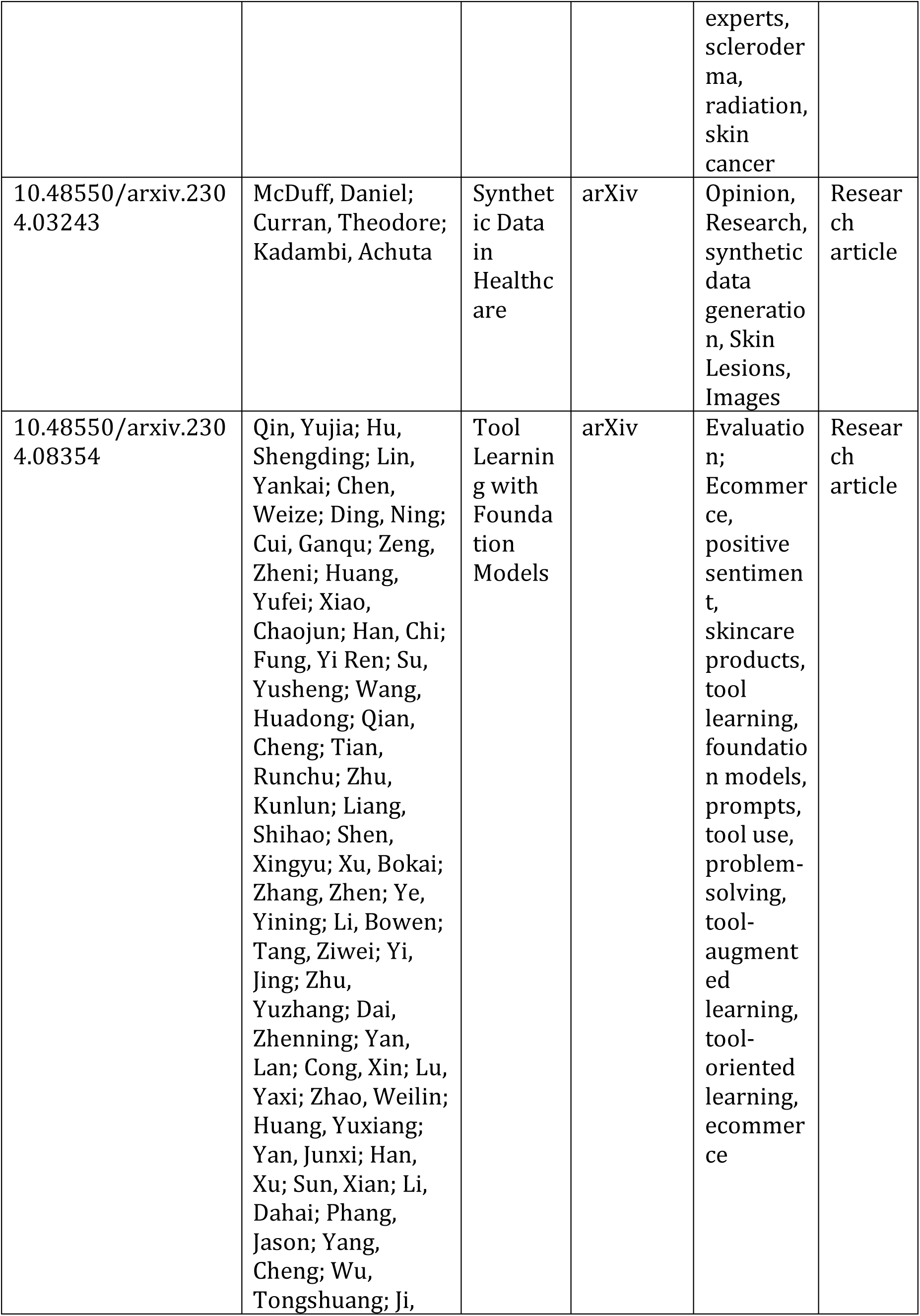
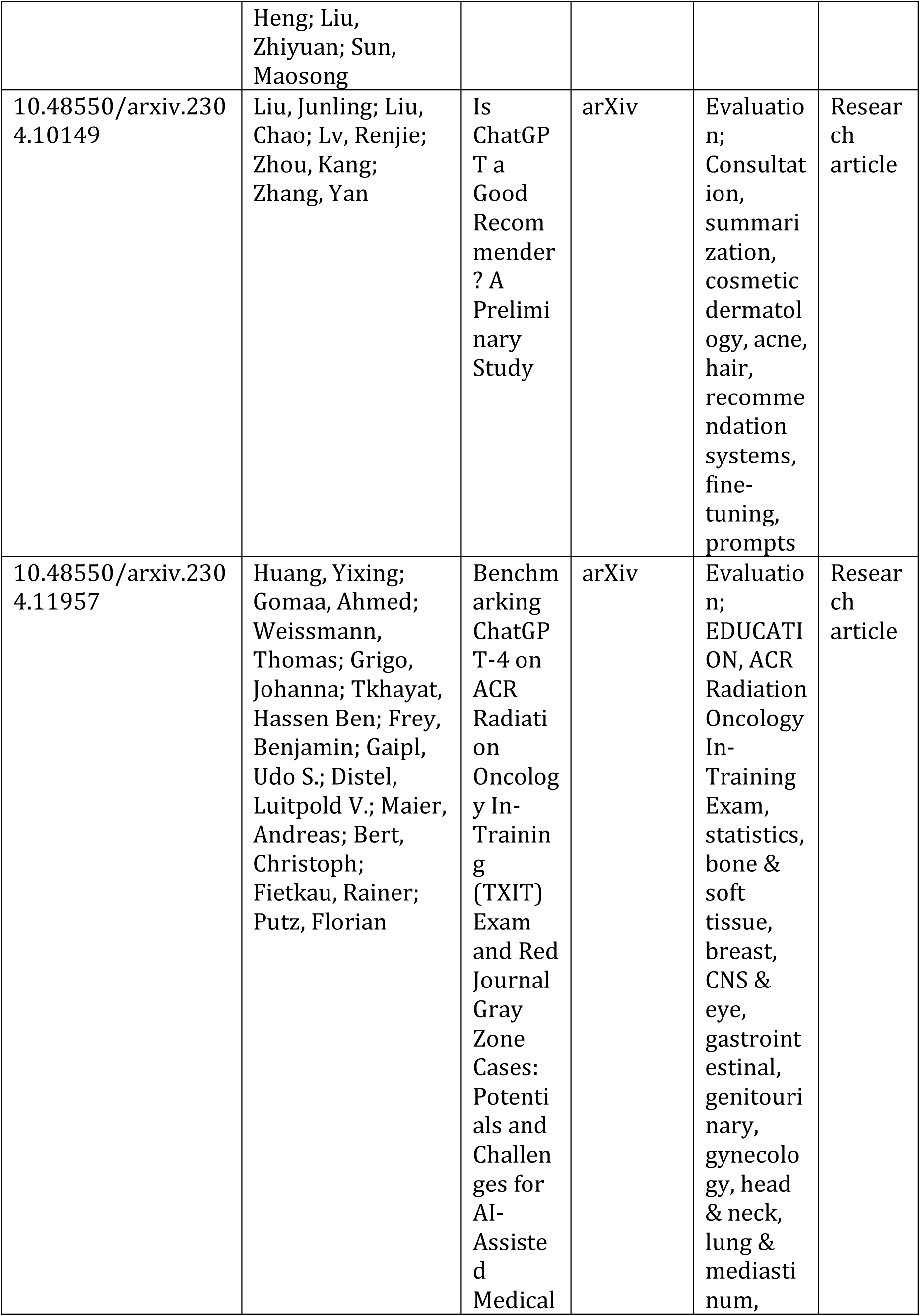
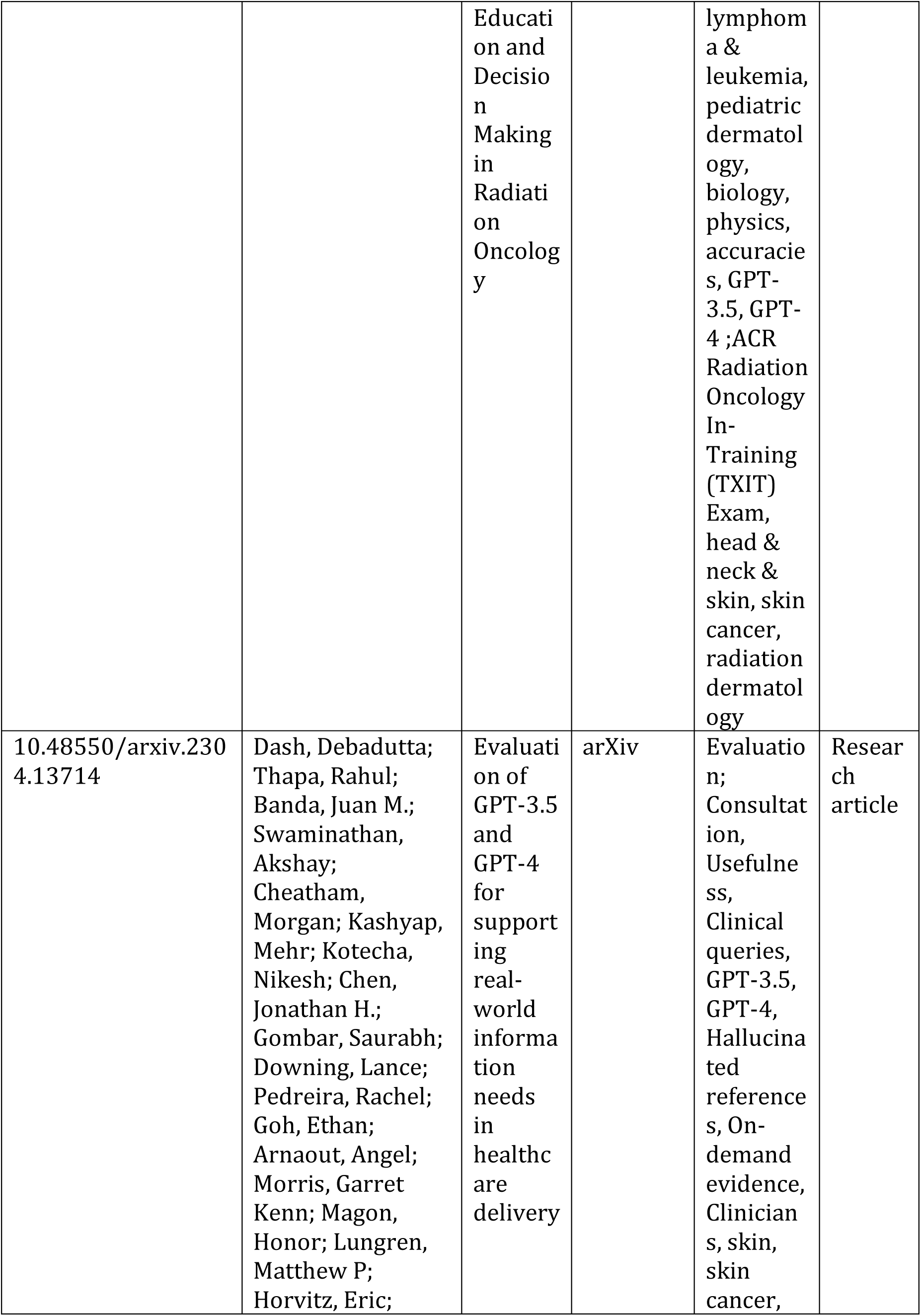
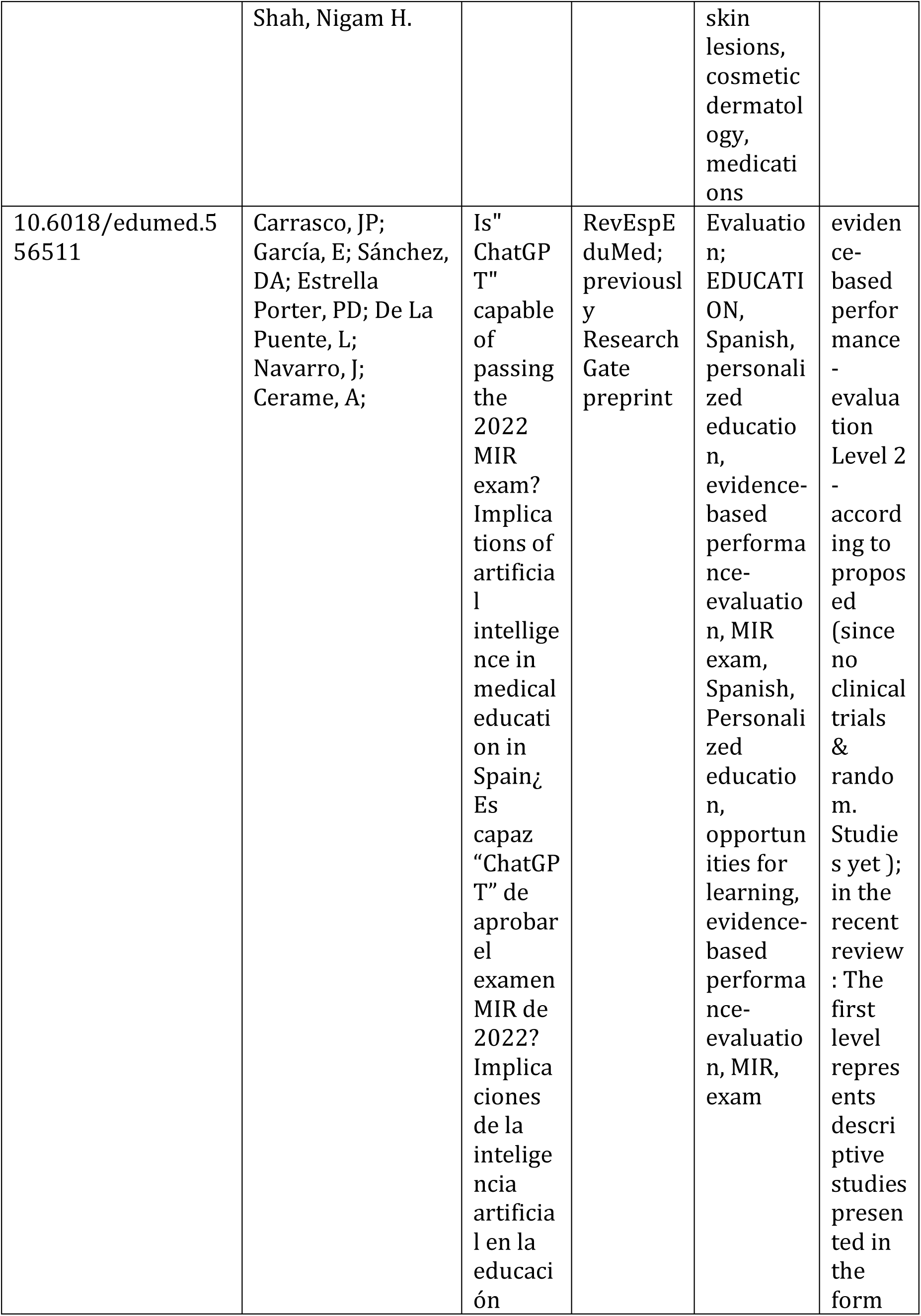
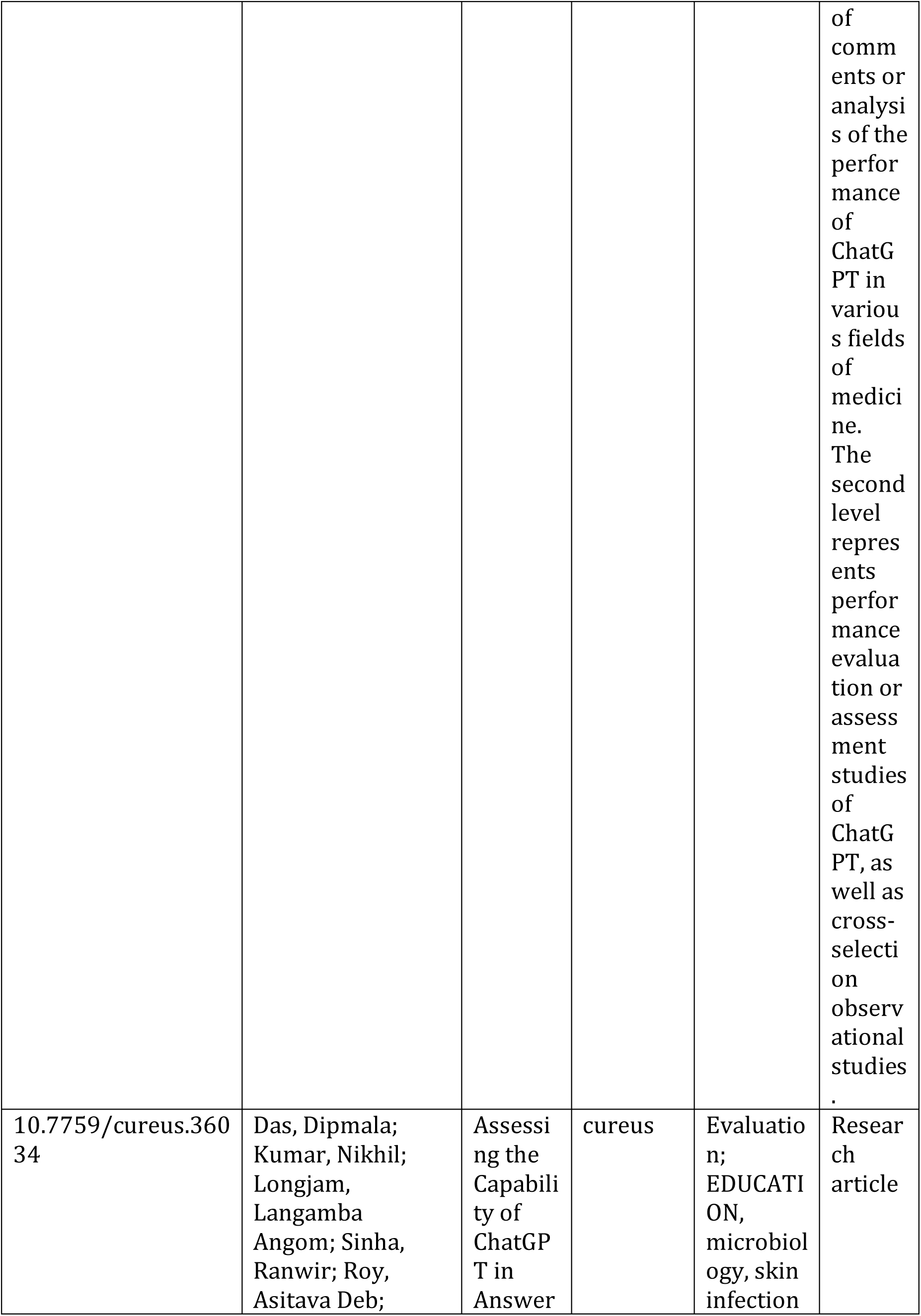
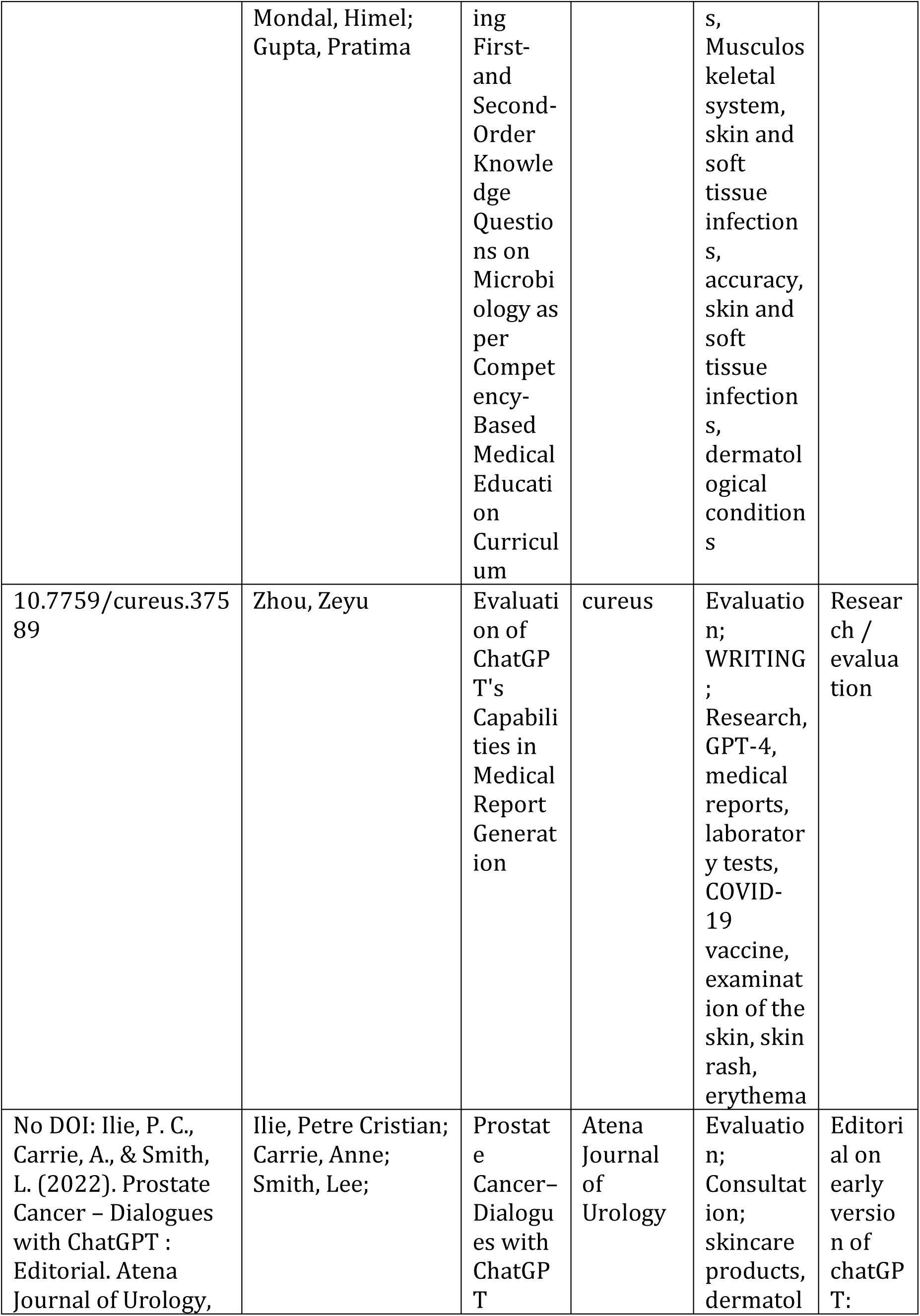
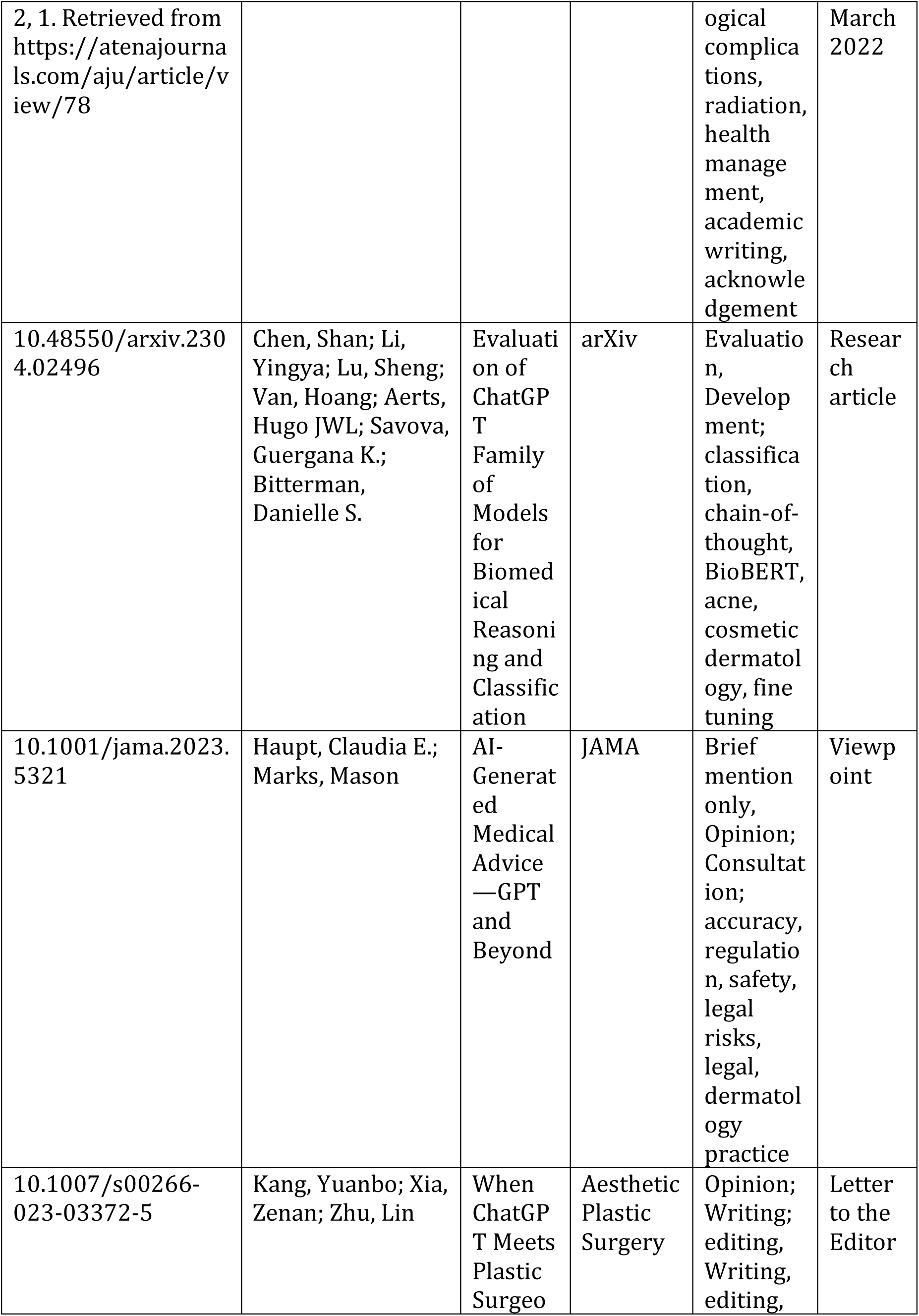
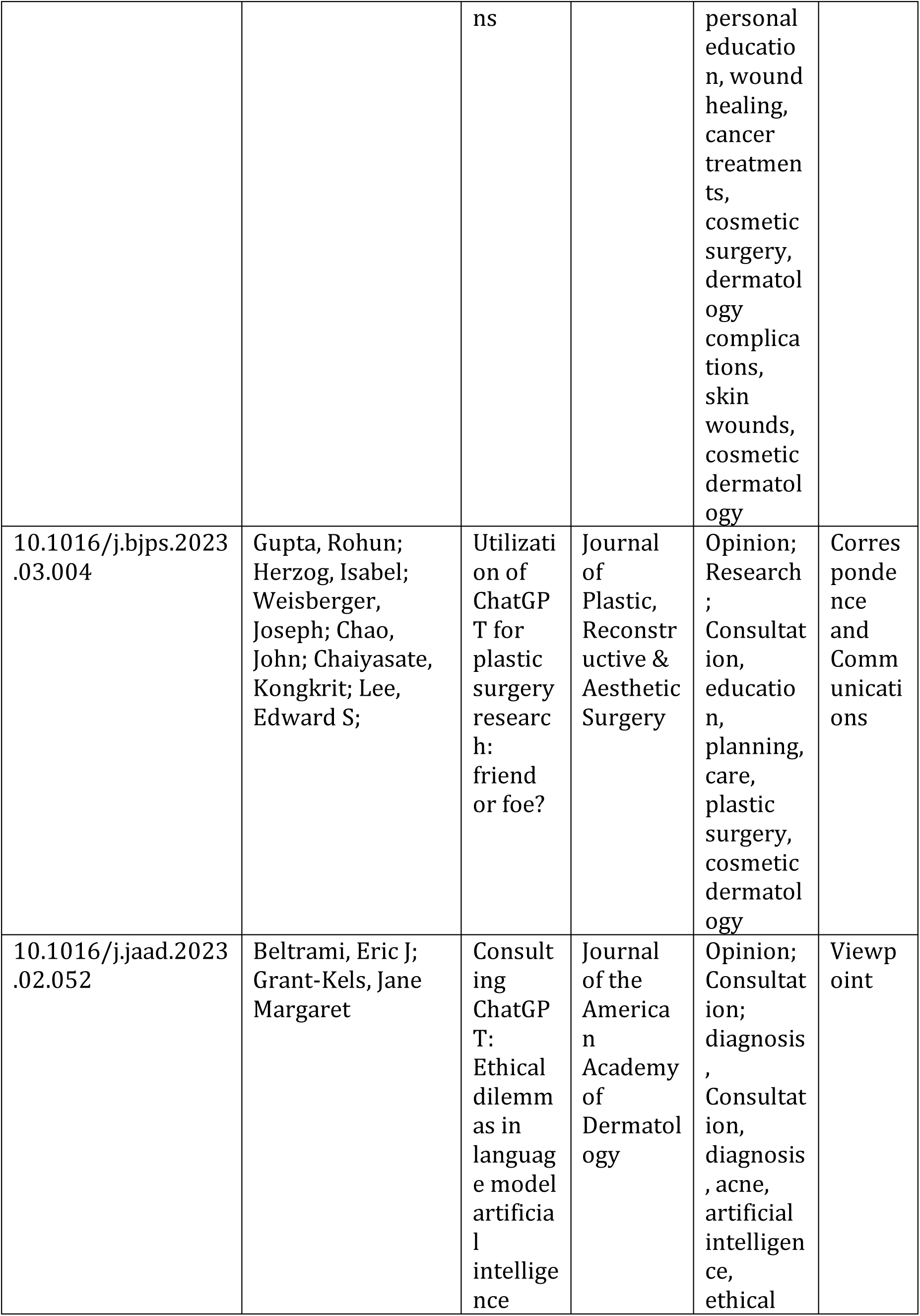
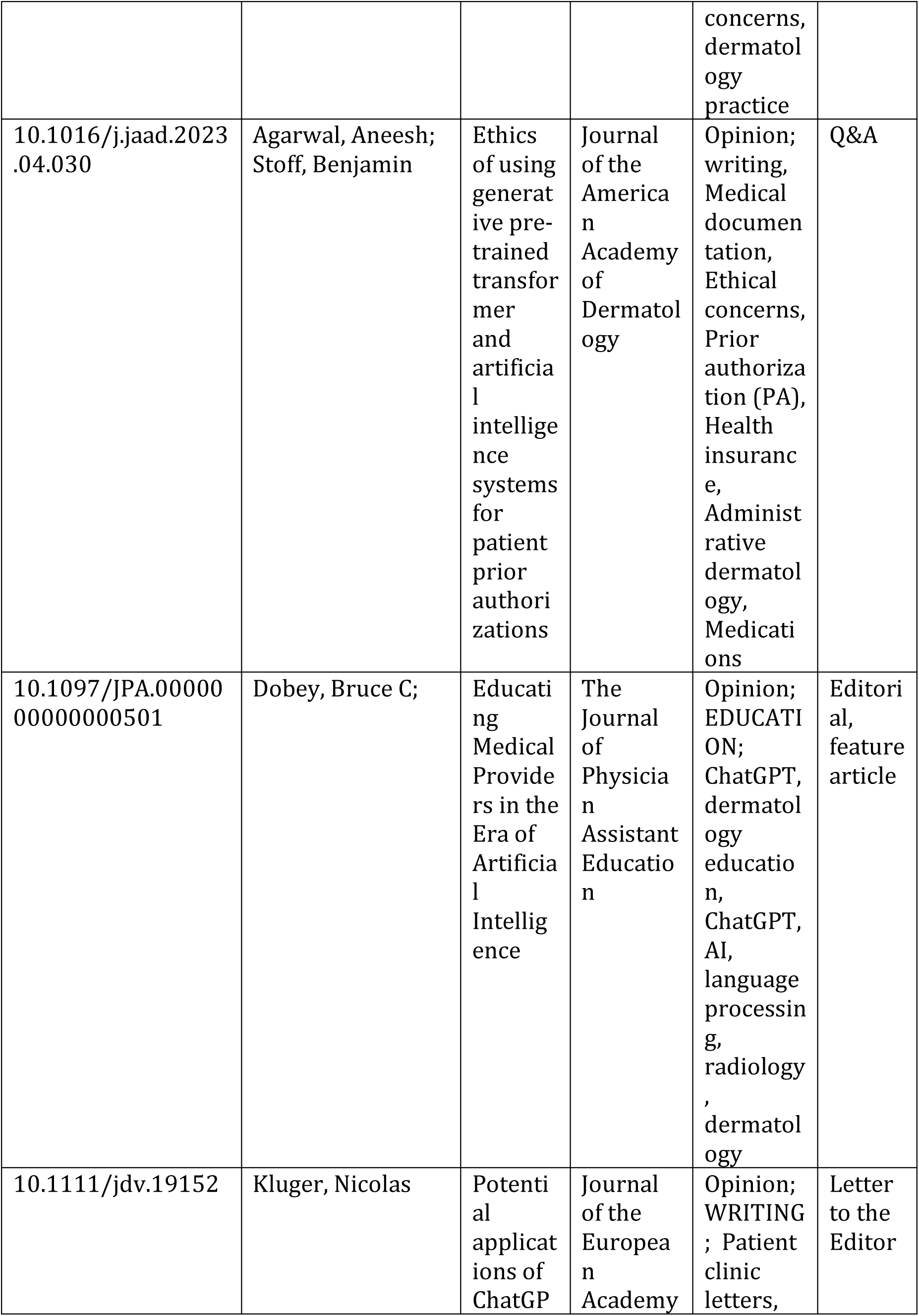
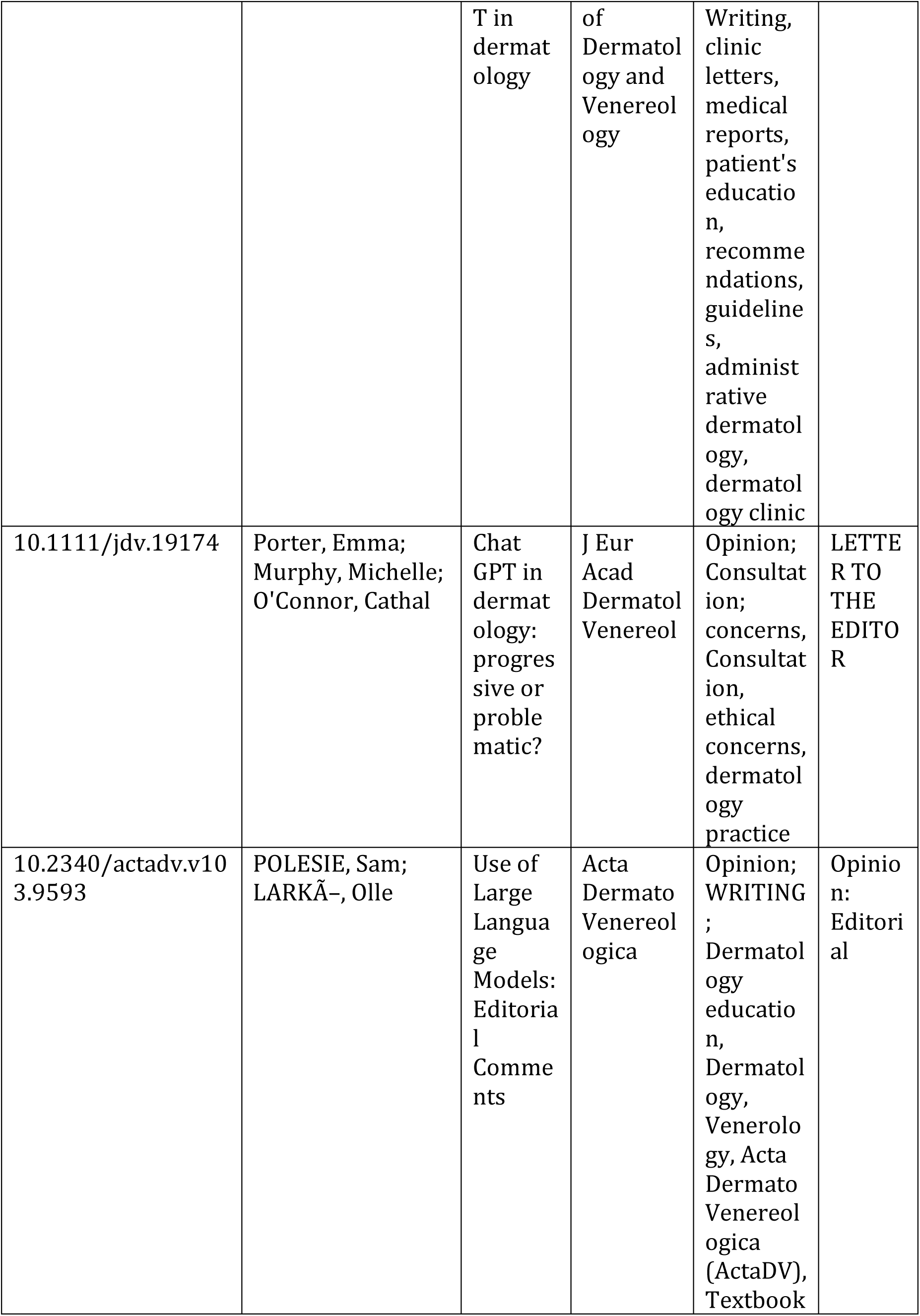
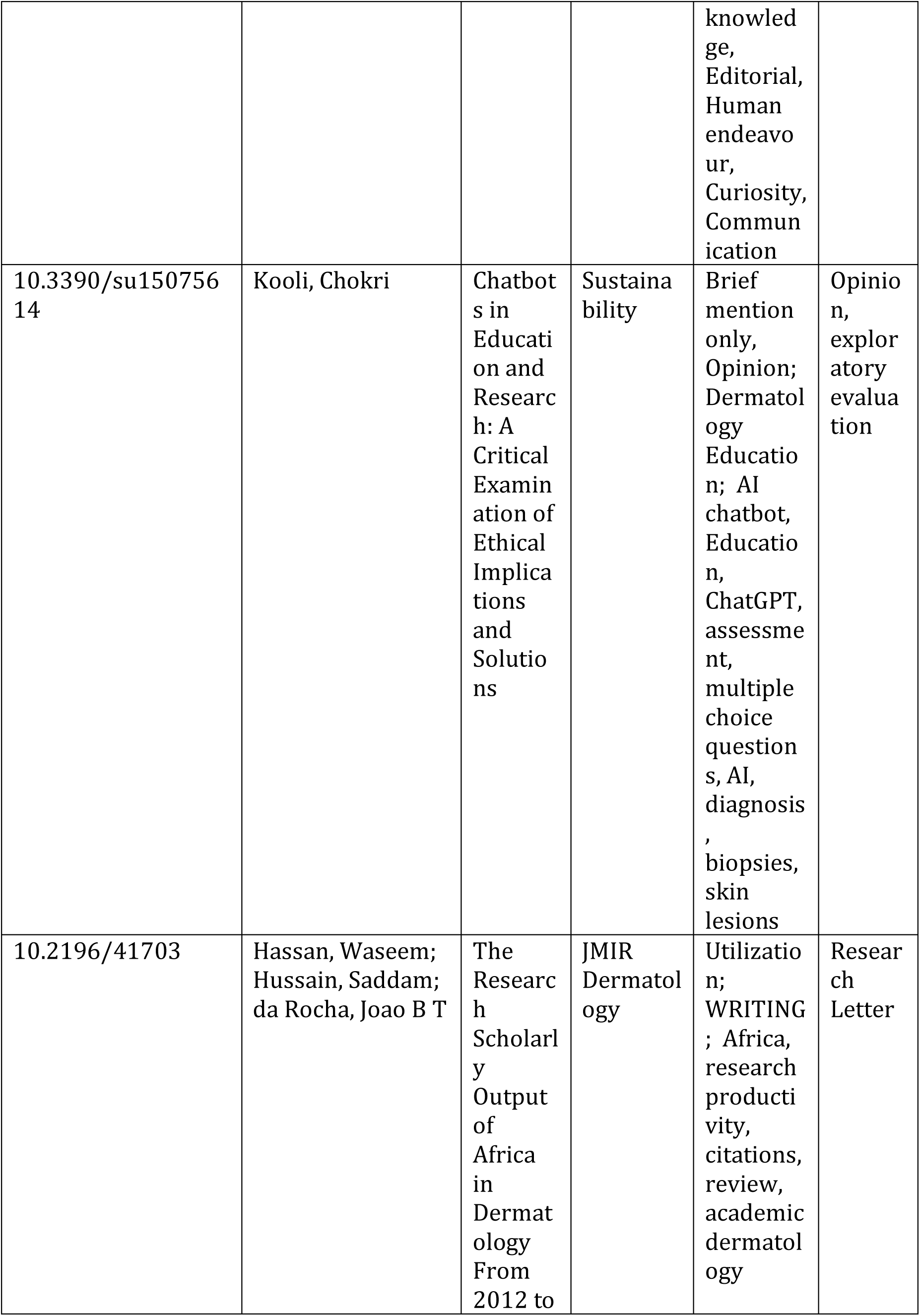
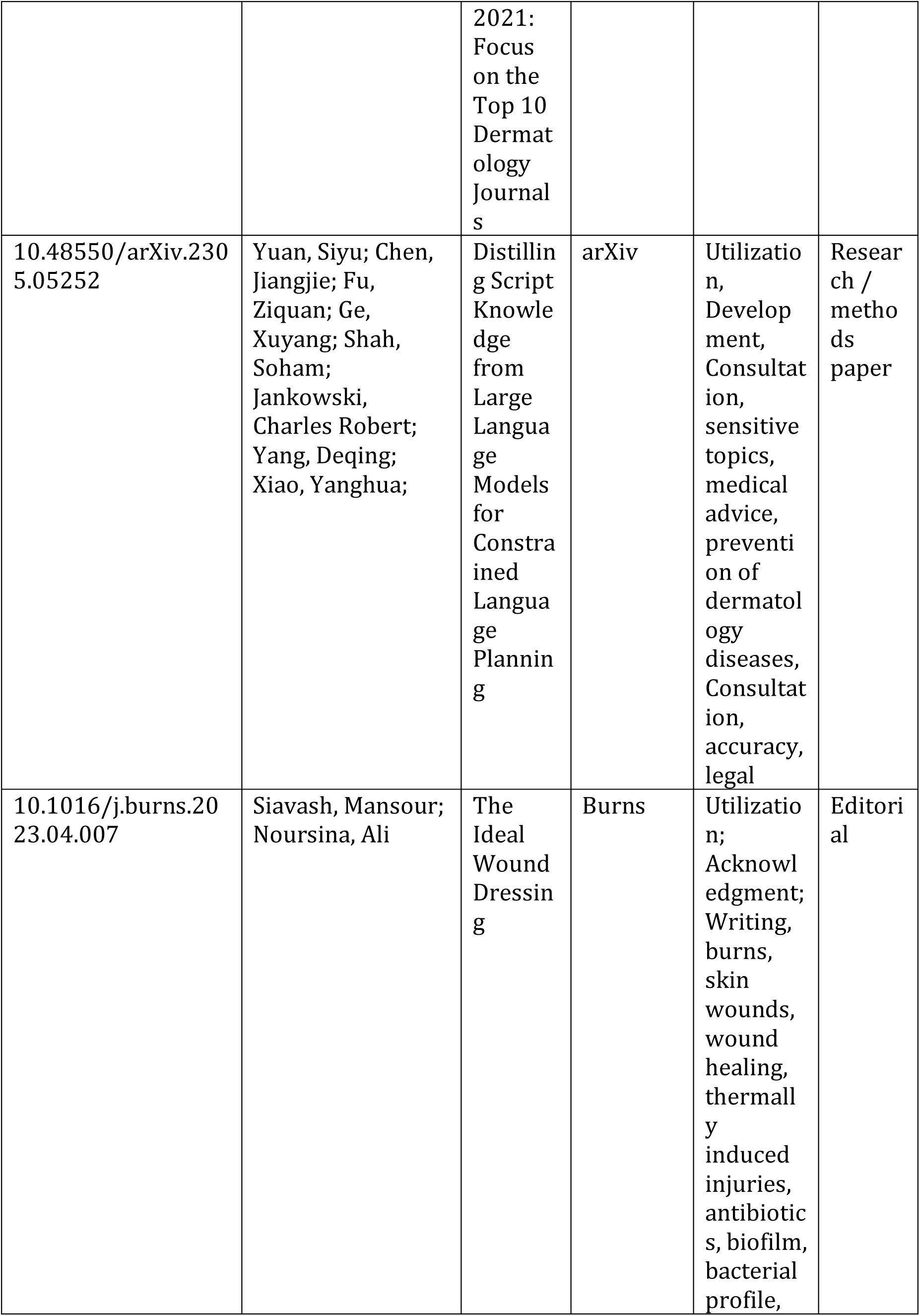
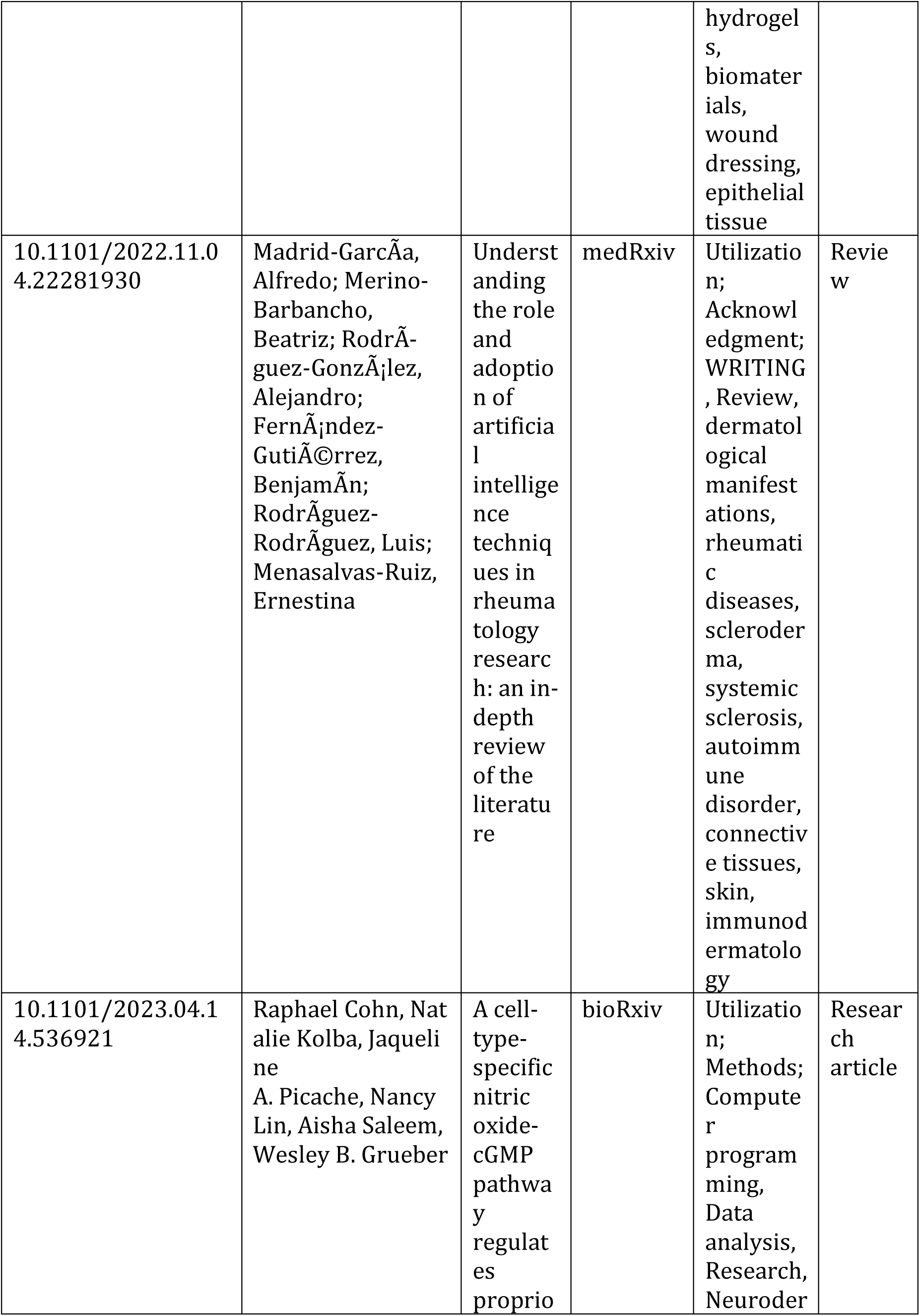
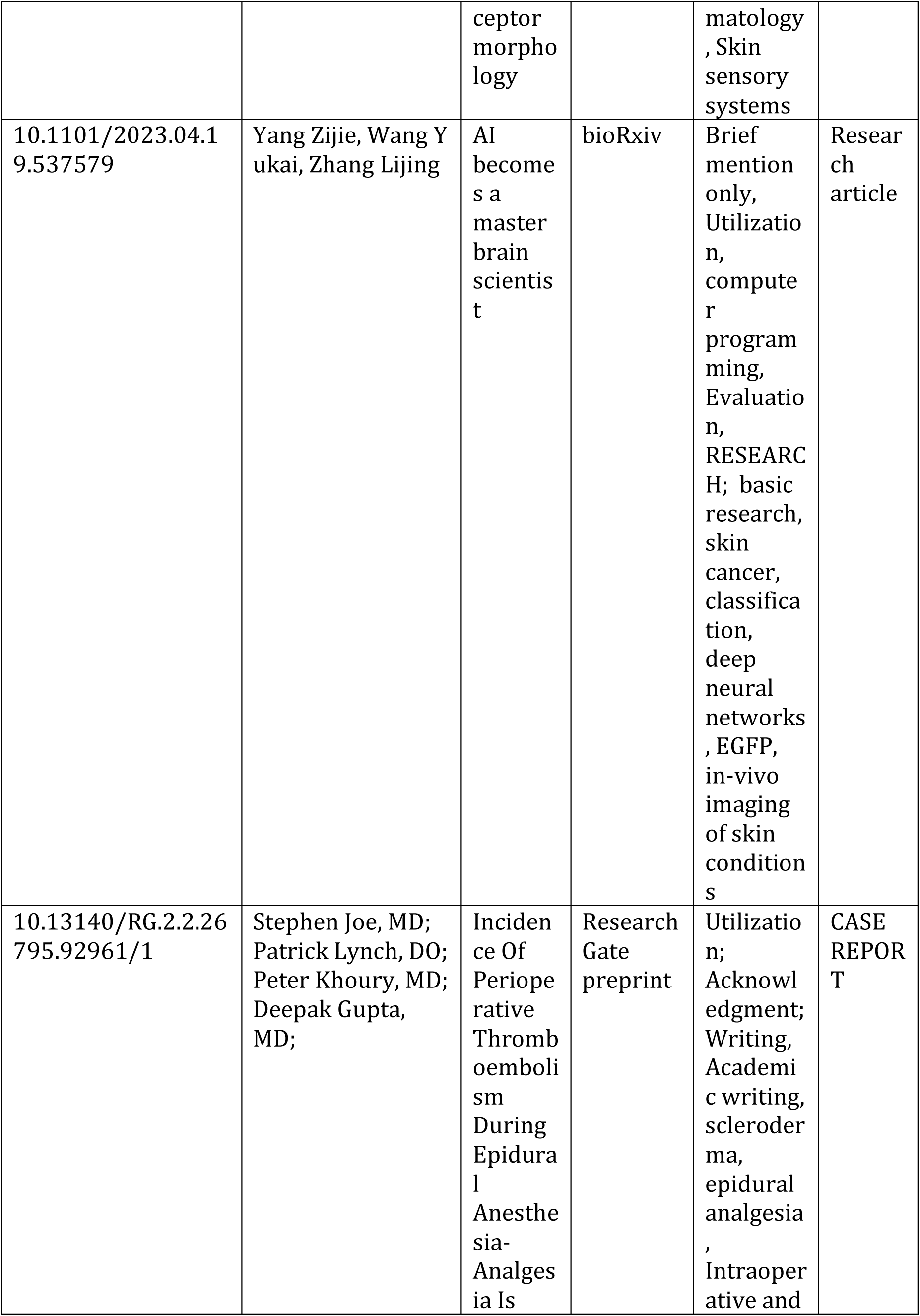
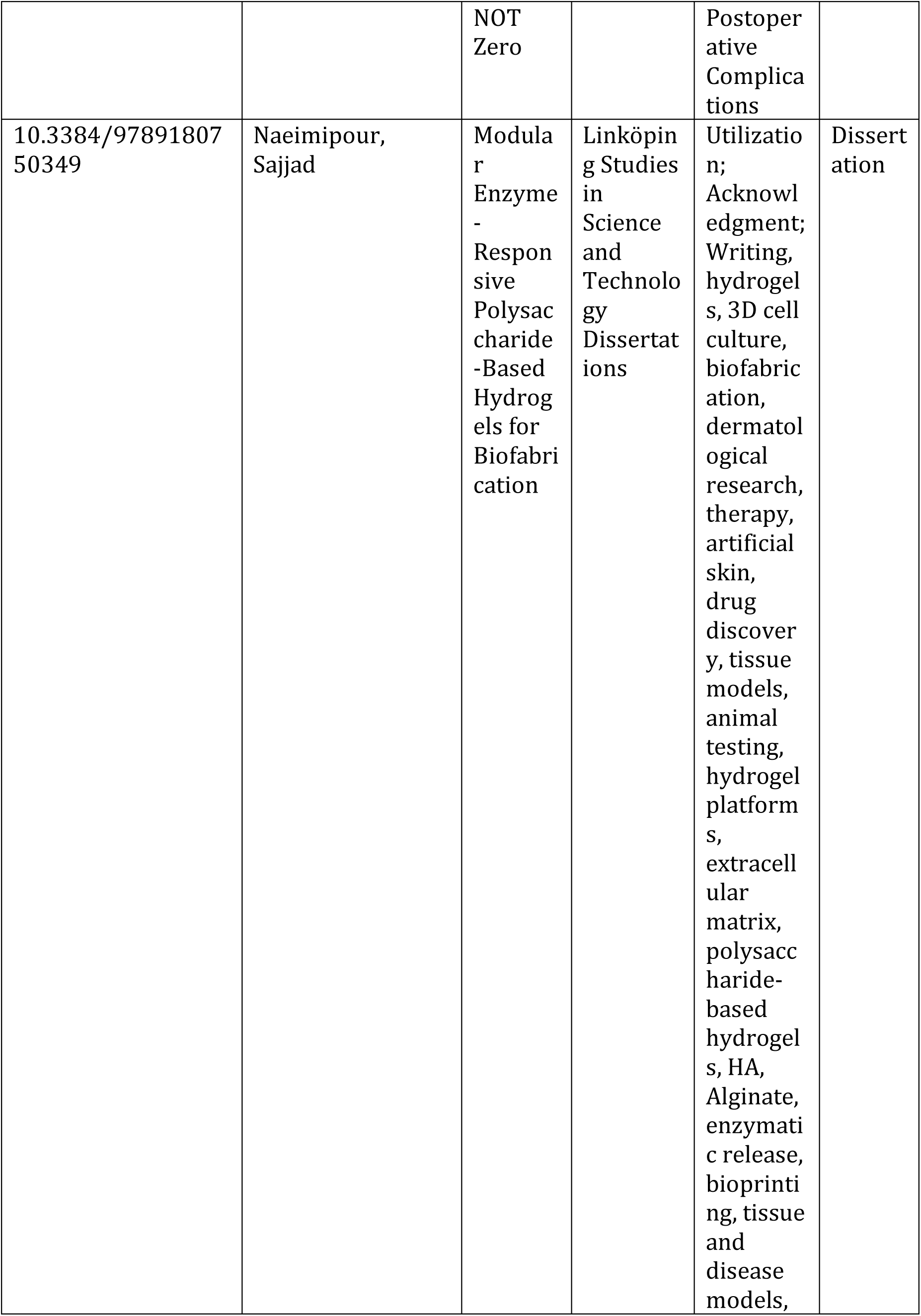
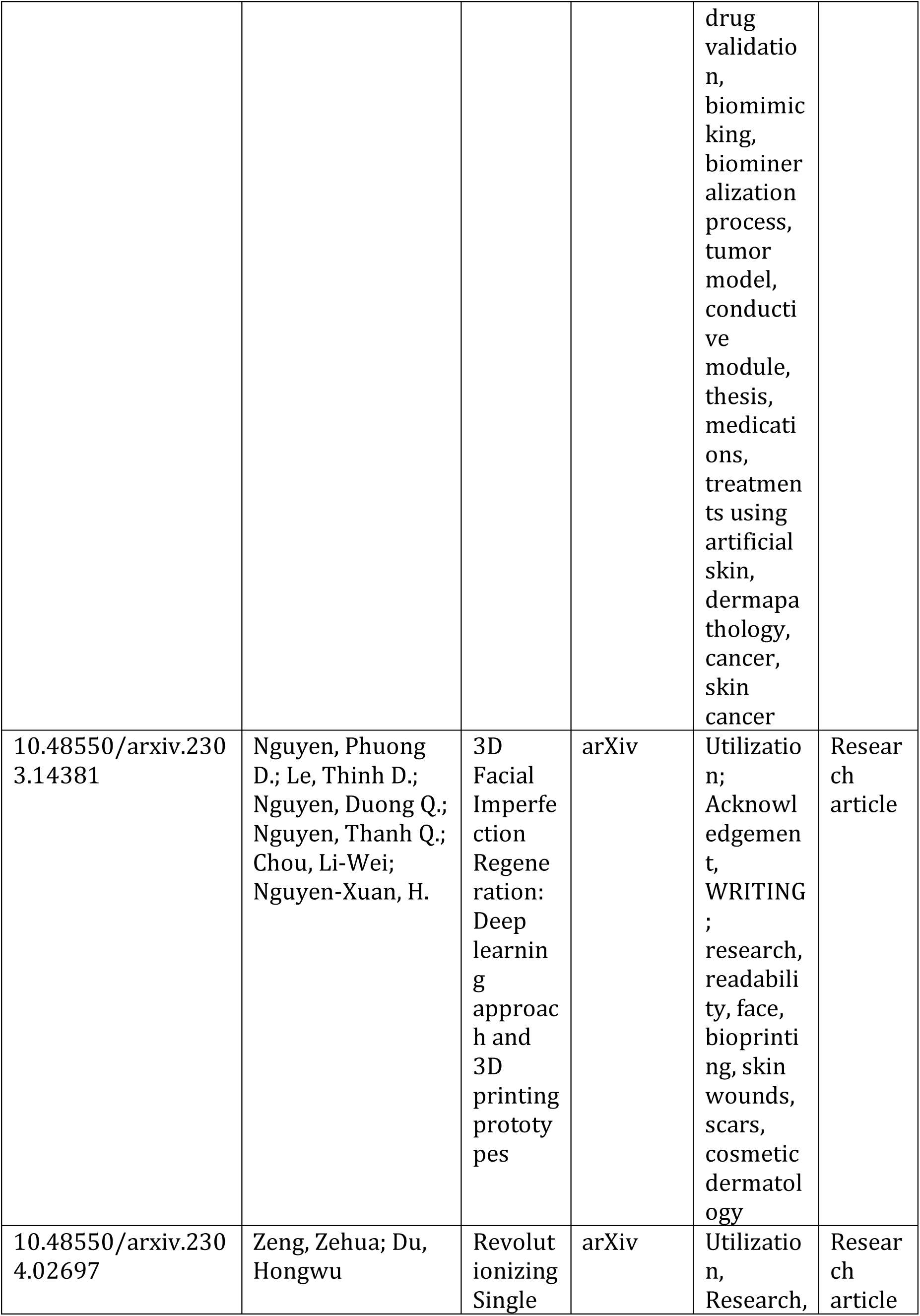
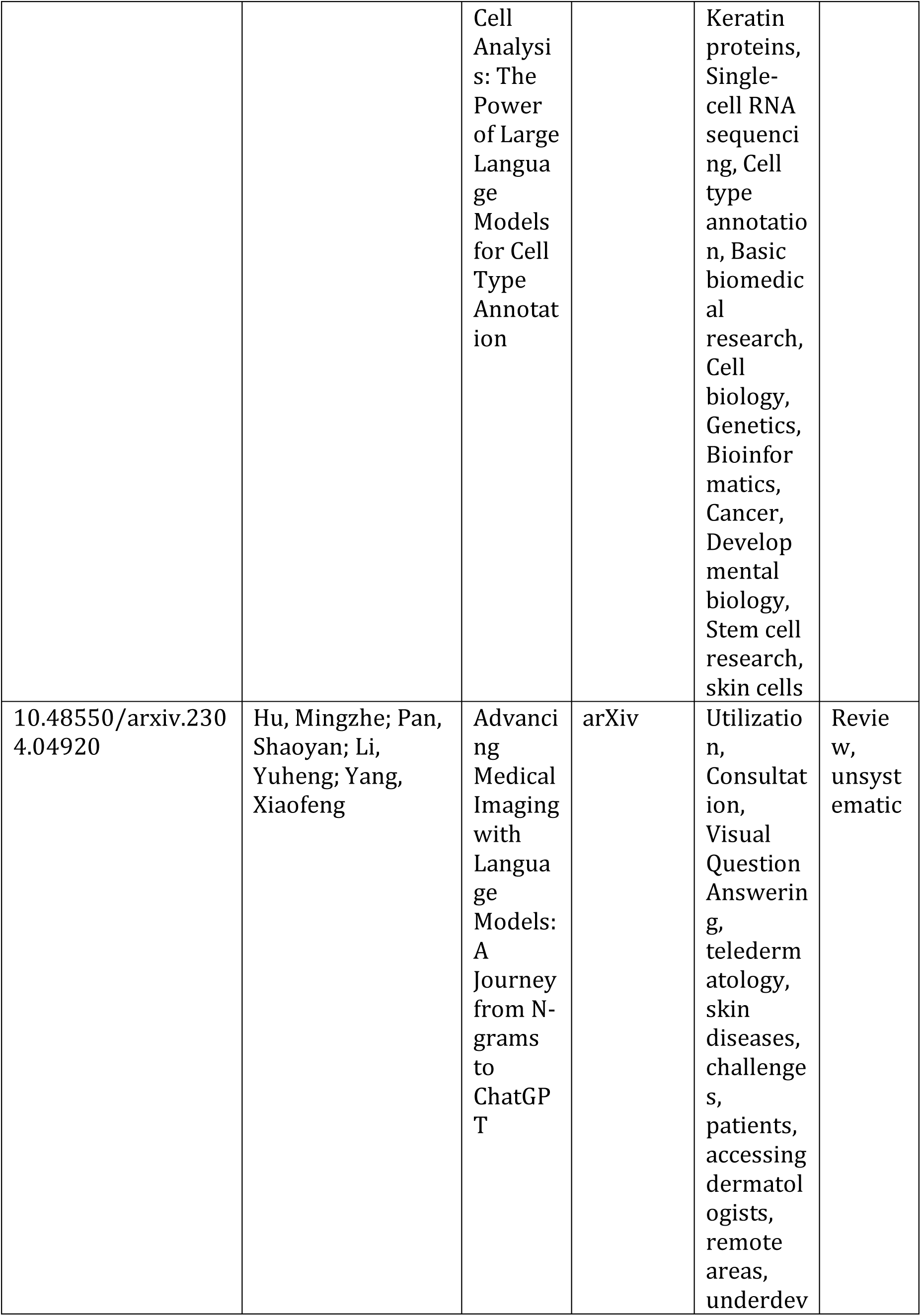
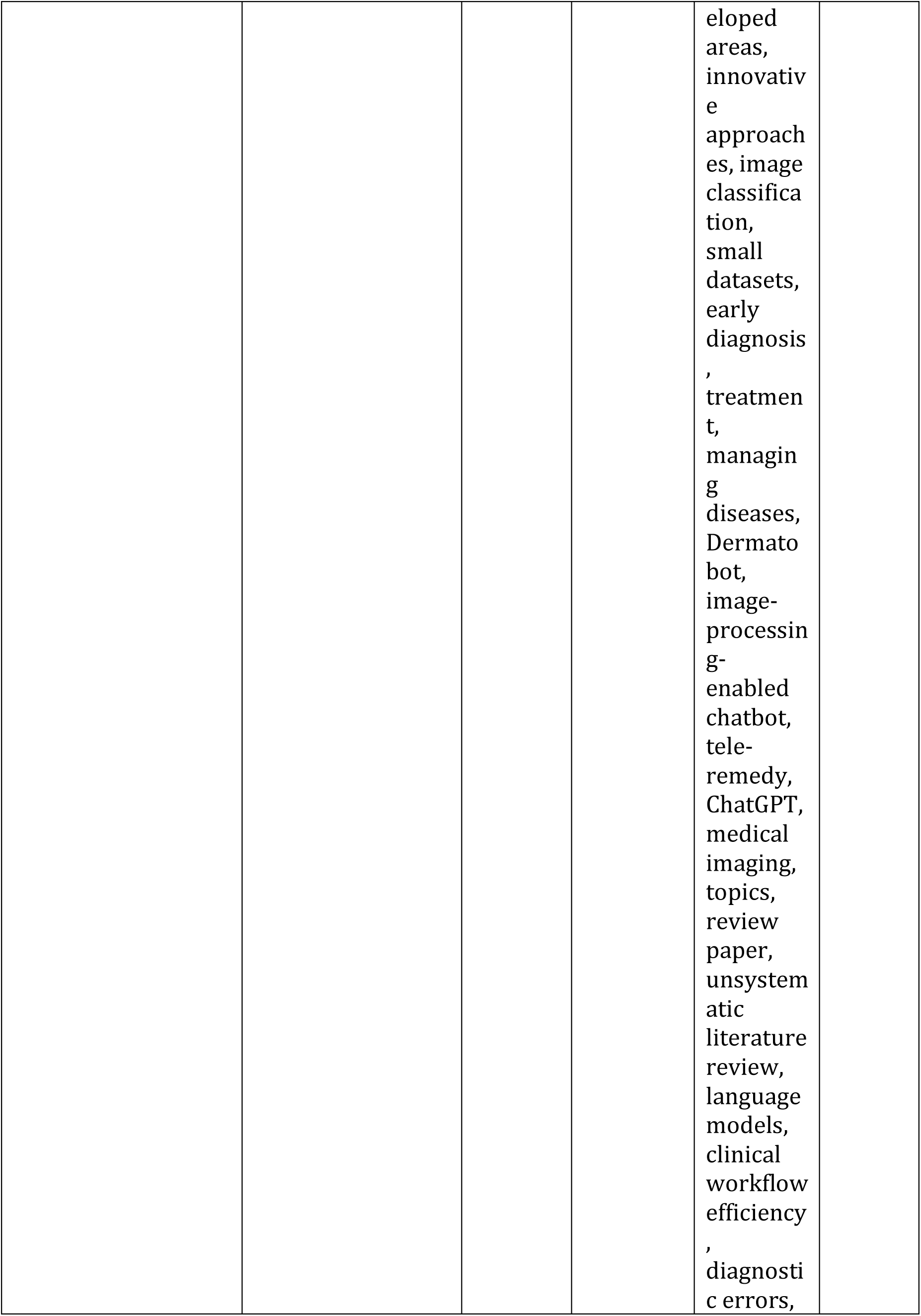
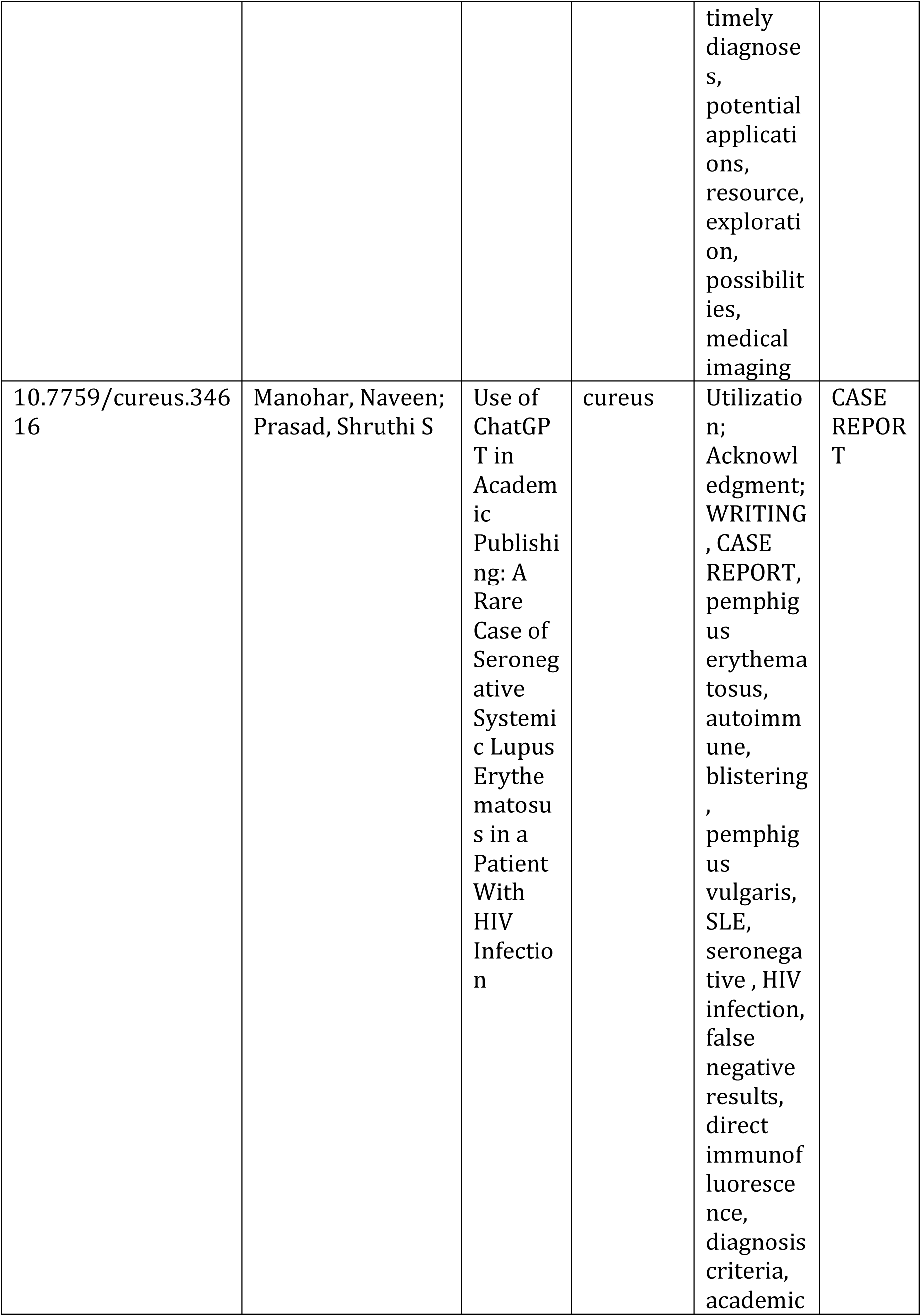
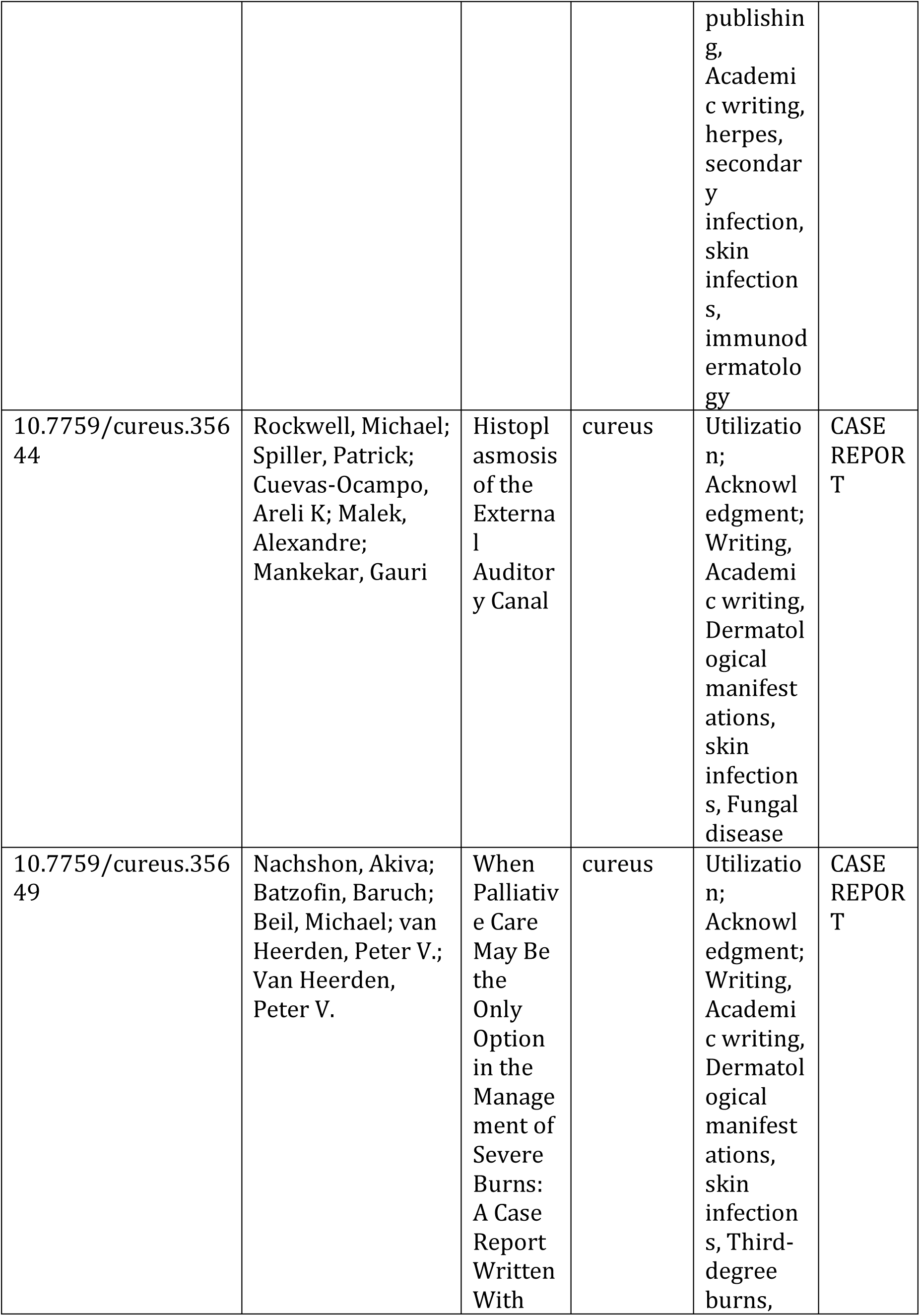
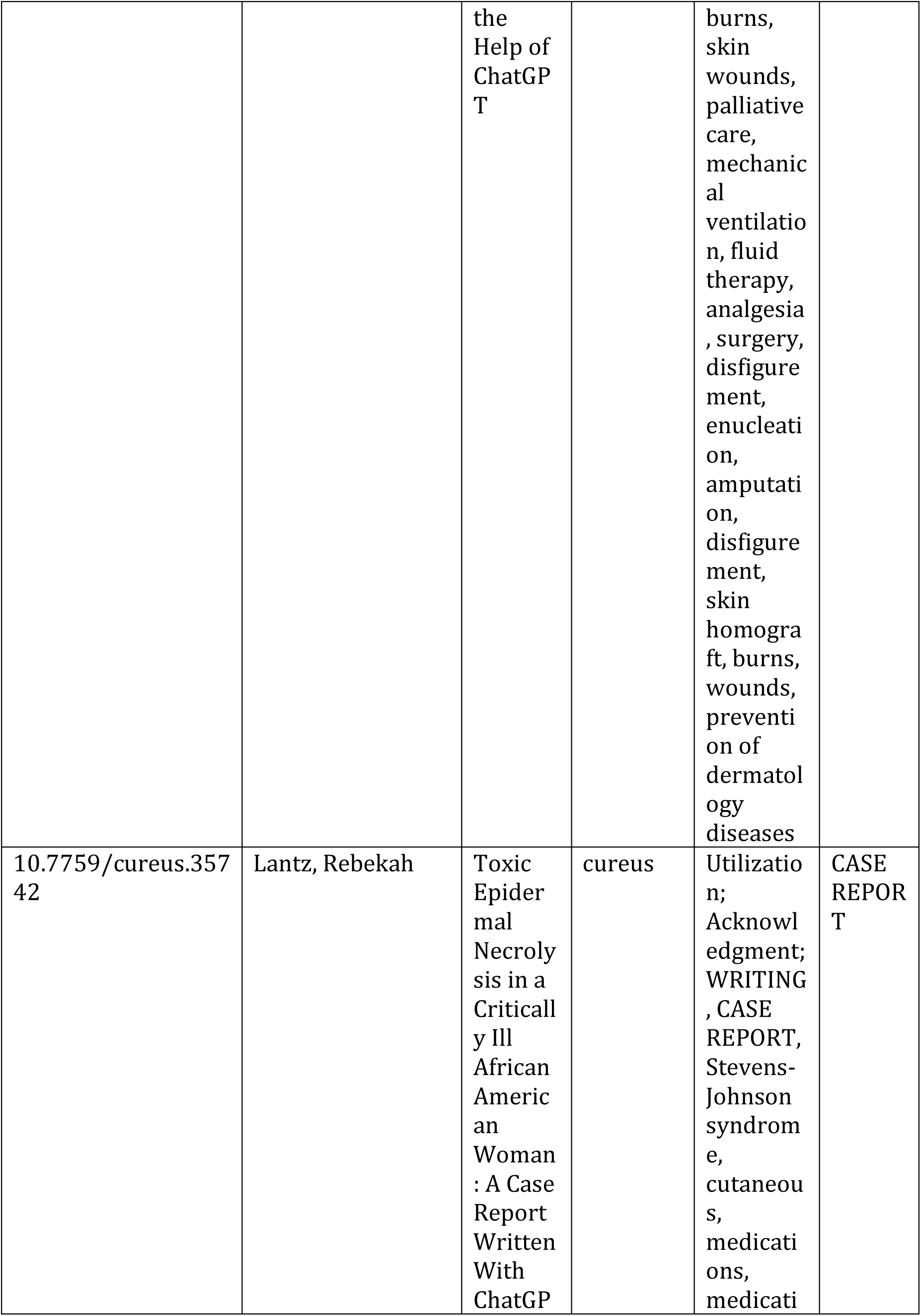
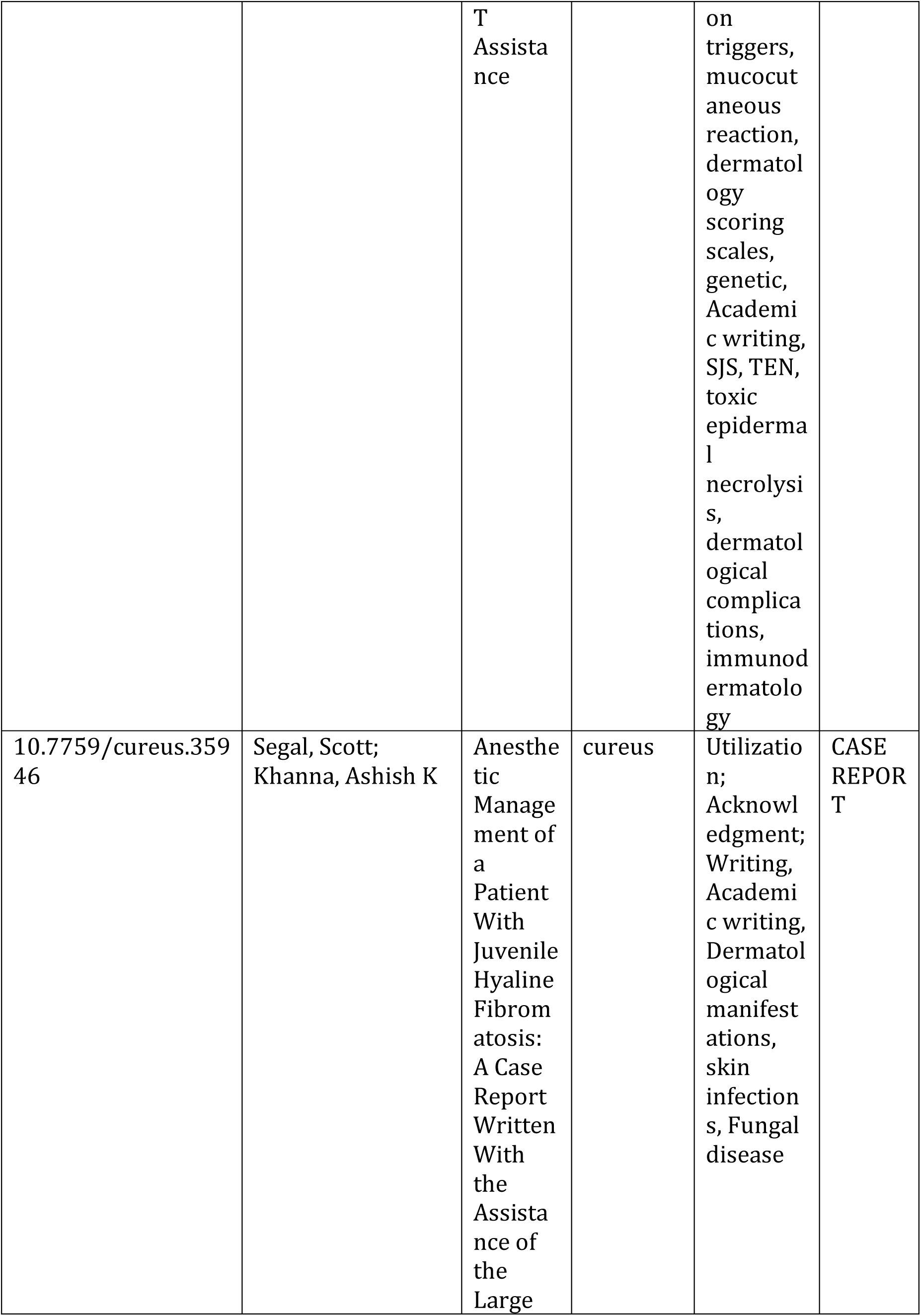
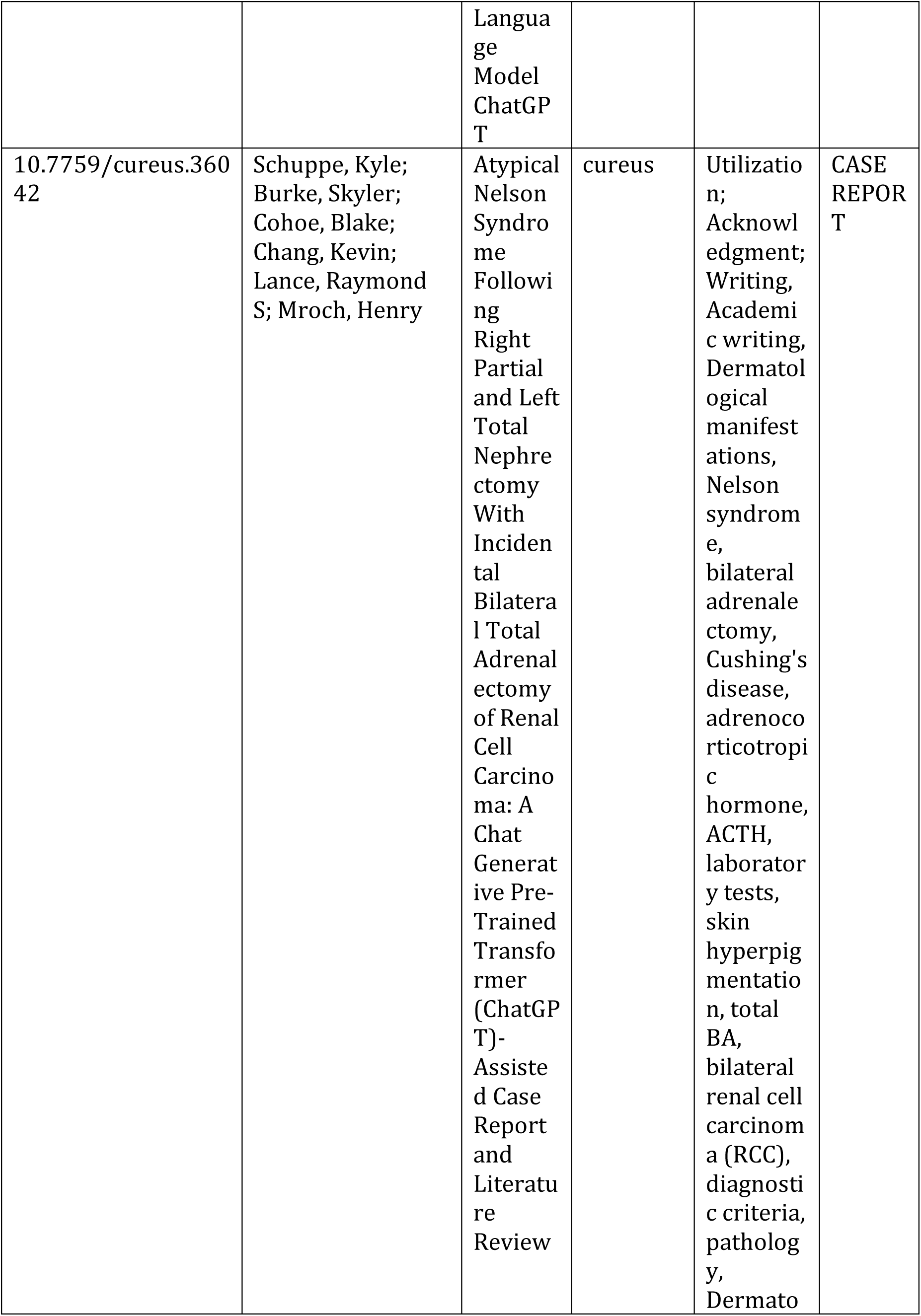
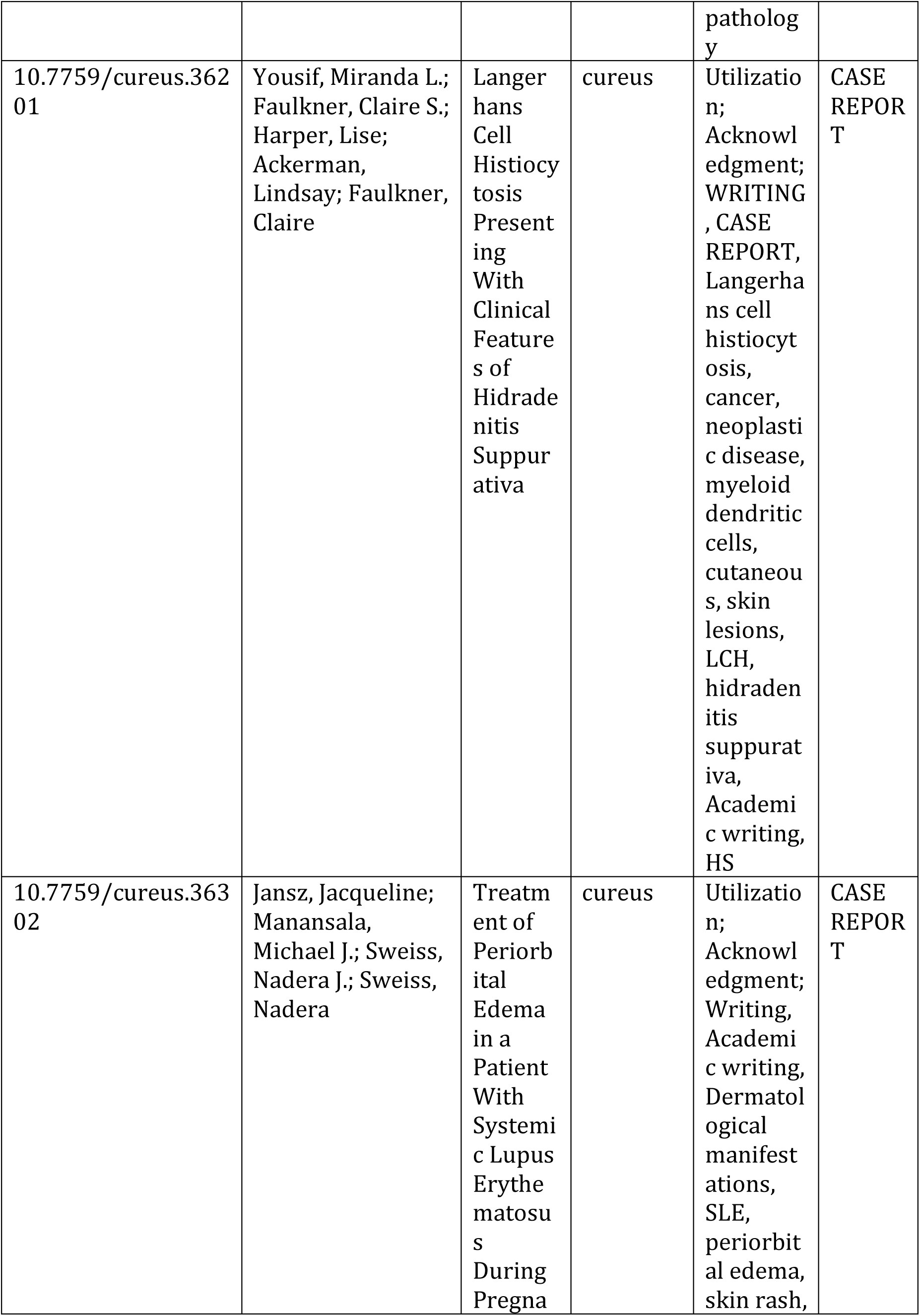
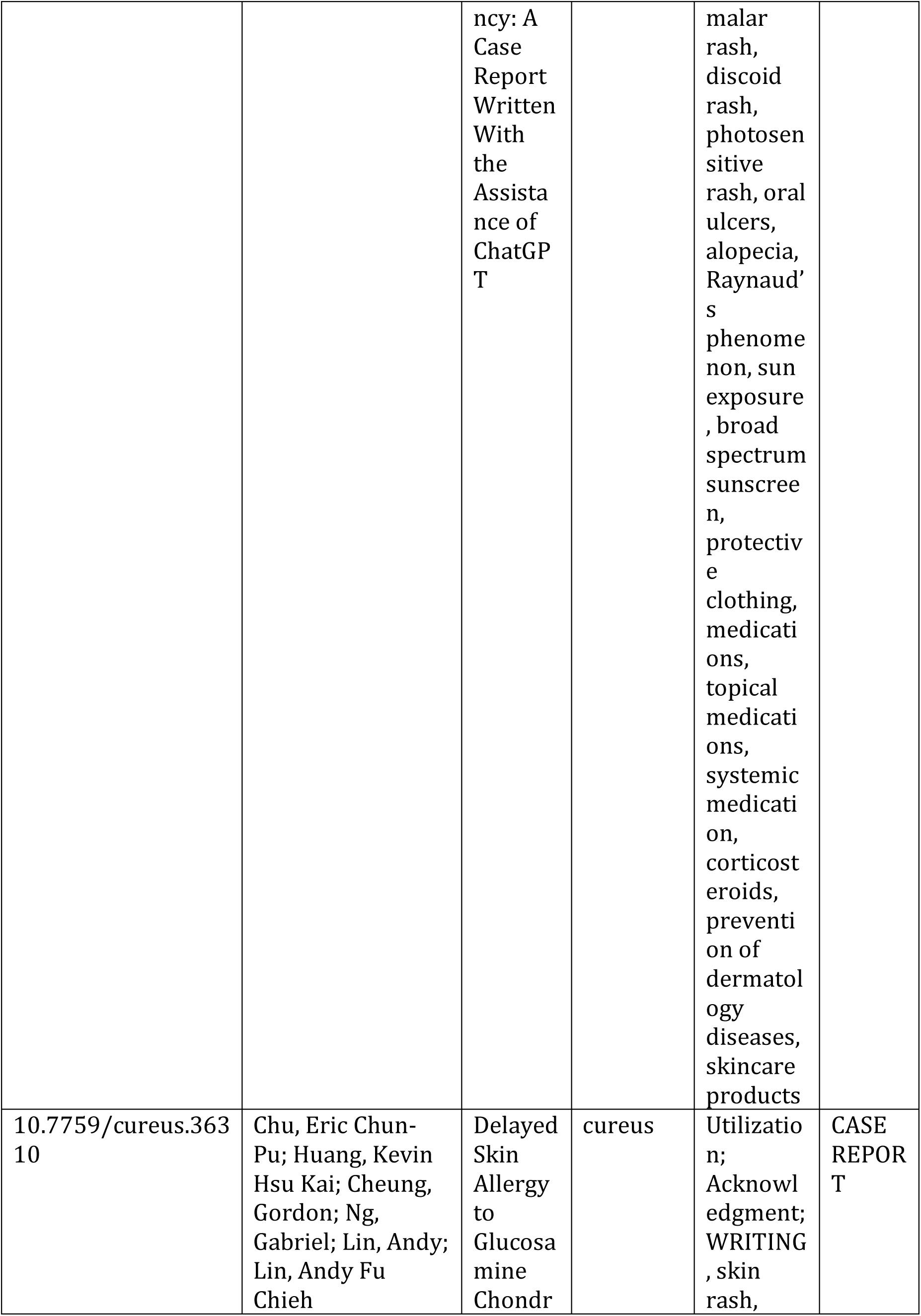
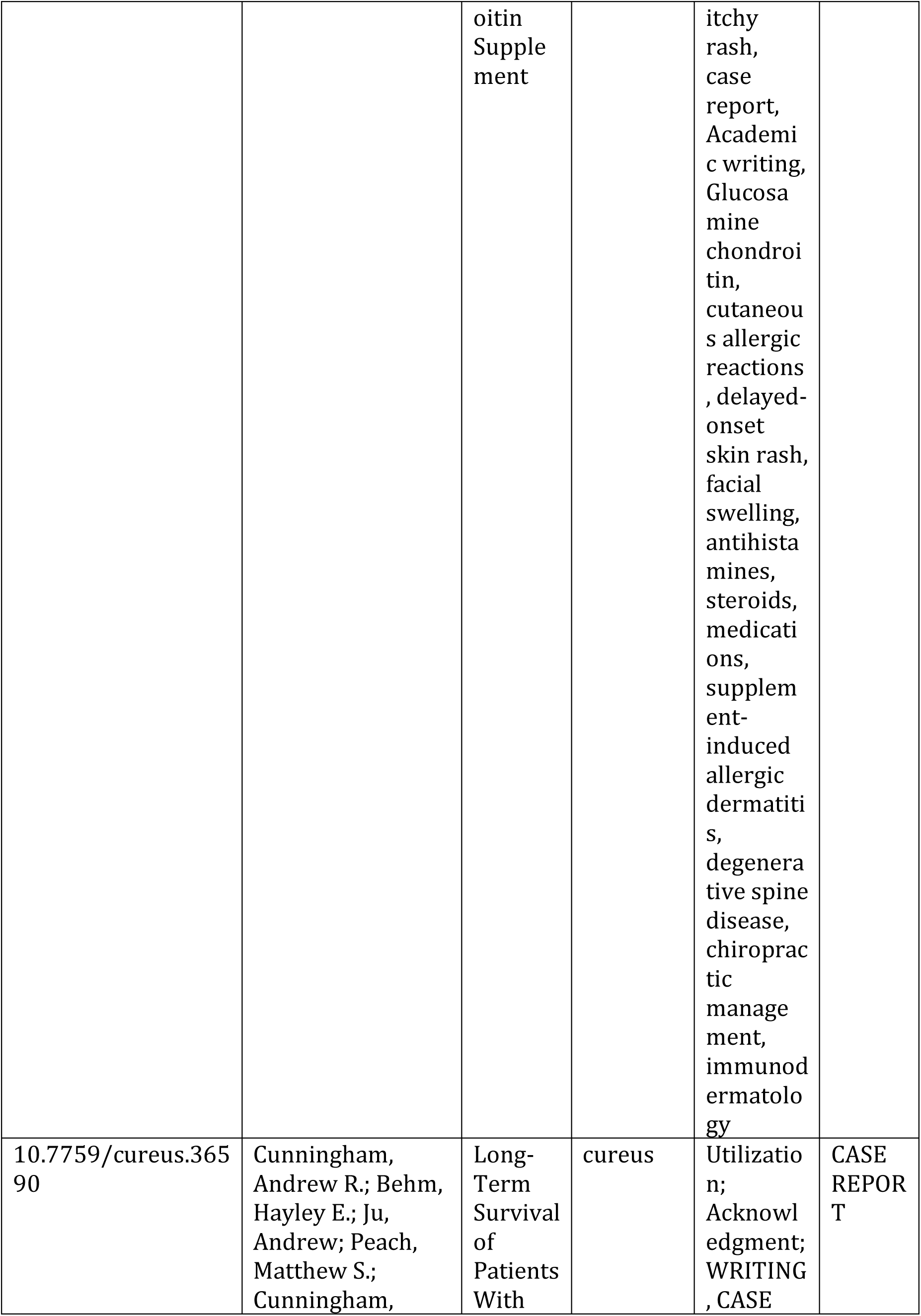
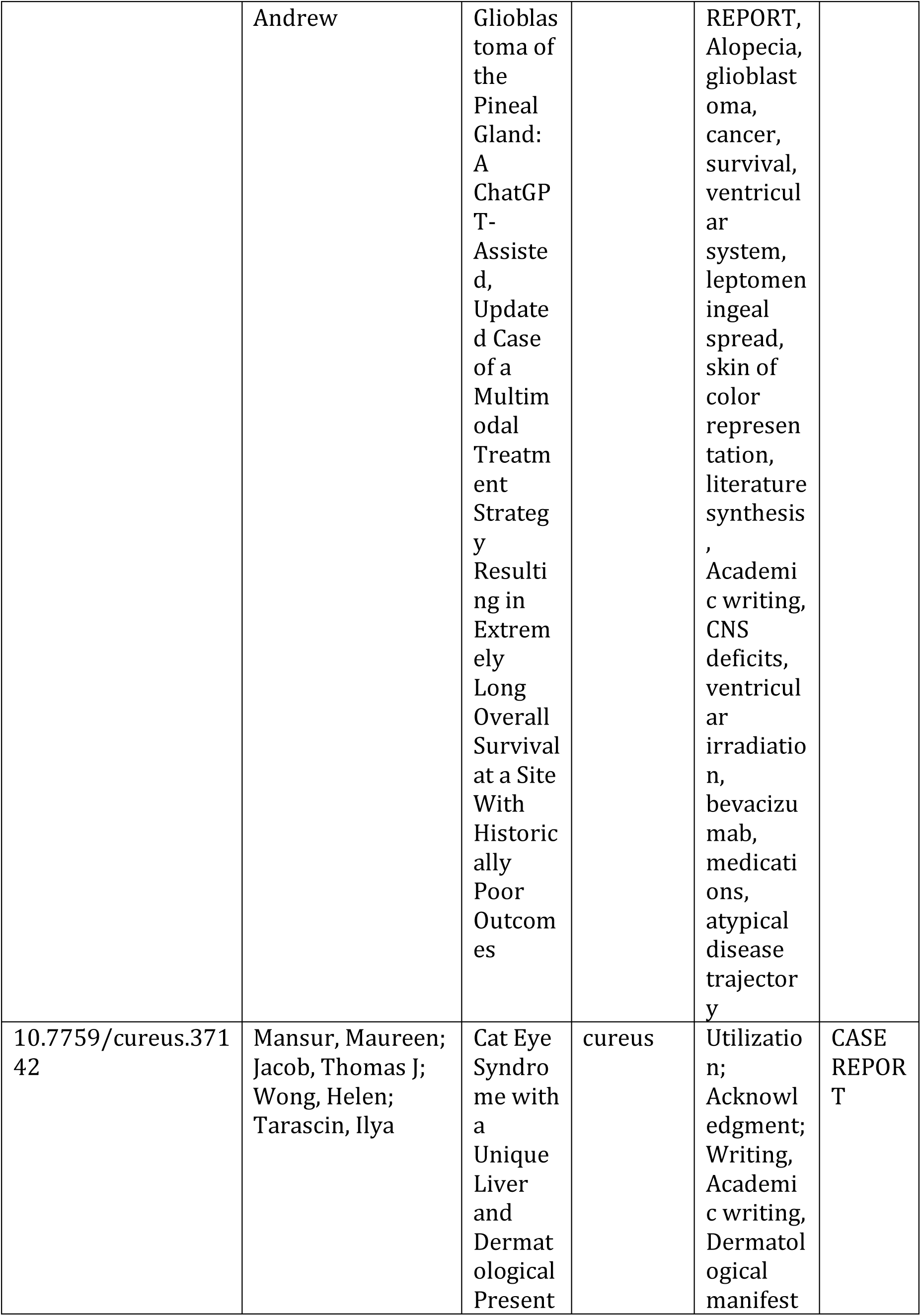
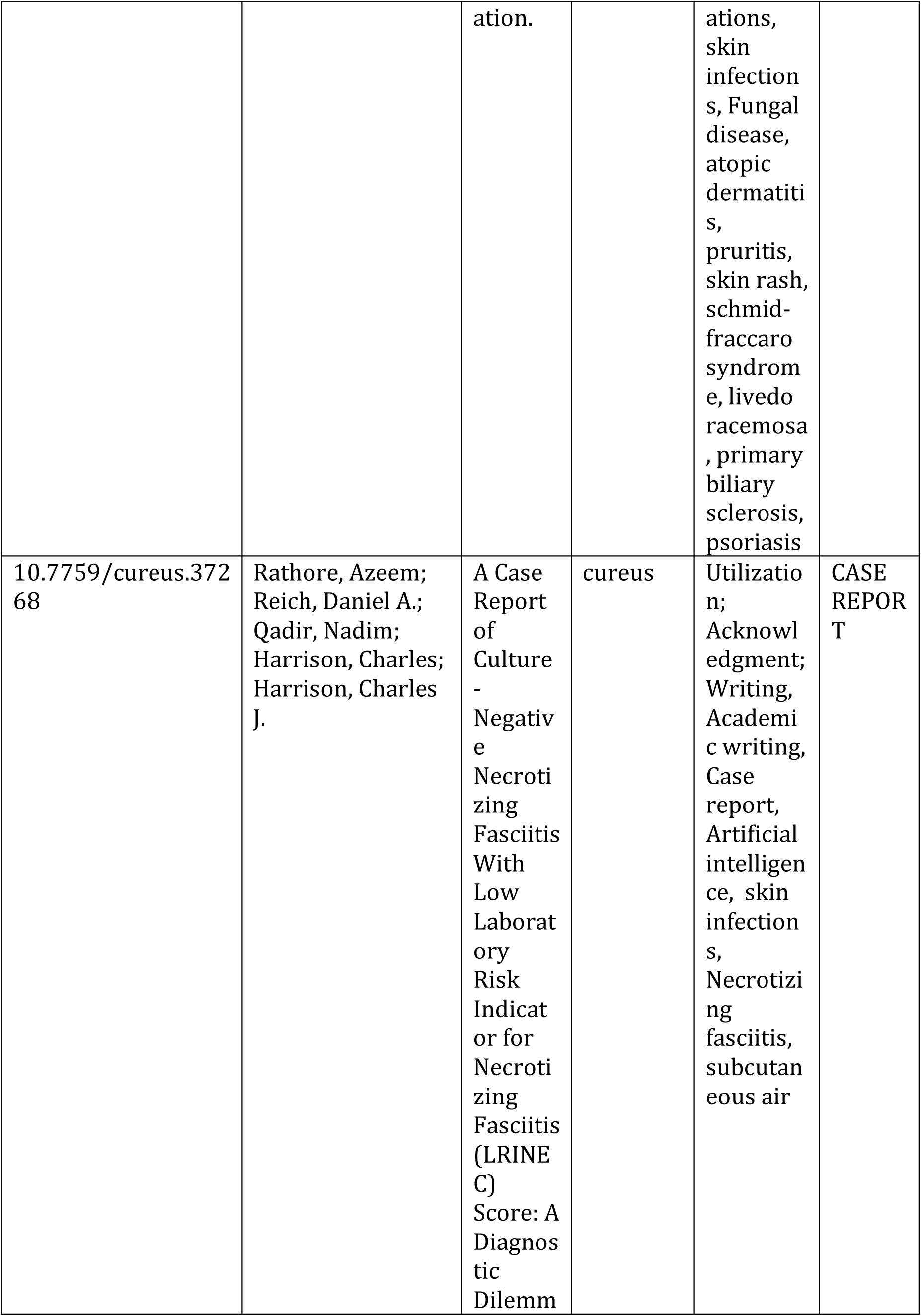
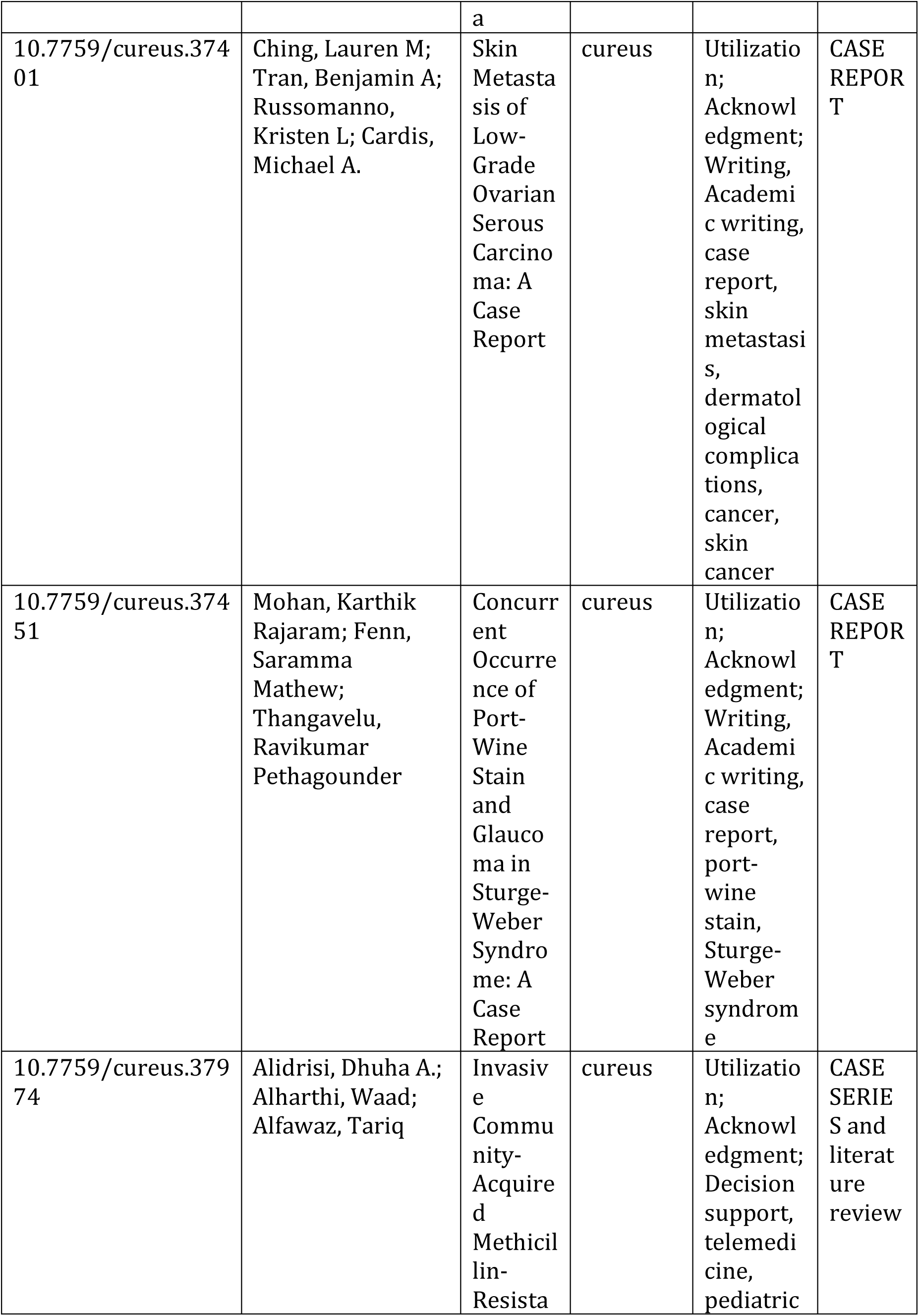
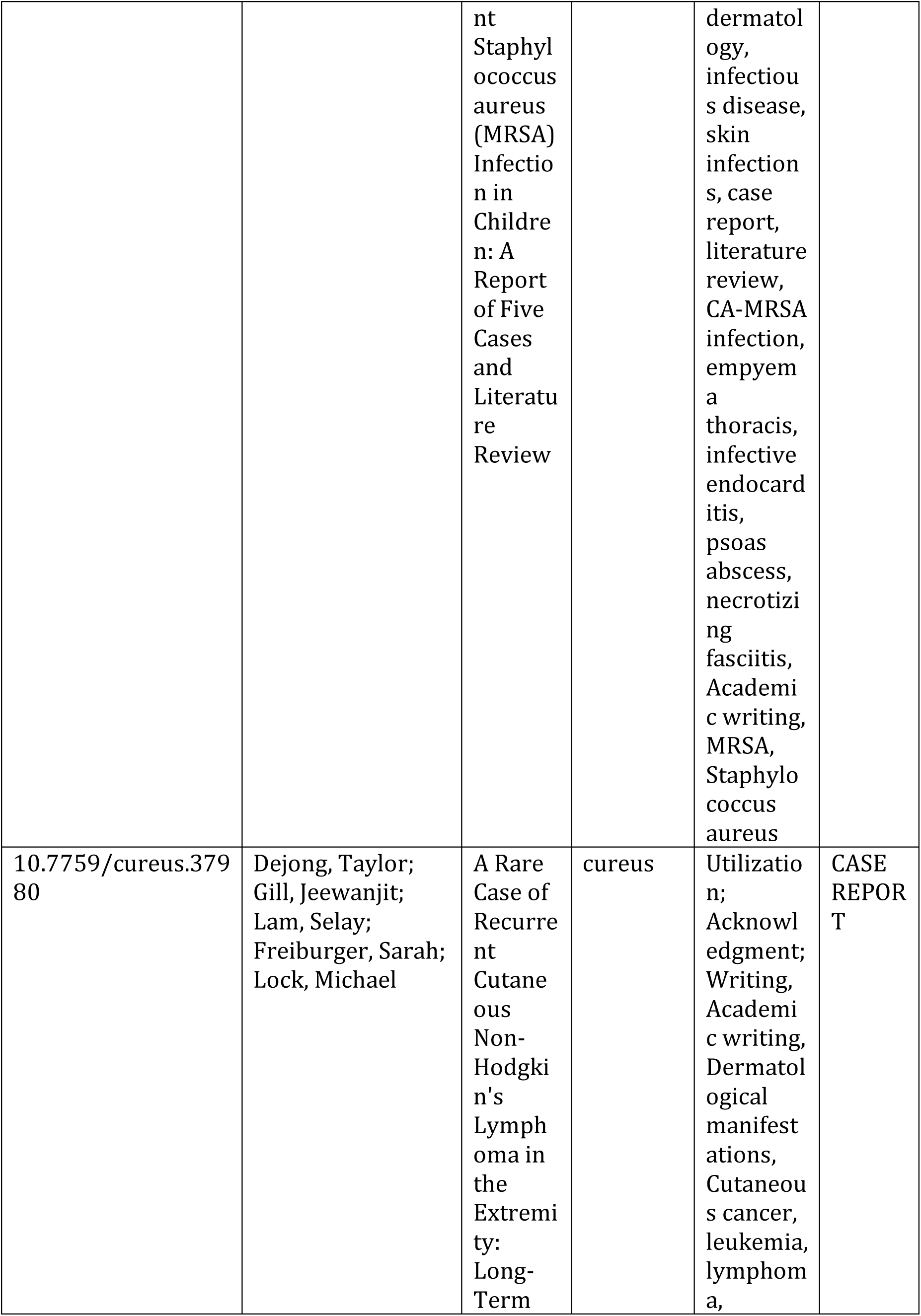
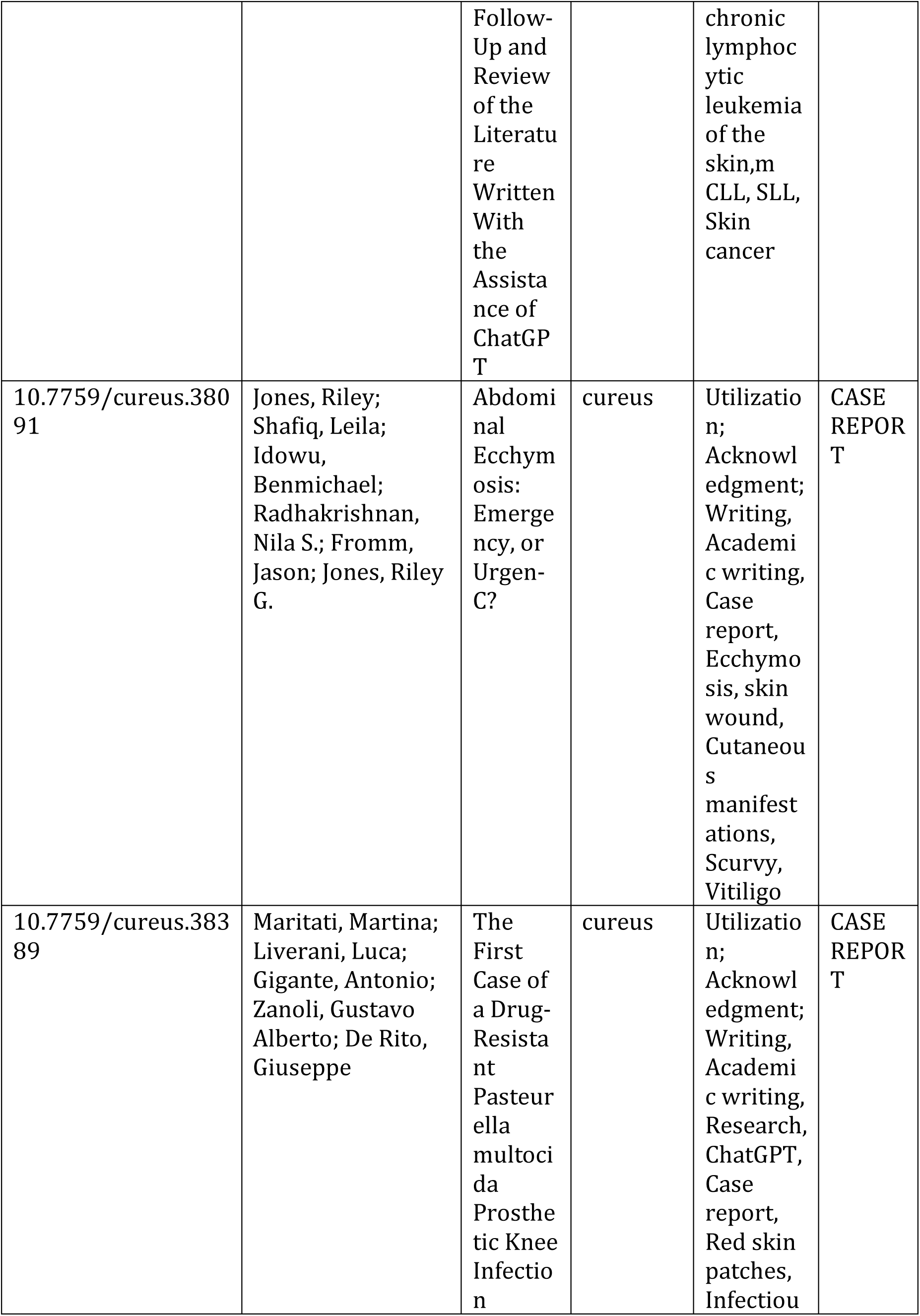
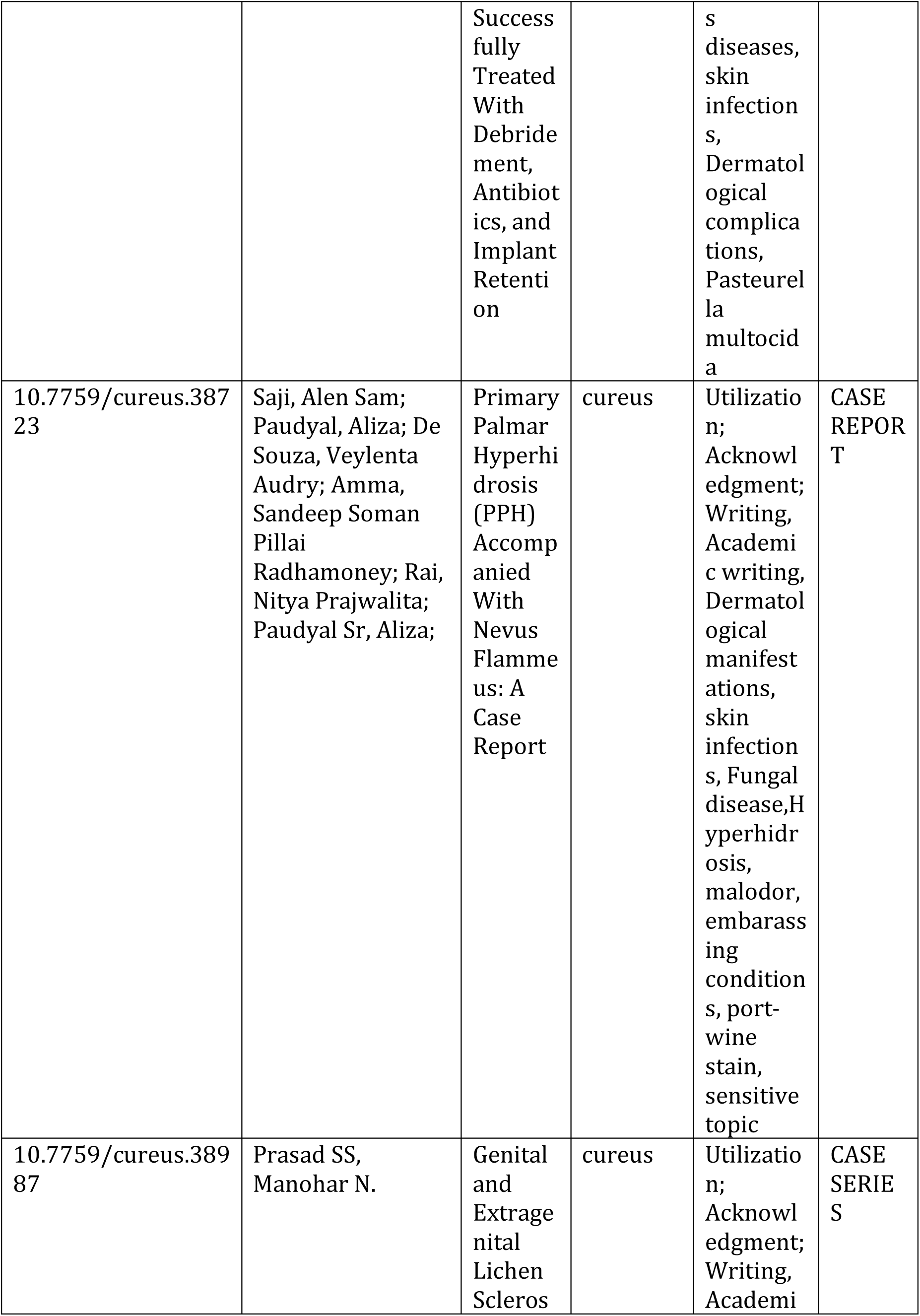
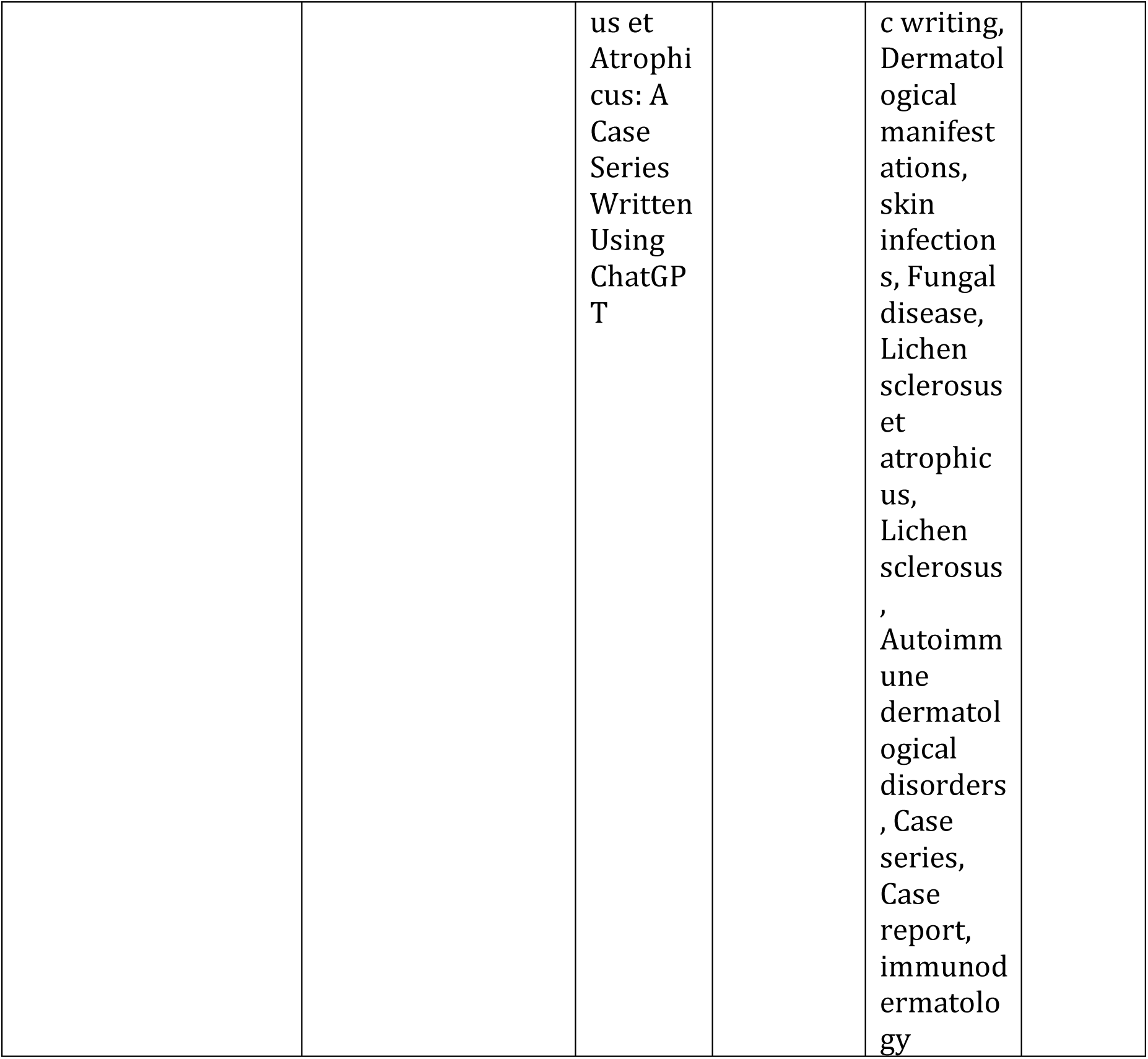

